# The effect of local extended families in epidemics: the case of COVID-19 deaths in the US

**DOI:** 10.1101/2025.08.12.25333440

**Authors:** Jericho McLeod, Eduardo López

**Affiliations:** Computational and Data Sciences Department, George Mason, University, 4400 University Drive, Fairfax, 22030, Virginia, United States

**Keywords:** COVID-19, kinship, epidemic modeling

## Abstract

Do extended family members that live near each other significantly affect the dynamics of large respiratory infectious disease outbreaks? While current epidemiological consensus recognizes household family as relevant, extended family (kin) groups are not viewed as playing an especial role, disregarding evidence of the frequent and unique support they provide each other. Indeed, if extended family was relevant, this disregard would likely lead to deficiencies in advice, modeling, and preparedness during an epidemic as neither the interaction patterns with extended family nor the effectiveness of non-pharmaceutical interventions (NPIs) on them are known. Here, through the creation of the first large-scale data set of obituaries in the US for research purposes (**~ 1.4** million deaths), we show for the first time evidence that between the beginning of 2020 and end of 2022 (the height of the COVID-19 pandemic) there were surges of deaths among kin members living in the same cities, consistently spanning summer periods. The kin surges we observe started simultaneously with overall COVID-19 population surges but continued even as the overall surges subsided. These surge patterns are consistent with extended families maintaining contact even through strict periods of NPIs implementation, potentially making them an unrecognized reservoir of disease spread helping outbreaks persist. Further, we find indications that kin-related deaths are more prevalent among genetic kin and display gender heterogeneity. Our results call for a renewal on research on infection-relevant contact patterns among extended US families and on the effects that NPIs may have towards these contacts.

## 1 Introduction

Established epidemiological literature has identified the main types of social contacts that contribute to the propagation of respiratory infectious diseases at scale such as seasonal or pandemic influenza or COVID-19 [1–3]. These include household (coresident) contacts, work contacts relevant to adults, school contacts relevant to children, and a generic background of contacts referred to as community contacts. In the US and several other countries the first of these, household contacts, are the *de facto* definition of family contacts in the way they are included in large agent-based models [4–6] and two-level mathematical models [7].

Yet, there are strong reasons to believe that equating family with households (the nuclear family focus [8]) may be an incomplete view of this social contact (tie) in epidemiological terms. First, social science highlights the central role played by *extended* family (kin) in supporting kin members through companionship and care [8–13] that is often essential to the daily functioning of households [10, 14]. Accordingly, it is no surprise that a large fraction of people live near extended family in the US (at least 75% [15, 16]). Second, situations such as the COVID-19 crisis, which required the implementation of extensive non-pharmaceutical interventions (NPIs) in 2020 and 2021 across the US, can make local extended family even more essential for some people who may find themselves needing to compensate for the reduction of social contact or services such as childcare (many school systems and day cares for very young children were closed for extended periods of time [17, 18]); the domestic US migration changes seen in 2020 and 2021 aimed at increasing family propinquity are consistent with this [19]. Third, epidemiological modeling approaches to community contacts (which currently include extended family by default) tend to treat them as low-probability homogeneous contacts, incompatible with what is known about extended family which may be more similar to a cyclical contact due to the regularity with which encounters occur [13]. These considerations suggest, especially under the implementation of NPIs, that the role of local extended family in population-level disease propagation needs further investigation to determine how appropriate it is to treat it as a generic community contact, playing a role that is no different than a constant background risk.

There are, however, important challenges surrounding this research problem. If one were to study the epidemiological impact of extended family via modeling approaches, one would quickly find that there is a dearth of data with which to calibrate it into those models. For example, little is known about the way extended family is locally structured into households, which is necessary information to properly model household-to-household interactions. Furthermore, there are indications that such structure may vary in systematic ways [10, 13] but the few data sets that support this conclusion are not sufficiently resolved to determine the details needed for models. Similarly, if one were to analyze the epidemiological impact of extended family empirically, there is yet no clear evidence that, at the population level, such family indeed lead to measurable propagation of respiratory diseases, although this would be at odds with the findings from the contact tracing literature that show that family living in separate households are among the riskiest contacts [20, 21]. What these comments lay bare is that, to clearly determine if extended family plays a role in population scale propagation of respiratory illness (and if so, what role), there is an urgent need for appropriate data.

In this study, we directly address this need for data with an innovative approach that makes it possible to determine statistically significant clustered deaths in short periods of time among extended family that live in the same city. Our focus is the US between the beginning of 2020 and the end of 2022, covering the most active and deadly period of the COVID-19 pandemic. Concretely, we build the largest research database of obituaries across the US to date, capturing approximately 7.59% of *all deaths* between the beginning of 2020 and the end of 2022. Obituaries published in the US customarily record a *decedent’s* name, city of residence, age, date of death, and, when they exist, names of members of their nuclear and extended family. Therefore, when different members of an extended family die and obituaries are written, the names in common across obituaries make it possible to detect the family connection, effectively pairing the obituaries. By counting statistically larger-than-expected numbers of pairings of obituaries, we are able to provide conservative estimates of kin-connected fatalities. We do this as a location-by-location process, where any obituary pairings of people in different locations are ignored, and then aggregate those local analyses into a national picture. This approach helps test if extended family play an important role in community disease transmission.

We find that kin-connected deaths are clearly detectable at the population level. Furthermore, during the COVID-19 pandemic, they displayed a unique temporal trend that grew alongside certain overall COVID-19 surges in deaths but then persisted independently even when the surges subsided. This temporal pattern is consistent with local kin ties independently sustaining disease spread during periods of the pandemic that have otherwise been considered of low risk of contagion. Our estimates show that surges in local kin-connected deaths consistently affected summer periods and had a maximum contribution to the contemporaneous overall US deaths in mid-2020 of no less than 3.45%, although this estimate is almost certainly an undercount. We present demographic analyses across age, gender, the types of family connections (spouses, genetic kin, or affinal kin acquired through marriage) among kin-connected deaths under several estimation scenarios. The analyses indicate that kin-connected deaths are by far more common among genetic kin, that gender differences are detectable among these deaths with a slight disadvantage to women, and that the these deaths begin to be observable from age 40 onwards but peak for the elderly. Finally, possible confounders are checked and discarded as being able to explain kin-connected deaths.

## 2 Results

### 2.1 Extracting kin-connected deaths

We begin by describing key information about the obituary data set (OB, see Methods) and what we can extract from it. After pre-processing and applying criteria to ensure our analyses use well-sampled cities, our sample includes 824, 576 obituaries located in 276 cities. These cities contain 77.7% of the 2020 US population.

Regarding the information we can obtain from each obituary, the most unique element relevant to our question is the set of names of the decedent’s family members.

We extract this information through an ensemble approach that identifies names and relationships in each obituary using a large language model (see Methods). When obituaries for different decedents share at least *m*_*o*_ names in common (which may include the decedents) and are separated by a small time window Δ*θ* of the order of days or a few weeks, they potentially signal the occurrence of deaths caused by a deadly infectious disease moving across kin. We denote the event of finding common people across obituaries as *obituary overlaps*. In the language of networks [22], one can conceive of obituaries as nodes and overlaps as links; possible *kin-connected deaths* due to infections are captured by those links. Furthermore, one can assess the ongoing contribution of kin-connected deaths to the overall death toll of a disease like COVID-19 by analyzing a time series for the formation of overlaps.

The main challenge to overcome to extract useful data from obituaries is that overlaps do not always reflect infectious disease effects among kin, but may instead have other origins. We can distinguish three separate types of effects: kin overlaps due to connected effects (which include the spread of deadly diseases among kin, but also the widowhood effect [23], and joint risks such as car accidents [24]), kin overlaps due to general trends (e.g. family members dying in periods of high mortality), and purely random overlaps due to the fact that names are not unique identifiers. We call the first effect *kin-connected overlaps* because the kinship relationship plays a role in *connecting* the deaths; we label the combination of the second and third effects *continuing overlaps* because they are ongoing and not related to the kin connection *per se* (see Methods for details). Continuing and kin-connected overlaps mix together into *observed overlaps* but, to identify the relevant signal, the continuing contribution must be excluded as it represents an additive noise.

Broadly, our method to generate the estimate of kin-connected deaths is as follows: for each location in the US that is sufficiently well-sampled in our data, we create a time series based on observed overlaps among obituaries in that location and subtract from it a baseline time series of the location’s expected continuing overlaps. Both observed and continuing time series count each single overlap through a weight that estimates the number of overlapping deaths it represents; in this sense, our time series are weighted but we omit the term for brevity. Overlap weights include a parameter that controls for the possibility of correlated behavior on obituary writing within families.

The subtraction of the local continuing overlap time series from the observed overlap time series is the time series of estimated *local kin-connected overlaps*. Because of the equivalence of overlaps and deaths, we describe local kin-connected overlaps as *local kin-connected deaths* (LKDs). Location specificity ensures we only account for local kin-connected events and controls for the variability of sampling in each location. Finally, we aggregate the results for all well-sampled cities in the US to create the time series of *aggregate local kin-connected deaths*, Eq. 1 (details are found in Methods). Since we typically focus on the aggregation of LKDs from this point, we use this acronym to refer to the aggregated time series.

Representing month *t* of the time series of observed overlaps as *s*(*g, t*) (Eq. 7) and of expected continuing overlaps as *r*(*g, t*) (Eq. 8), both for location *g*, the estimated number of aggregate LKDs is given by

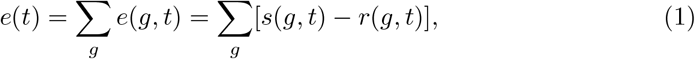

where *e*(*g, t*) = *s*(*g, t*) − *r*(*g, t*) are the LKDs in *g*. It should be noted that when referring to month *t* throughout this work, this specifies the year and month of interest. We abuse notation for *e* by using the same symbol for a specific location *g* or summed over locations (in this latter case, *g* is omitted as one can see on the left-hand side of Eq. 1). Equation 1 implicitly contains parameters *m*_*o*_, Δ*θ*, and a set of values *α*_*z*_ quantifying the correlated behavior for obituary writing within families when two relatives are related as *z* = genetic kin (GN), affinal kin (AF), spouses (SP), or, infrequently, there are indeterminate cases (ID) for some relationships (see Methods). The results presented in the main text are representative and use *m*_*o*_ = 4, Δ*θ* = 30 days, and *α*_*z*_ values as explained below. We use *M* = 100 realizations to obtain the average baseline *r*(*g, t*) and apply sampling criteria to ensure that we only include cities with reliable samples (see Methods). Robustness checks of parameter choices are presented in the Supplementary Methods S1.5.

To aid in the interpretation of the results presented next, we discuss *e*(*t*) as defined in Eq. 1. Note first that our method measures location-specific overabundance of obituary overlaps (*e*(*g, t*) = *s*(*g, t*) − *r*(*g, t*)) above statistical expectation (*r*(*g, t*)) and, because overlaps rely on shared names of kin among obituaries, the overabundance is kin-related. Therefore, our approach is essentially the detection of excess signal in LKDs in a similar way to measurements of, for example, COVID-19 excess deaths [25]. Our method does not directly find chains of infections and therefore cannot be used in its current form as a contact tracing approach (this is also true for methods of COVID-19 excess death estimations). Second, and related to the first point, the estimate provided by *e*(*t*) includes all causes of deaths as obituaries are not specific to any one cause of death. In this sense, LKDs estimated by our method include individuals whose fatalities may be directly due to COVID-19, due to combinations of factors such as COVID-19 and decedent frailty (likely present among decedents in our sample which tend to bias towards the elderly as we show in Supplementary Methods S1.4.1 through S1.4.3), or due to other unobserved factors. However, it should be noted that because of the temporal pattern of *e*(*t*), it is strongly supportive of COVID-19 being its main contributing factor (explanations provided below). Third, we reiterate that *e*(*t*) estimates numbers of excess *overlaps* and not numbers of *all* deaths involved in those overlaps. Concretely, although an overlap involves two obituaries, it is the overlap itself that we count as this is what represents the possibility of a connection between the two deaths involved. Fourth, we emphasize that our weighing scheme, fully explained in the discussion leading to Eqs. 5 and 6, means that *e*(*t*) has units of numbers of deaths that we attribute to kin-connected events.

Concluding this section, we address the parameters *α*_*z*_ (in principle, one for each *z*), necessary for the calculation of *e*(*t*). Briefly, we introduce *α*_*z*_ to account for two effects, (i) the possibility that the publication of an obituary for one family decedent signals a greater likelihood of publication for another family decedent (including the limit of no publication at all), and (ii) that the choice of where obituaries are published make them detectable in our sample. Although there is no literature to draw on in choosing the values for these parameters, we combine what is known about who obituary authors are with some basic intuition [26, 27] to propose reasonable values (see Supplementary Discussion S2.3). One step is to simplify our approach by using only *z* = *{*spouse, non-spouse*}*, where the second element of the set means that the same *α*_*z*_ value is used for *z* = GN, AF, and ID. This acknowledges the practicalities that underlie obituary writing given family relationships. Most of the figures in the main text, with the exception of Fig. 2 which explores correlations broadly, show a representative pair of values (*α*_spouse_ = 0.75, *α*_non-spouse_ = 0.1) we consider conservatively plausible. However, we also discuss a wider exploration throughout, using two broad correlation scenarios, a low and a high. Both scenarios span the same range of *α*_spouse_ but distinct ranges of *α*_non-spouse_ (detailed values are shown in Supplementary Table S7). The representative values in the main text figures are part of the low correlation scenario. Qualitatively, the distinct scenarios do not change our main conclusions in any significant way. In Supplementary Discussion S2.3, we present a complete analysis of *α*_*z*_, argue in favor of the low correlation scenario as more likely to be a better estimate than the alternatives, show a comprehensive robustness analysis of our results for varying *α*_*z*_ (highlighting the low and high correlation scenarios in more detail), and present results for a third scenario for no correlations.

### 2.2 General patterns of local kin-connected deaths

Our first result is an estimation of *e*(*t*) between the start of 2020 and the end of 2022, shown in Fig. 1 (black curve). The light purple curve in Fig. 1 with the shaded area underneath shows the time series curve *e*_*o*_(*t*) of estimated COVID-19 deaths over the same cities the OB data cover spanning the time frame from March 2020 to August 2022 [25]; this is obtained from the EXS data set (see Methods). For the purposes of the figure, *e*_*o*_(*t*) is vertically scaled in order to be visually comparable to *e*(*t*) (however, in our analysis we use the true *e*_*o*_(*t*) values without any scale parameter). The first feature to notice about Fig. 1 is the period of time P_0_ stretching from the start of 2018 to the end of 2019, where *e*(*t*) is small in magnitude and oscillates around 0, supporting the validity of our method for obtaining *e*(*t*) as, during this period of time, there was no major deadly disease outbreak across the US.

**Fig. 1.**
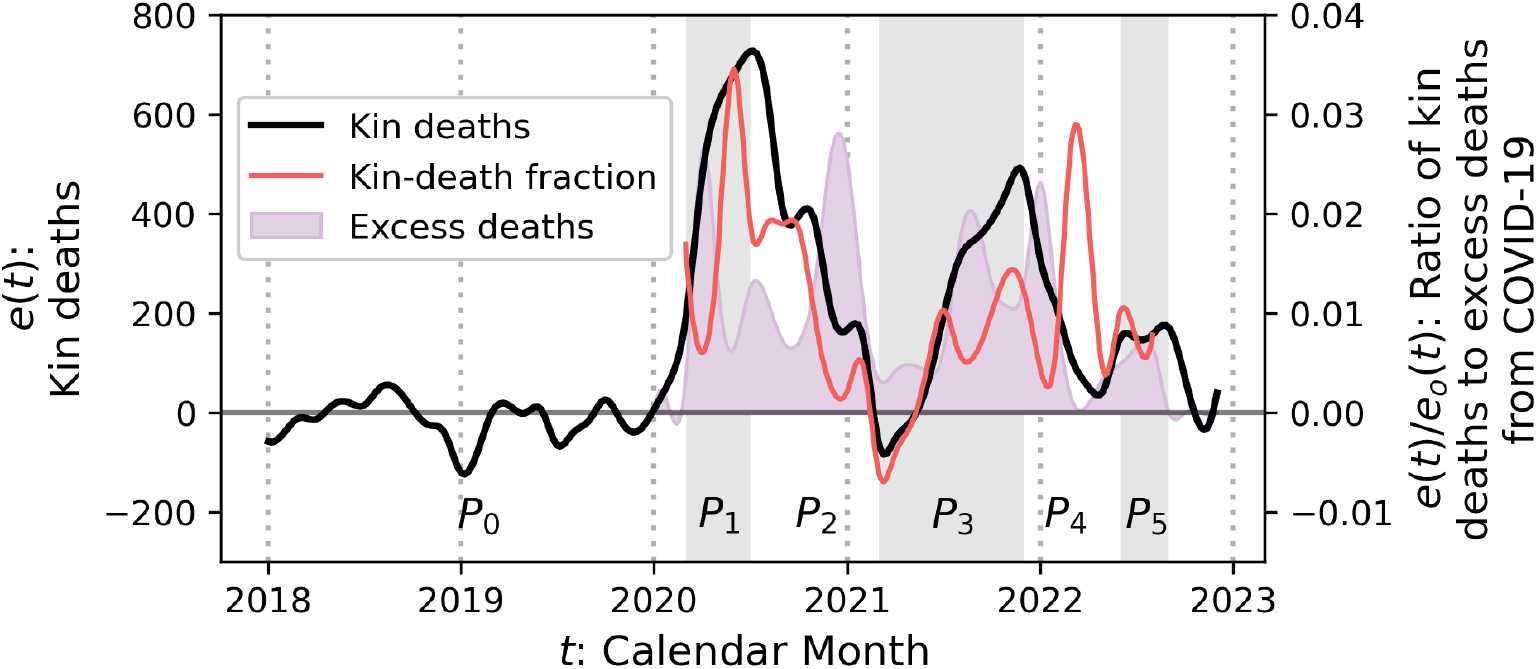
Estimation of the magnitude of aggregate local kin-connected deaths (LKDs) in the US. The black curve is *e*(*t*), the time series of LKDs aggregated over our large sample of US cities (left vertical axis). Several periods of growth and sustained kin-connected deaths can be identified in 2020 (P_1_), 2021 (P_3_), and 2022 (P_5_), as well as periods when *e*(*t*) decays and becomes quite small (P_2_ and P_4_), separating the periods of elevated *e*(*t*). The light purple curve with shaded area underneath corresponds to *e*_*o*_(*t*), the sum of estimated excess deaths attributable to COVID-19 from March 2020 to August 2022 across our sample of cities, created from [25]. The curve is reduced by a factor of 20, 000 in order to be visibly comparable to *e*(*t*) to be used as a qualitative guide of the temporal pattern of major COVID-19 surges affecting the US. The red curve corresponds to *e*(*t*)*/e*_*o*_(*t*), the time series of fractional aggregate contribution of LKDs with respect to excess COVID-19 deaths (right vertical axis). The fractional contribution peaks to ≈ 3.45% in June 2020, but has other temporal peaks later in the pandemic. We also highlight period P_0_, prior to the pandemic declaration, which shows only small deviations away from *e*(*t*) = 0, providing support for the validity of our method to detect overlaps above baseline.

After P_0_, *e*(*t*) displays a set of alternating periods of growth and decay (labeled as seen in Fig. 1). The general pattern is that growth occurs approximately during the springs for 2020 (February), 2021 (April), and 2022 (May), while decay takes place in the autumns of those years (September 2020, December 2021, and October 2022). Thus, it would be a good approximation to think of these periods of large *e*(*t*) as summer surges. Furthermore, it is noteworthy that, while the increases of *e*(*t*) in 2020 and 2021 occur alongside increases of *e*_*o*_(*t*), the temporal patterns these curves subsequently follow are distinct. This supports the hypothesis that the social processes driving LKDs operate partially independently of those driving the major COVID-19 surges. For example, the sustained aggregate LKDs of 2020 are consistent with a population that maintained contact with locally available extended family in ways that would not be equivalent to the overall combination of population social contacts (otherwise both *e*(*t*) and *e*_*o*_(*t*) would show matching temporal patterns). The pattern of LKDs in 2021, which peaks later in the year compared to 2020, may have been influenced by factors not present in 2020 such as the loosening of NPIs leading to family face-to-face reconnections and widening availability of vaccines during the early months of 2021 (see Sec. 2.4 and Supplementary Discussion S2.2 for additional considerations). In 2022, there is somewhat greater similarity between *e*(*t*) and *e*_*o*_(*t*), suggesting a return to pre-pandemic social patterns, although *e*(*t*) still remains elevated slightly longer than *e*_*o*_(*t*). We note that, although the description above refers to the low obituary writing correlation scenario, there is no qualitative difference in these results when using the high correlation scenario (see Supplementary Discussion S2.3).

The third curve in Fig. 1 (red) is the quotient of estimated kin-connected deaths to estimated excess COVID-19 deaths, *e*(*t*)*/e*_*o*_(*t*), which provides a time series estimate of the fractional contribution of kin-connected deaths to all COVID-19 deaths. This contribution reaches its maximum peak in June 2020, at which point [*e*(*t*)*/e*_*o*_(*t*)]*×*100% ≈ 3.45% based on the values of *α*_non-spouse_ and *α*_spouse_ stated above. To assess more broadly the impact of the correlations *α*_non-spouse_ and *α*_spouse_ to the peak contribution, in Fig. 2 we show the estimated maxima of *e*(*t*)*/e*_*o*_(*t*) in 2020 (June, panel A) and 2021 (November, panel B) as functions of those correlations. The maximum impact of kin is more sensitive to *α*_non-spouse_ than to *α*_spouse_, reflecting the underlying larger frequency of *z* = non-spouse among overlaps. For the June 2020 maximum, the majority of the area in the contour plot corresponds to percentages above 1%. The peak contribution reached in November 2021 is smaller than 2020 but constitutes only a small reduction in actual deaths (see *e*(*t*) in Fig. 1). The only parameter that contributes significant change in the estimate is *α*_non-spouse_, consistent with results below (Fig. 3) that show that spousal overlaps are rare in 2021. There is a significant peak in *e*(*t*)*/e*_*o*_(*t*) in early 2022, when COVID-19 excess deaths decay to low values but we estimate a significant portion of those to be kin-connected. It is pertinent to caution the reader that these percentages (2020 through 2022) are likely a considerable underestimate due to our own conservative methodology and the limitations of obituary data (see Discussion).

**Fig. 2.**
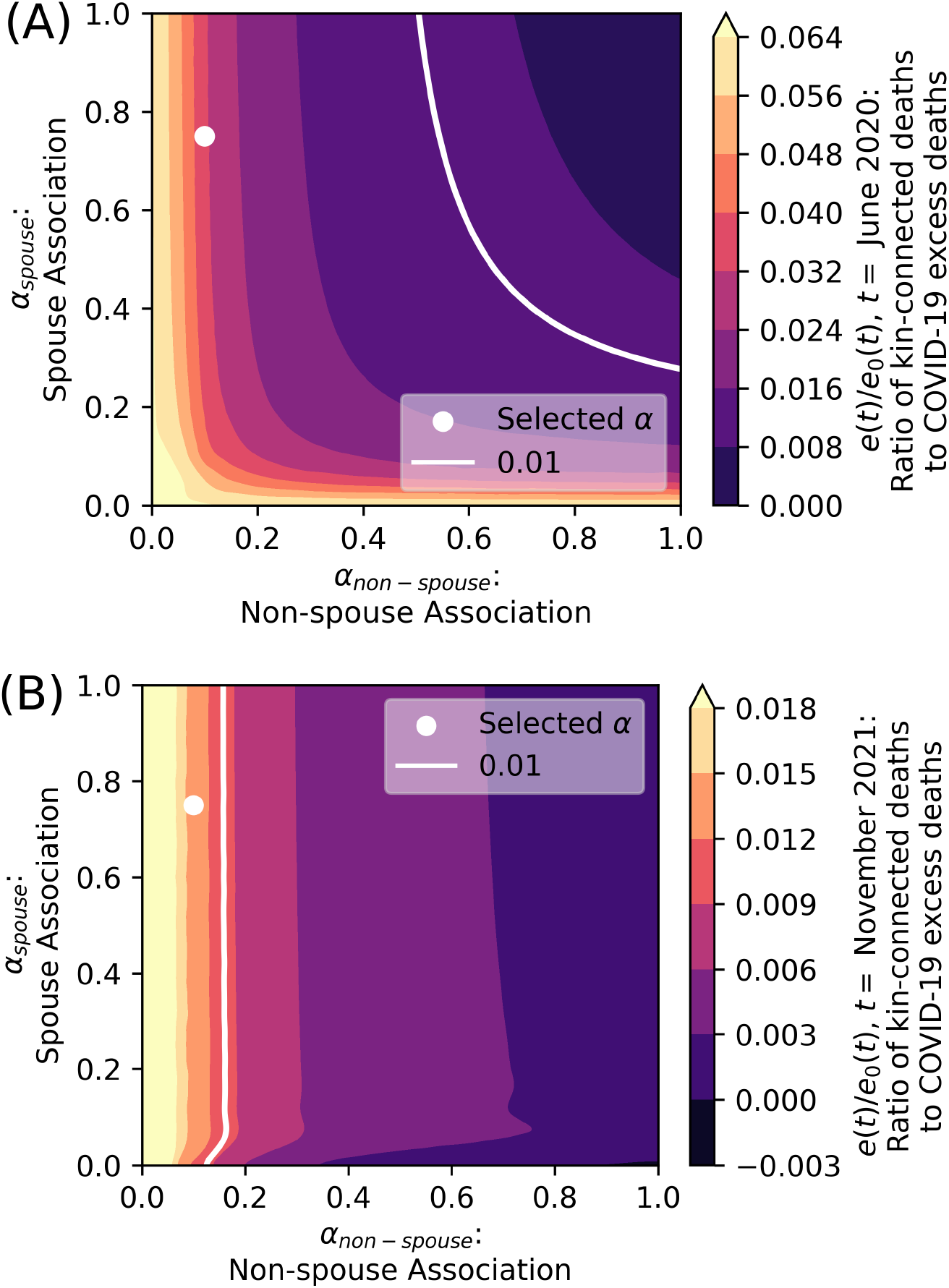
Effect of *α*_*z*_ parameters on yearly maxima of aggregate LKDs proportional contribution to contemporaneous deaths (2020 in panel A and 2021 in panel B). The contour plots show that the effect of *α*_non-spouse_ dominates changes, whereas *α*_spouse_ induces limited change, only becoming significant in 2020 for values near 0. Both plots show a white dot for choices of *α*_*z*_ we have used for the rest of the analysis in the main text. A referential line at 1% of kin-connected deaths for each year has also been added in order to gauge the range of parameter values of *α*_*z*_ that may lead to this or other percent estimations of mortality in the respective yearly proportion peaks.

**Fig. 3.**
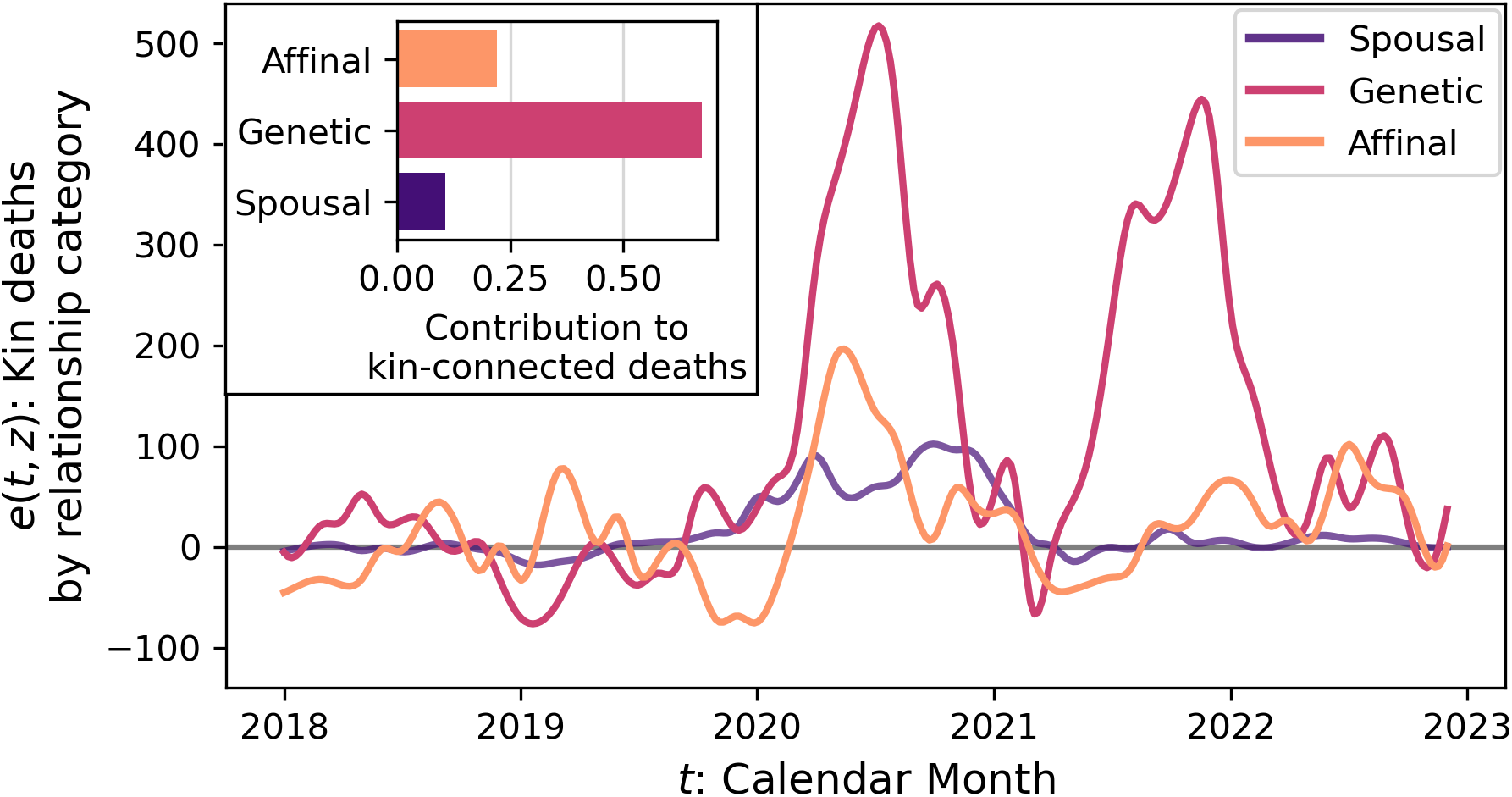
Estimation of the magnitude of aggregate LKDs by relationship category. Main plot shows *e*(*t, z*) by relationship categories: *z* = affinal (orange), *z* = genetic (magenta), and *z* = spousal (violet). At the start of the pandemic in 2020, genetic and affinal relationships begin a rapid growth that peaks earlier and at a lower level for affinal relationships in comparison to genetic relationships. The affinal relationship peak is slightly less than half as that of genetic relationships. In 2021, while *e*(*t, z* = AF) appears depressed and peaks around the holiday period, *e*(*t, z* = GN) exhibits a peak that resembles 2020 in width and height, although shifted to later than in 2020. Spousal relationships also show elevated behavior in 2020, but occurring later in the year than both affinal and genetic relations, indicating a different social process underpins this contagious contact. Inset shows the overall proportions of aggregate LKDs attributable to each *z*, where only *e >* 0 is counted.

Since the OB data set is not the product of a controlled sample, it is important to determine if our estimates of LKDs are affected by the number of COVID-19 deaths. We test this by checking location by location if the percentage of LKDs compared to COVID-19 excess deaths shows trends as a function of COVID-19 deaths. However, as we show in Supplementary Methods S1.7, no significant trend is detected, confirming that our estimate of LKDs is robust to the intensity of COVID-19 excess deaths.

### 2.3 Demographics of local kin-connected deaths

To further uncover the details of the social processes contributing to kin-connected deaths, in this section we analyze *e*(*t*) stratified by relationship categories (*z*) and by the genders of the first and second decedents in an overlap (respectively called here index and secondary deaths). There are several advantages to demographic analyses as they help assess different effects including the proportional contributions to LKDs coming from different relationship categories and any indications that gender roles might be relevant to LKDs. They also help in analyzing the extent of household versus non-household LKDs and kin-connected deaths more likely to contain both infectious and non-infectious effects (such as the widowhood effect), discussed in Sec. 2.4. We complete the section with a description of age statistics associated with overlaps.

To construct the *z*-specific time series *e*(*t, z*), we take all overlaps in each location *g* that generate *s*(*g, t*) and *r*(*g, t*) and, by selecting only those that match the desired *z*, respectively generate new series *s*(*g, t, z*) and *r*(*g, t, z*). We then apply Eq. 1 to these new series to obtain *e*(*t, z*). Although we only count overlaps of *z* in *e*(*t, z*), other relationship categories *z*′ ≠ *z* can have an effect as determined by the application of our method (see explanation leading to Eqs. 5 and 6).

In Fig. 3, we present the separate contributions to *e*(*t, z*) coming from spousal (violet), genetic (magenta), and affinal (orange) relationships using the correlations applied in Fig. 1. The results show that, from March of 2020 until August 2022 (inclusive), the largest proportion of overlaps involves genetic relations (e.g. parents, genetic siblings, children). Furthermore, the role of genetic kin is robust across correlation scenarios (see Supplementary Methods S2.3.5 for extended discussion). Defining the cumulative temporal contribution of each *z* as

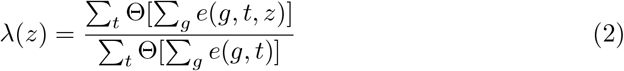

where Θ[*x*] = 1 when *x >* 0 and 0 otherwise, we find that genetic kin contribute between 61.0% and 71.2% of LKDs in the low correlation scenario. In the high correlation scenario, genetic kin still has the largest contribution to LKDs with a range between 46.2% and 55.5%. Spousal and affinal contributions are both smaller and which of them is second to genetic kin depends on the correlation scenario. For low correlation, affinal kin is typically the second contribution to LKDs, ranging from 19.9% to 22.9%, followed by spousal with 5.9% to 19.2%. In contrast, in the high correlation scenario spousal kin are the second contribution to LKDs, ranging from 26.0% to 38.4%, with affinal kin accounting for 15.4% to 18.5% of deaths. The particular case for the set of *α*_*z*_ used in Fig. 1 yields a contribution of 67.5% for genetic kin, 21.9% for affinal kin, and 10.6% for spousal kin (Fig. 3(inset)).

With regards to gender, a similar approach to the derivation of *e*(*t, z*) is used where, instead of selecting for *z*, overlaps are picked on the basis of ordered pairs *y* of the genders (*F* =female and *M* =male) of the index and secondary deaths. Along related lines, Eq. 2 is applied to define the contributions *λ*(*y*). This leads to *e*(*t, y*) shown in Fig. 4 for each type of *y* using the correlations from Fig. 1. As the inset shows, contributions *λ*(*y*) from all *y* are relevant, although there is an asymmetry slightly increasing female secondaries. From the temporal standpoint, in 2020 most pairs of genders contribute to *e*(*t, y*) simultaneously except for male-to-male overlaps, which displayed a delayed and smaller peak than for other gender pairs. However, in 2021 there was a considerable spike in female-to-female overlaps. Finally, in 2022, female-to-male overlaps where the largest contributors, although *e*(*t, y*) was at a more modest level (note that spousal relationships, as seen in Fig. 3, are not responsible for this increase). Performing a more comprehensive examination of the low and high correlation scenarios (presented in Supplementary Discussion S2.3.5) we find that, while in low correlation scenarios female-to-female pairs are typically the overall greater contributors to LKDs, in high correlation scenarios male-to-female and female-to-male contributions become more dominant; in all scenarios, male-to-male contributions are the smallest. Although the effect of *y* shows less differentiation than that of *z*, these results signal the presence of gendered roles in LKDs.

**Fig. 4.**
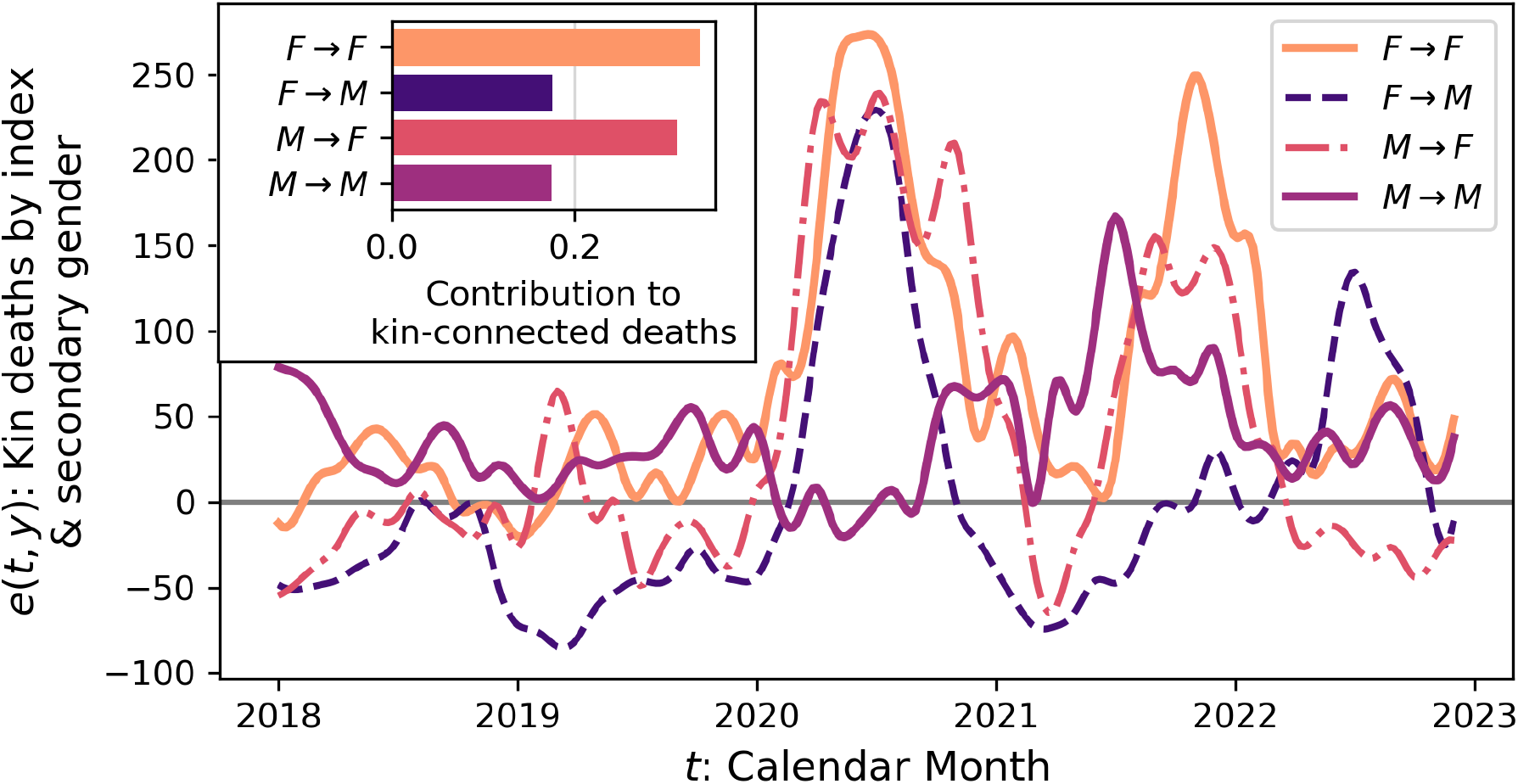
Estimation of the magnitude of aggregate LKDs by index and secondary genders. Main plot shows *e*(*t, y*) with *y* = *F* → *F* (orange), *y* = *F* → *M* (violet), *y* = *M* → *F* (magenta), and *y* = *M* → *M* (light purple). At the start of the pandemic in 2020, most gender pairs *y* show increased *e*(*t, y*) and peak simultaneously, remaining elevated for a considerable portion of that year, decaying before the holiday period. Male indexes show a smaller peak later in 2020, visible in both *M* → *F* and *M* → *M*. The series *e*(*t, y* = *M* → *M*) shows a different timing than other *y*, remaining elevated through the 2020 holidays. The pattern in 2021 shows more differentiation between gender pairs. Of greatest note is the curve *e*(*t, y* = *F* → *F*), which is by far the largest, peaking near the end of the year. Inset shows the overall proportions of aggregate LKDs attributable to each *y*, where only *e >* 0 is counted.

To close this section, we discuss the statistics of LKD ages. Combining together indexes and secondaries (see Fig. S2 and method details in Supplementary Methods S1.4.3), we find that the median age is 79 years. This does not indicate that LKDs only affect the elderly, as we measure contributions from ages starting at just over 40 years. However, LKDs may indeed be biased towards the elderly naturally under the effects of a contagious and deadly disease like COVID-19 due to the fact that its fatality rate increases rapidly with age [28] and because they are a joint event where the greatest probability of occurrence applies to the largest joint probabilities. Furthermore, since our data sample the elderly at a greater rate, they may be more sensitive in finding LKDs in those age groups. The last two points do suggest a counter point that COVID-19 cases rather than deaths could well be active among kin in younger age groups for which fatality and sampling rates are lower, hence making them less likely to appear as LKDs than those we do detect.

### 2.4 Studying confounders of LKDs among the elderly

Since we find that overlaps are frequent among the elderly, it is important to check if LKDs are affected by factors of particular relevance to this age group. For example, one could ask if the LKDs we detect can be a signal of elderly family members dying at similar times but not in a connected manner, or if elderly family members may live together and hence contribute to LKDs. While the first of these two effects (similar times of deaths by the elderly) are *included* in the baseline *r*(*g, t*) and thus *excluded* from *e*(*g, t*), as Eq. 1 requires, deaths among kin living together can legitimately be called LKDs. Therefore, we study three different confounders that could influence LKDs: (i) coresidence among kin members, (ii) deaths among nursing home or assisted care facility residents (which had large COVID-19 mortality rates in the US [29]), and (iii) overall confirmed COVID-19 deaths in elderly age groups. Except for coresidence, these confounders cannot be studied directly from our OB data as the variables necessary are not all present. Instead, we make comparisons between *e*(*t*) and the temporal signals of appropriately selected groups of COVID-19 deaths to argue that LKDs possess unique temporal patterns that cannot be explained by any of these confounders directly.

First, coresidence can readily lead to connected deaths because of the ease with which infections can be transmitted among members of a household. To determine if coresidence could in fact be an important contributor to LKDs, in Supplementary Discussion S2.1 we construct three separate calculations to estimate the probability of coresidence as a function of kin relationship *z* among the population aged 65 and older. The calculations rely on the Public Use Microdata Samples (PUMS) from the 2022 5-year sample of the American Community Survey. For *z* = spouse, the estimation is straightforward and yields a probability to live together of close to 98%. In contrast, based on the size and composition of households (spouses, relatives, and non-relatives that coreside) compared to the number of kin reported in obituaries, we estimate that the probability for *z* = non-spouse to coreside is close to 16% (this is likely a considerable overestimate based on current projections of living kin in the US [30]). Therefore, given that the percentage of overlaps among spouses in our data is small (see Supplementary Table S8) this supports the conclusion that LKDs are more typical among non-coresident kin. Figure 3 provides further evidence in this direction, as one can note that the spouse relationship temporal pattern is different than that of genetic kin which, in contrast, is basically the pattern of *e*(*t*). These observations do not support the explanation of LKDs from coresidence.

A second possible confounder among the elderly in the US during the critical stages of COVID-19 pandemic were the numerous deaths that took place within nursing homes. To determine what role, if any, nursing homes may have had on LKDs, in Fig. 5 (panel A) we present analysis for the year 2020 (when data were analyzed in considerable detail [29, 31]) that compares the pattern of COVID-19 deaths in nursing homes (orange) with the overall excess deaths *e*_*o*_(*t*) (light purple) and with *e*(*t*) (black). The results show a strong match between the temporal series of COVID-19 deaths in nursing homes and *e*_*o*_(*t*), but the pattern patently differs from that of *e*(*t*). The match between the first two time series suggests that measures to isolate nursing homes from the overall COVID-19 waves in the broad population were not particularly effective, with transmissions potentially driven by importation of infections through staff or possible external visitors. Furthermore, and crucially for our analyses, the difference between *e*(*t*) and COVID-19 nursing home deaths supports the interpretation that LKDs are a distinct process.

**Fig. 5.**
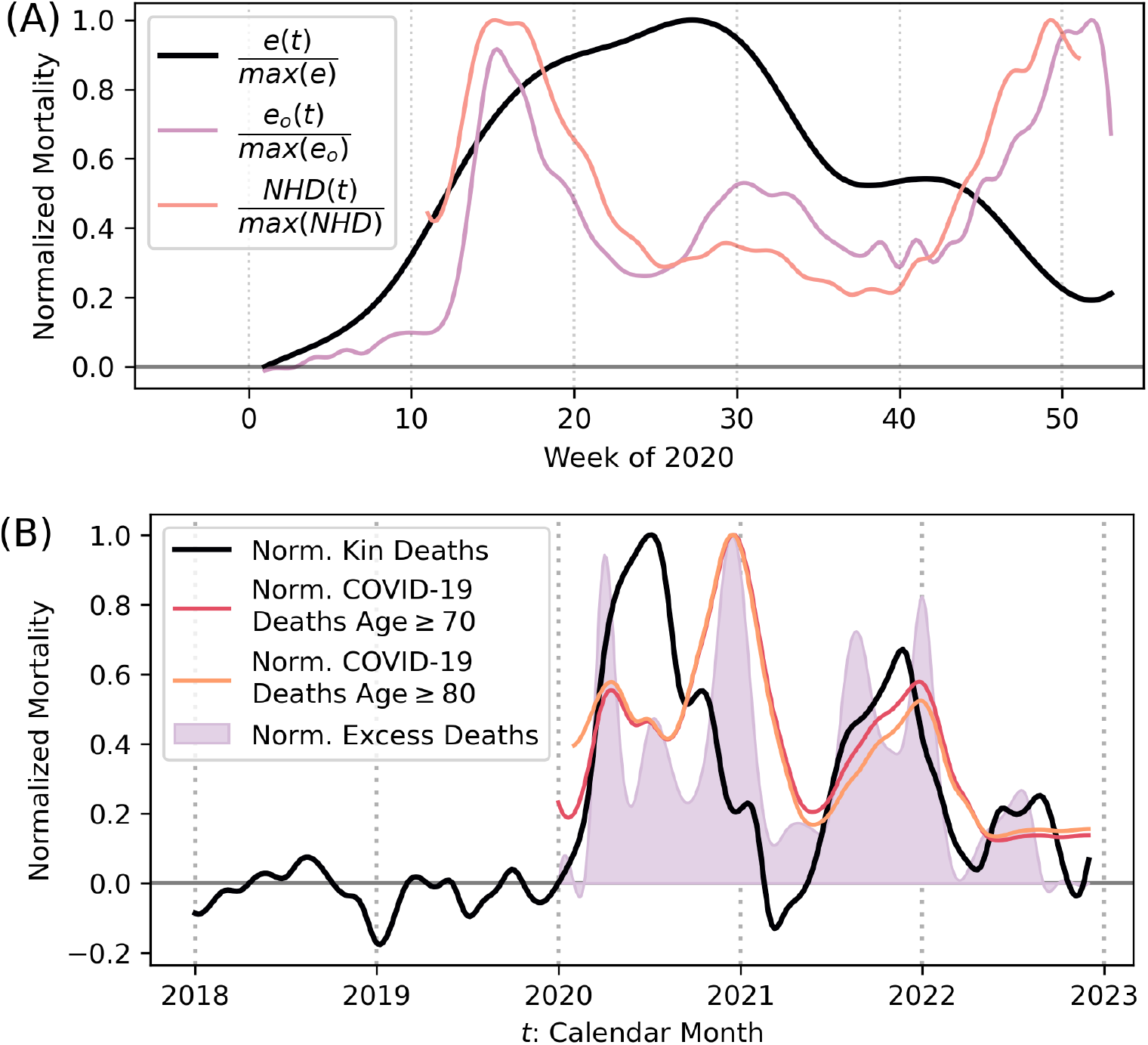
Comparisons between LKDs and other time series of possible confounders. All series are normalized by their maximum value. Panel A compares LKDs (black) with COVID-19 overall excess deaths (light purple) and nursing home COVID-19 deaths (orange) as defined in Ref. [29] in 2020. From the plot, it is clear that both excess deaths and nursing home deaths are similar to each other but differ from LKDs. Panel B compares LKDs (black) with COVID-19 overall excess deaths (light purple) as well as COVID-19 deaths among the elderly (in two age groups, 70 and above–red–and 80 and above–orange) as recorded in the MCOD data. The two age groups have virtually identical curves. In 2020, LKDs and elderly COVID-19 mortality are distinct in their temporal pattern but become more similar in 2021. This suggests that, while in 2020 LKDs and overall COVID-19 elderly mortality are not driven by the same mechanism, there appears to be greater similarity in 2021, possibly due to a relaxation of NPIs and a desire for people to reconnect socially within and outside families. It is interesting that in 2021 overall COVID-19 excess deaths still do not entirely align with LKDs and COVID-19 deaths among the elderly, suggesting differences that require further research to be clarified. For the time series above, smoothing is applied as explained in the Supplementary Methods S1.8, but *e*_*o*_(*t*) and nursing home deaths start from weekly data.

The third confounder, frailty, is common in elderly populations who were at particularly increased risk of death from COVID-19. This effect is partly taken into account when analyzing nursing homes but, because only a small fraction of the elderly reside in them (2.5% of 65 and over, 3.3% of 70 and over, 7.0% of 80 and over, from 2020 decennial census), we perform a more general comparison by extracting deaths coded as due to COVID-19 among individuals aged 70 and older and 80 and older from the comprehensive data set of deaths in the US provided by the Centers for Disease Control and Prevention in the US (MCOD data described in Methods). There are several interesting observations that emerge (see Fig. 5, panel B). First note that the two age ranges selected do not differ much and therefore the next discussion does not distinguish between them. Year by year, one can highlight interesting points. In 2020, elderly deaths from COVID-19 (red and orange) differ substantially from *e*(*t*) (black). Over the summer period, overall deaths among the elderly remained limited but increased as the end of the year approached along with *e*_*o*_(*t*) (light purple), in contrast to LKDs which were elevated over the summer and decreased substantially towards the end of the year. In 2021, there is greater similarity between overall deaths among the elderly and LKDs, both of which show a difference in trend with *e*_*o*_(*t*) in the latter half of the year, a pattern that would fit with families seeking to reconnect after a prolonged period of social distancing. These patterns support the interpretation that contact among kin differed from that of the elderly overall in 2020 but began to show more similarity from 2021 onwards with the loosening of NPIs.

From the results above, it appears unlikely that LKDs could be explained purely by coresidence effects, being a resident of a nursing home, or frailty of the elderly. In addition, other effects where the kin connection is relevant to joint deaths but does not involve infections such as the widowhood effect or accidental deaths appear insufficient to explain our results. First, the widowhood effect that typically affects a fraction of spouses [23], can only have a limited influence on LKDs due to the fact that most overlaps occur between non-spouses. Supplementing these considerations, we note that one connected hazard, traffic accidents, declined considerably in 2020 [32] and returned to slightly above pre-pandemic levels in 2021 and 2022 [33, 34], but the changes in counts of these fatalities are drastically smaller than the magnitude of LKDs we measure, excluding them as an explanation.

### 2.5 Discussion

In this work, we have presented evidence that deaths among local kin members, typically non-coresident, occur in excess of statistical expectations through various stretches of the COVID-19 pandemic during which household-to-household interaction was being intensely discouraged. This result is consistent with people maintaining active contact with local non-coresident family, which can lead to quietly sustaining population-level disease propagation. The kin-connected deaths we uncover through our novel approach have unique temporal features not shared with overall COVID-19 excess deaths, suggesting that if they represent an ongoing social process, they have a distinct temporal pattern to that of general social contacts. The temporal pattern of LKDs displays nuanced differences depending on the kin relationship type and gender, and affects people starting from an approximate age of 40. We have constructed our estimates in a conservative way, accepting the likelihood that we undercount the magnitude of LKDs. These results, together with the few other articles that identify whole or partial extended families as an infectious pathway in other contexts [20, 21, 35, 36], support the idea that kin infections and deaths are a broadly active and understudied phenomenon with appreciable consequences.

While at this stage one can only speculate about the social dynamics that could be compatible with the observed periods P_1_ through P_5_ of LKDs (see Fig 1), some clues can be obtained from the dates when each of the periods occurred (a more detailed account of the reasoning is provided in Supplementary Discussion 2.2). Each of the periods P_1_, P_3_, and P_5_ begins with the rapid increase of LKDs and their subsequent sustained elevated value for a number of consecutive months; periods P_2_ and P_4_ correspond to decaying LKDs. It is plausible the elevated periods, which consistently cover the summers, are partly connected to school year patterns (in addition to other modifiers that emerged during the pandemic). Summer months see more family interaction by way of child care [37, 38]. In 2020, this might have began earlier given the drastic reduction of child care through schools [18] or pre-schools [17] as part of NPIs. Indeed, empirical work has determined that extended family were the main non-coresident providers of child support during the closure of facilities in 2020 [39]. Period P_3_ in 2021 decayed later in the year, showed resemblance to overall COVID-19 cases among the elderly, and exhibited more LKDs among females, possibly reflecting social reconnection and effects such as the “kin keeper” role women are known to play in families [9]. In 2022, when most NPIs had been terminated and social contact had mostly returned to pre-pandemic patterns, it is not surprising that the dates of P_5_ are consistent with the yearly school break. By contrast, periods P_2_ and P_4_ take place roughly during children’s school year and do not show evidence that the typical holiday travel season is able to elevate LKDs, probably because travel is not a local effect.

Taking a broader perspective, the existence of non-negligible patterns of contact among kin would mean that the current epidemiological approach to community contacts may have shortcomings and that detailed consideration of such contacts could improve epidemic models. Focusing on kin community contacts, we first note that just as much as school calendars or holiday travel have particular times of the year when they occur and contribute significantly to disease propagation, our results suggest that interaction with local kin also has a yearly seasonality. Yet, most of the modeling and analysis of community contacts does not treat them (kin or other community contacts) seasonally. A second consideration is related to the evidence that kin are not uniformly distributed over the US but that, in fact, there are systematic trends by which it is more likely for people living in smaller population locations to be near extended family [10, 13]. Third, although not the direct subject of our study, one should also keep in mind that other types of community contacts like friends or neighbors can play the role of a regular contact for important activities such as childcare, although evidence shows that this occurs with a much smaller frequency than with extended family [39].

We should note that large agent-based models such as those found in [4, 40] are at a great advantage because they can be adjusted to replicate the kin and non-kin contact tendencies (seasonality and population effects) our work and the work of others indicate. This is because such models explicitly include household-to-household interactions, the kind of contact that best describes kin face-to-face interaction. Thus, such contacts could be given intensities that vary through the year, or the numbers of interacting households that are currently assumed in those models to be a fixed number across cities could be, for example, adjusted to reflect the trends in extended family availability as a function of city population [10, 13].

One other consequence of our study is worth highlighting around the possible medical and public health implications of genetic kin-connected infectious disease transmission. It is known that shared genetics can be an indicator for the correlation of severity of outcomes of infectious disease [41]. Therefore, kin transmission of illnesses may be accompanied by a dichotomy of family outcomes, some families suffering from little or no impact whereas others may see extended severe outcomes across the genetic kin group. These comments are consistent with the results shown in Fig. 3. This is an unexplored area of research at the population level which our work suggests should be considered.

The current study has a number of limitations, most of which suggest the magnitude of kin-connected effects is larger than estimated here. However, we first reiterate that our method does not directly track individual instances of disease contagion. Instead, it estimates deaths among local kin in excess of statistical expectation, similar in spirit to the concept of excess deaths. The details of actual chains of disease transmission among local kin cannot be clarified from our data as (i) non-fatal infections are not observable and (ii) obituaries are not comprehensively published so even fatal infections are not always captured. Yet, for our purposes the fact that excess deaths among local kin increase simultaneously with surges of COVID-19 in the population and then take their own course, strongly supports our interpretation that indeed local kin socialize under their own pattern, driving LKDs. With regards to our assessment that our method undercounts LKDs, we present the following arguments. First, there is likely an inverse relation between obituary publishing and financial means. This bias could lead to undercounts of LKDs by obscuring this effect among the more economically disadvantaged who, in addition, are more likely to require the aid of family in day-to-day activities [42–44]. Moreover, there is evidence that those at the lower socioeconomic strata where more affected during the pandemic [45], which could compound the lack of visibility of actual LKDs through our method. Second, obituary writing is idiosyncratic, illustrated by the fact that not every family member is mentioned explicitly but instead summarized in statements such as “many grand-children.” Also, some relationships or entire family branches can be completely omitted due to privacy or family estrangement. These practices can prevent LKDs from being detected through our approach. Although we do not have an exact estimate of the missing family, comparing with recent work in demographic projections of kin in the US, there are clear indications that obituaries typically only record a fraction of living family (see Ref. [30]). Third, our method is designed to avoid false positive overlaps and overestimation (see Methods and Supplementary Methods S1.2.5), leading to a high rate of false negatives. This means, in practice, that we accept lower recall with as much as 19% of signal remaining unused. Fourth, obituary sampling increases with decedent age, which means that fatalities among younger age groups (including those due to kin), are harder to detect (see Supplementary Methods S1.4.2). Fifth, the absence of research on the patterns of obituary writing limit our ability to make improvements in several aspects of our study, such as a more nuanced probabilistic model of obituary sampling or more demographic detail in our analysis.

In summary, we believe that the addition of local kin to the list of identified relevant contacts for respiratory contagious diseases may constitute an important advancement for epidemiology. The patterns we identify can readily be included into current epidemiological theories and models by updating household-to-household interactions to replicate contact with local kin. However, in order to achieve the best possible results, we believe it is necessary to undertake the collection of specific data to overcome the limitations of the non-probability sample we use, enabling better calibration of the magnitude of the effect. Such data could also address the question of the difference between LKDs among genetic or affinal kin, the effects of having local kin available on local infectious disease propagation and fatalities, and how to improve NPIs to include possible local kin effects.

Regarding this last point, the development of updated NPIs to combat future large outbreaks, if distancing with kin proves impossible, the US could benefit from adopting in a systematic way the formation of “support bubbles” with key extended family members. This strategy, while receiving some informal attention in the US during the COVID-19 pandemic [46], was in fact adopted as policy for NPIs in various countries such as New Zealand and the UK [47]. This strategy appears to be fairly effective [48].

## 3 Methods

### 3.1 Data and spatial aggregation

This article uses five sources of data. The first is a large database of obituaries (OB) constructed from the web. The second is the Multiple Cause of Death (MCOD) restricted-use vital statistics data compiled by the National Center for Health Statistics of the Centers of Disease Control and Prevention (CDC), which records geographically-disaggregated monthly all-causes mortality in the US [49]. The third is the database created in Ref. [25] estimating county-level excess deaths associated with COVID-19 across the US (EXS). The fourth data set is the American Community Survey (ACS) from the US Census (see multiple sections in Supplementary Methods for details). The fifth is a freely available crosswalk to map between geographic resolutions [50]. The first two data sets cover the period between January 2018 to December 2022, whereas the third includes from March 2020 to August 2022. Each analysis is performed with the pertinent time frame based on the data, and explained in the relevant sections.

The OB data set is generated from individuals publicly posting the deaths of *decedents* in newspapers and/or funeral home websites across the US. We collect obituaries from an intermediate source (Legacy.com) that centralizes the original obituary texts. Obituaries are published with reference to cities (based on the serving newspaper’s or funeral home’s location) and on specific dates. *Typical* obituaries in the US contain information such as the date of death (*θ*_*i*_ for decedent *i*), information allowing age inference (which we convert to age *a*_*i*_ at death), and the decedent’s family such as parents, spouses or former spouses, siblings, aunts and uncles, children, step-children, grandchildren, great-grandchildren, children in-law, in-law siblings, to name the most common, all with their specific names. Very elderly or very young decedents can sometimes report descendant or ascendant family further removed (e.g. great-great-grandchildren or grandparents). However, obituaries are not comprehensive, not always mentioning all relatives. For instance, some relatives are completely omitted and others mentioned in generic ways, e.g. “many grandchildren.” Commonly, obituaries make references to a decedent’s life and, in doing so, the descriptions offer pronouns from which decedent’s *i* gender, *x*_*i*_, is inferred. Decedent’s *i* city *g*_*i*_ is recorded at the time of web scraping which is done as a location-based query. The date of death *θ*_*i*_ is recorded as *i* is scraped. The month and year of death *t* = *t*_*i*_ is the one that includes the date *θ*_*i*_. From these data, we count the number of obituaries 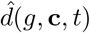 found in location *g*, month *t*, for people in the age and gender bin **c** (see Sec. 3.4.1 for bin definitions). Finally, name and relationship information for the decedent is found through the process described in Sec. 3.2, and recorded in an ordered set **o**(*i*) of *f*_*i*_ pairs of names and relations, where *f*_*i*_ constitutes the number of unique people found in obituary *i*, including decedent *i*.

Cities in the OB data set approximately correspond to core-base statistical areas (CBSAs). Thus, the next two data sets, coded to Federal Information Processing Standard (FIPS) corresponding to US counties, are processed to provide CBSA-level values. This is done by using the crosswalk downloadable from the National Bureau of Economic Research [50].

The MCOD data provide anonymized individual records of each death, the calendar month when it occurred, decedent’s age, gender, cause of death using the International Classification of Diseases version 10 codes (ICD-10), and location using FIPS codes [49]. After mapping FIPS to CBSAs, we generate numbers of deaths *d*(*g*, **c**, *t*) per location *g* (a CBSA), month *t*, and age and gender **c** consistent with those defined for OB.

The EXS data set contains the monthly time series of excess deaths attributable to COVID-19 for every US county (FIPS), which include deaths directly coded as COVID-19 as well as other deaths that are likely attributable to COVID-19 [25]. Aggregating to CBSAs, we generate series *e*_*o*_(*g, t*) for each of the cities covered by our OB data. We also further aggregate this information over time to cities (*e*_*o*_(*g*)) and over cities for each month (*e*_*o*_(*t*)).

### 3.2 Data Treatment to generate OB database

To determine information from obituaries, we apply the methods described next. The result of these methods is the creation of 1, 406, 321 obituaries over 882 CBSAs with a high likelihood of being unique (one per distinct decedent) and containing the information that enables our analysis. Obituaries where needed information cannot be determined (dates, age, gender, or family members) are discarded. The retained obituaries contain a total of 18, 244, 177 family relationships. However, additional rules (especially those stated in Sec. 3.4.1) to analyze the best sampled cities reduce this further; the final numbers are stated in Sec. 3.4.1.

#### 3.2.1 Initial cleaning

First, we remove from each raw obituary scraped uninformative HTML code not related to the decedent such as hyperlinks to ads. The result of this are 1, 874, 766 ASCII-format obituary documents. To this document corpus, we apply two data engineering pipelines that run in parallel, information extraction and de-duplication, explained in the next two sections. Further details are provided in the Supplementary Methods S.1.1 to S.1.3.

#### 3.2.2 Relationship and demographic information extraction

To extract relationship and other information from the resulting documents, we combine a large language model (LLM) and regular expression methods. The LLM allows us to determine family members mentioned in each obituary reliably and at scale when an appropriate prompt is used for this purpose. We submit prompts that include obituary text and a decedent’s name via Google’s API to the text-bison@001 model, instructing the model to perform named entity resolution for each family member of the decedent and relationship extraction for the relationship of the family member to the decedent (model prompt is specified in the Supplementary Methods S1.2.3). We require the LLM to generate a formatted output record for each name and associated relationship but, when this fails, the particular record is discarded.

We apply two controls against possible hallucinations by the model. First, for any output of the model to be accepted into our obituary data set, the names and relationships that the model produces are checked against the input text to verify that the output does not contain hallucinated content. Second, all obituaries are submitted at least twice to the model. If the two outputs match entirely (all names and relationships), the names and relationships are accepted. For those obituaries with persistent discrepancies after two applications of the model, we execute up to two additional runs and use a majority rule, along with confirmation of the output text, to accept the output. The majority rule checks if, out of the model outputs, two of them match entirely and, if they do, the matching information is used for the obituary. If after the fourth run there are unresolved discrepancies between model outputs for an obituary, we construct a new set of family members and relationships for the obituary that still requires model consensus between executions. To do this, we check names and relationships individually; if a named family member and the corresponding relationship appear in more than one result, we accept the family member as accurate, otherwise we remove the family member; the check to determine if the accepted name and relation can be found in the obituary is still applied.

Individual names and relationship terms are standardized. First, names are restricted to a first name, a blank separating space, and a surname. Prefixes (e.g. “Dr.”), suffixes (e.g. “Jr.”), middle names, and names that match a type of family relationship (e.g. “John Cousins”) are discarded as they may lead to false positive overlaps. Because this step can lead to duplicate names (e.g. father followed by “Sr.” and son by “Jr.”), only the first occurring instance of the repeated name is kept. Second, the LLM copies relationship terms used in the obituary. Thus, grandchild may appear in many forms such as “grandkid” or “grandbaby.” We create a table of equivalencies among relationship terms and, for each distinct relationship, pick a term to which all other equivalent terms are standardized (see data repository [51]). One record is created for the decedent, with relationship “self.” To capture the results of the process, we refer to the *n*-th name in *i*’s obituary as **o**_*n*,1_(*i*) and respective relationship as **o**_*n*,2_(*i*); these two variables effectively define (**o**_*n*,1_(*i*), **o**_*n*,2_(*i*)), the *n*-th element of **o**(*i*).

Gender *x*_*i*_ and age *a*_*i*_ for decedent *i* are determined by regular expression methods. Gender is found by counting the frequency of male and female pronouns used in the obituary. When the number of these is equal, the obituary is discarded. Age is determined by extracting candidate age information from each obituary document and then applying preference rules to definitively assign the age. The candidate age information in each obituary can be of three forms: based on birth and death years from HTML code, based on stated ages in the obituary text, or based on dates stated in the text. If more than one candidate age is possible from this information, we compare them and apply preference rules by which to pick *a*_*i*_ in case they are not consistent with each other (see Supplementary Methods S1.2.3). If no age information is found with a positive value less than or equal to 120, the obituary is discarded.

We annotate 386 obituaries with 5, 002 family members manually to validate our results, identifying each family member and their relationship to the decedent for each obituary. From this annotated dataset we calculate recall of 0.8065, precision of 0.9963, and an F1-score of 0.8914. For details on model performance and data treatment, see Supplementary Methods S1.2.5. It should be noted that because of the choices involved in several of the steps of the process above, including which names are eliminated (e.g. surnames that match a family relation), unknown biases of the LLM, and the de-duplication steps outline in Sec. 3.2.3, it is possible that changes in those steps could lead to small variations of our detailed results. Although we have not performed a fully systematic analysis of these possible variations, we have not found qualitatively or quantitatively relevant differences during extensive testing of our method.

#### 3.2.3 Deduplication

The initial set of HTML documents are de-duplicated to unique URLs, as exact replications occasionally arise from inconsistencies in the order of search results over the time in which we sampled the records. This straightforward de-duplication reduces the original 1, 874, 766 obituaries to 1, 630, 258. Then, the execution of the steps the LLM and other other data extraction (Sec. 3.2.2) further reduce the data to those obituaries for which relevant information for our analysis can be extracted, including decedents’ age, gender, and family members.

The application of the process of overlap detection described in Sec. 3.3 uncovers additional duplication in the form or *near-duplicate* obituaries, i.e. non-identical obituaries for the same person. Near-duplicates are typically the result of real-world processes such as an obituary submitted to multiple newspapers or submitted to multiple online resources that lead to slightly different posted documents. The overlaps generated by near-duplicates are fictitious and create problems in our calculation of obituary weights (Sec. 3.4.1) and estimation of meaningful overlaps (Sec. 3.5).

Near-duplicate obituaries are systematically eliminated as follows. We begin with the set of all pairs of obituaries found to form overlaps within all cities in the data, using Δ*θ* = 5 years and *m*_*o*_ = 1 which provides the most comprehensive set of overlaps and, consequently, possible near-duplicates. Then, we define a set of scores that measure obituary similarities (an indication of possible near-duplication) across all overlapping obituary pairs based on names of decedents, similarity of dates of death, similarity in the names and relationships with family members, and what fraction of family members mentioned in both obituaries take part in the overlap. Next, we perform three steps to use the machine learning classification model XGBoost to aid in near-duplicate search [52]. First, we extract a sample of the overlapping pairs. The sample is created in a sorted way, starting with obituary pairs that score high in the characteristics that signal near-duplicates. Obituary pairs in the sample are manually labeled duplicates or pairs of distinct individuals. This approach over-samples the near-duplicate class (the rarer of the two), reducing sample class imbalance effects on the training of the classification model. The sample consists of 953 overlapping pairs, 55.7% of which are near-duplicates. Second, the model is first trained with the sample and subsequently run over all obituary overlaps to classify them as candidate duplicates or not. In addition, XGBoost provides a probability (which is the information it uses to classify the pairs), and we use this probability to help in the final determination of duplication. Third, the resulting obituary pairs labeled as duplicates by XGBoost are treated as candidate duplicates over which we make a final determination. We first check obituary pairs in case most or all of their key information matches (decedent name, age, gender, month of death, and family members and relations) and, if it does, automatically confirm the pair as duplicates. The remaining candidate pairs are all inspected visually. When a near-duplicate is found, we eliminate all but one obituary among the duplicates, leaving the one with most relationships (or one at random if the number of relationships is the same). See the full description of de-duplication in Supplementary Methods S1.3. We reach accuracy and F1-score of approximately 0.92 in finding duplicate obituaries. Once near-duplicates are excluded, we obtain a sample of 1, 406, 321 obituaries of distinct decedents which represent 9.77% of the 14, 389, 763 all-causes deaths reported in MCOD in the period 2018 to 2022. These obituaries undergo the rest of the analysis described below.

### 3.3 Overlap identification

To identify an overlap between index obituary *i* and secondary obituary *j*, as well as to find the overlaps that have *i* as their index, we use the concepts and criteria now described.

Let us use **o**_·,1_(*i*) to refer to the set of first components of **o**(*i*), i.e. only the names in obituary *i*. The number of shared names found in obituaries *i* and *j* is given by

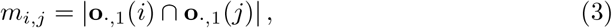

which is the cardinality of the intersection between **o**_·,1_(*i*) and **o**_·,1_(*j*). Next, if the date *θ*_*i*_ of obituary *i* is in month *t* (which from now we state as *θ*_*i*_ *∈ t*), we define 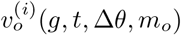, the set of obituaries that overlap with *i* in location *g*, month *t*, and satisfying the tolerance parameters Δ*θ* and *m*_*o*_. A node *j* belongs to this set if

1. *m*_*i,j*_ ≥ *m*_*o*_, where *m*_*o*_ is an integer ≥ 1,
2. 0 *< θ*_*j*_ − *θ*_*i*_ *≤* Δ*θ*, and
3. when *j* also satisfies conditions 1 and 2 with any other obituary *h*, but *θ*_*h*_ *> θ*_*i*_.

However, if *θ*_*h*_ = *θ*_*i*_, *j* was first found during processing of obituary *i*.

The third condition guarantees that *j* can only be secondary of a single index obituary (here *i*), thereby ensuring that any set of connected overlaps are a tree, consistent with typical epidemiological assumptions regarding infection spread. In all the overlaps between *i* and the obituaries in the set 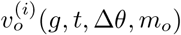, *i* is the index death and each of the elements of the set are secondary deaths to *i*.

### 3.4 Estimation framework for local kin-connected deaths. Weights

To estimate population level LKDs based on observed overlaps in our data, we have to construct an appropriate weighing scheme. Weights estimate how many events (obituaries or overlaps) a single observed event in our analysis is meant to represent in the population. We now explain our approach.

Each *overlap weight* (generically symbolized by *u*) is estimated based on a probabilistic model of obituary sampling. Let us describe the intuition. Imagine an ideal data set of “perfect obituaries” where all local deaths are captured. Applying the rules in Sec. 3.3 with given *m*_*o*_, Δ*θ*, and *t*, we would find all applicable LKDs. We introduce the vector **x** to record the features of all individual deaths and the pairs they form, including dates of deaths, location, each decedent’s demographics, and specific relationships among pairs of decedents. In contrast, in the data we gather each instance of paired deaths is captured by an obituary overlap with a sampling probability defined as follows: if there are *p*_**x**_ *paired deaths* in the population characterized by the feature vector **x**, 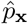 of which are found in our data, their sampling probability is given by 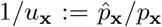. Then, the total number of paired deaths satisfies 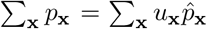 where the sum is over all possible **x** of the population represented by the data. In practice, however, *p*_**x**_ is not known and therefore, to calculate these sums, we must postulate a reasonable model for the probability 1*/u*_**x**_ given **x**. With this model in hand, we can estimate paired deaths from 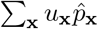.

We estimate the probability 1*/u*_**x**_ as the product of two contributions: the probability that the index death is sampled multiplied by the probability that, given the sampling of the index death, the secondary is also sampled. Each of these probabilities relies on two others: the probability that a death at a specific month, location, and demographic group is sampled, and the probability that if the index and secondary deaths are related by relationship *z*, that the secondary is sampled. The detailed arguments are next.

#### 3.4.1 Obituary sampling and obituary weights

The probability to sample an isolated obituary *i*, independent of possible shared family tendencies, is given by the ratio of the number of obituaries 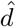 to deaths *d* in the same location *g*_*i*_, month *t*_*i*_, and with the same personal characteristics of age and gender **c**_*i*_, or

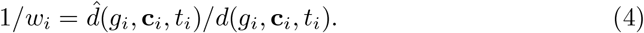

These probabilities are determined from the OB and MCOD data sets. All obituaries in the same location *g*, month *t*, and for people with shared personal characteristics **c** have the same weight *w*(*g*, **c**, *t*). The age and gender vector **c** is binned to two genders and three age ranges in years, [0, 70), [70, 80), and ≥ 80 to obtain sample sizes that facilitate our analysis (see Supplementary Methods S1.4.1).

Two different limitations can produce *w*(*g*, **c**, *t*) values that lead to problems in our analysis: low numbers of obituaries for a combination *g*, **c**, and *t*, and the possibility that there is geographic mismatch between the city reported in an obituary (usually the location of the nearest or preferable newspaper or funeral home) and the city of death of the decedent. The first effect can lead to poor estimation of the obituary weight. To manage this, we only use combinations of *g*, **c**, and *t* where 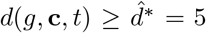, a threshold value obtained from robustness checks of the results (see Supplementary Methods S1.5.2). In case an overlap involves an obituary in one of the excluded combinations of *g*, **c**, and *t*, the overlap is not counted.

The second effect typically occurs around small-population cities where the local newspapers or funeral homes can be located outside one or more such small cities in order to serve a wider market. In this case, one can find that the *g* of the obituary source (denoted *g*_*s*_ in this argument) and of the city(ies) covered (denote by *g*_*c*_ here) are such that *d*(*g*_*s*_, **c**, *t*) *≪ d*(*g*_*c*_, **c**, *t*). On the other hand, because the obituary source is reporting deaths from both *g*_*s*_ and one or multiple *g*_*c*_ (comprising a sum of deaths such as 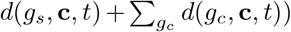, this can lead to 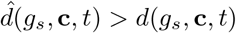. We are generally not able to determine this geographic mismatch in a direct way from information on obituary text or their source (e.g. newspaper). Therefore, to avoid such cases, we eliminate any *g* where *any* combination of *g*, **c** and *t* is such that 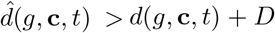, where *D* is a parameter. The case of *D* = 0 is presented in the main text, but other values are shown in the Supplementary Methods. S1.5.1.

After the application of these two sampling rules, the number of cities that remain in the data are 276 (listed in our data repository [51]) and the number of obituaries in the surviving combinations of *g*, **c**, and *t* in those cities is 824, 576. To provide a sense for how well this sample captures those cities, our obituaries constitute 7.13% of those cities’ 11, 559, 468 all causes deaths, as reported in MCOD. On the other hand, the cities that remain in the sample had a population of 257, 553, 104 according to the 2020 ACS 5-year estimate, which represents 77.7% of the total US population found in the 2020 decennial Census (331, 449, 281).

#### 3.4.2 Overlap weights

To calculate the weight *u*_*i,j*_ of two overlapping obituaries *i* and *j*, our logic is as follows. Let us call *q*_*i*_ the probability to sample index obituary *i*. Given the correlation parameter 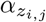, where *z*_*i,j*_ represents the relationship between *i* and *j*, a secondary obituary *j* overlapping with index *i* is published due to family correlations with joint probability 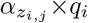. However, there is also a conditional probability 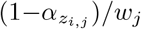 that, given the publication of *i*, such a secondary obituary is not published due to family correlation but, instead, purely based on obituary sampling given by Eq. 4, leading to a joint probability 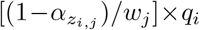. This means that in an overlap of obituaries *i* and *j*, the secondary *j* is published with probability 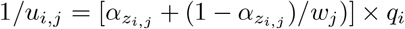. Finally, if *i* is an obituary that is not a secondary to some other earlier obituary, then *q*_*i*_ is equal to 1*/w*_*i*_ from Eq. 4, which contains all statistical information we have about *i*; otherwise, if it is secondary to an obituary, say *h, q*_*i*_ is equal to the probability to reach *i* from *h*, which is 1*/u*_*h,i*_. In summary, the weight *u*_*i,j*_ of an overlap satisfies

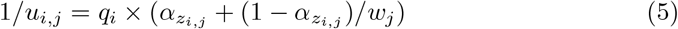

where,

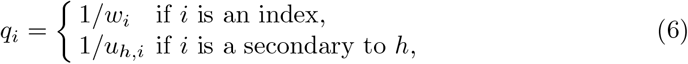

It should be understood that the parameters *g, t*, Δ*θ*, and *m*_*o*_, as well as the *α*_*z*_s of all overlaps relevant to the *i* and *j* overlap, are implicitly involved in Eqs. 5 and 6. To obtain the final value of *u*_*i,j*_, Eqs. 5 and 6 need to be applied recursively until an obituary is reached that is not a secondary in any overlap.

### 3.5 Local kin-connected deaths (LKDs)

To write the correct expression for the time series of observed overlaps, *s*(*g, t*), we use the discrete Heaviside function Θ(*k* − *b*) for two positive integers *k* and *b*, equal to 1 if *k* ≥ *b* and 0 otherwise. Thus, first applying Eqs. 5 and 6 without the restriction on 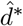 to obtain *u*_*i,j*_, *s*(*g, t*) is given by

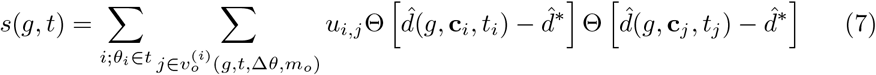

Equation 7 is also applied to generate each random realization *r*_*μ*_(*g, t*), *μ* = 1, …, *M*, of overlaps used to create the baseline *r*(*g, t*) for continuing overlaps, but this is done after the obituary dates *θ* have been randomly shuffled under some conditions (explained next) through the entire time window from January 2018 to December 2022 within each location *g*. No obituaries are duplicated or lost during each realization, only reordered if possible. The condition we apply is that, if in the real timeline of obituaries in *g*, obituary *i* occupies time *θ*_*i*_, another randomly chosen obituary *h* can take the place of *i* if and only if **c**_*h*_ = **c**_*i*_ and *f*_*h*_ = *f*_*i*_. However, regardless of what obituary is randomly assigned to time *θ*_*i*_, in applying Eqs. 5 and 6 to determine the weights of any potential overlaps, the weights used for the obituaries are those of the observed time series; in other words, the weights are pinned to the observed timeline and do not travel with the obituaries.

The time-shuffling rules just described account for a number of statistical patterns that may affect continuing overlaps. First, such overlaps are more likely to occur during periods of high mortality because of a likely increase in numbers of obituaries. Therefore, preserving equivalent deaths (of equal **c**) at each time point achieves this effect. Second, obituaries that mention large numbers of people are more likely to randomly overlap. This leads to the rule requiring that obituaries can only occupy a time where another obituary had equal *f*. And third, obituary reporting and consequently overlap frequency may differ by location due to local obituary affordability, availability of outlets (local newspapers), local demographics (e.g. age and gender), and local frequency of particular family names (e.g. as would occur if a location was characterized by certain historic and/or immigrant group). This justifies the rule of shuffling obituaries only within the same location *g*. Our approach has the two effects of, first, breaking any legitimate temporal connection between obituaries while, second, leaving a sample of correctly-weighted alternative obituaries that preserve the temporal sequence of mortality of given features, locations, and family sizes.

The overall baseline of continuing overlaps for *g* is given by the average

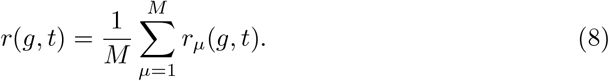

Equations 7 and 8 are plugged into Eq. 1 to generate aggregate LKDs, where any *g* that violates the *D* condition stated in Sec. 3.4.1 is discarded.

### 3.6 Determination of relationships in indirect overlaps

Obituaries can overlap *directly* or *indirectly*. In the first (direct) case, the name of one decedent can be found in the obituary of the other decedent, or both decedents are named in each other’s obituaries (along with at least *m*_*o*_ − 1 additional common names). In the second (indirect) case, neither *i* nor *j* are mentioned in each other‘s obituaries, but there are at least *m*_*o*_ names mentioned in *i* that are also mentioned in *j*. The difference between these two situations require that the inference of relationships between decedents be done in different ways.

When overlaps are direct, the relationship is explicitly known in **o** of either decedent. For example, for an overlap between *i* and *j*, if *n* corresponds to **o**_*n*,1_(*i*) = *j*, then **o**_*n*,2_(*i*) specifies the relation between *i* and *j* and from that, we can determine the relationship category *z* (genetic, affinal, spousal, or indiscernible). In contrast, to determine what type of specific relationship the two decedents *i* and *j* have from indirectly overlapping obituaries, we need to employ a logical procedure. Furthermore, although it is generally not possible to determine the specific relationship in indirect overlaps because obituaries do not provide genealogies, relationship categories are typically inferable.

By definition, there are *m*_*i,j*_ intermediary names in the indirect overlap. For each intermediary *h* we determine the two relationships *z*_*i,h*_ and *z*_*j,h*_. From this pair of relations, we infer the nature of the relationship category between *i* and *j through h*. Let us call this 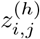. This determination is done based on a table of relationships constructed by the authors and provided in the data repository [51]. The logic is straightforward and based on knowledge of family relations. To illustrate how the table is constructed, consider the case of *i* being the parent of *h* and *h* being a sister-in-law to *j*, then the relationship between *i* and *j* through *h* would be recorded as affinal 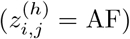.

Because the relationship category one can infer between *i* and *j* depends on the intermediary *h*, once all the *m*_*i,j*_ relationship categories are determined based on the relationship table, we must compare them against each other to make our best inference of the relationship category between *i* and *j*. This is done based on an order of precedence among such relations. Concretely, the set 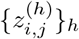 of *m*_*ij*_ relations determined through *h* are such that if one relation 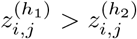, then from these one would conclude that 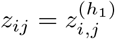. There are two mutually exclusive orders of precedence we use to perform such comparisons, distinguished by whether “spousal” is present or not in the set of relationships: “genetic” *>* “affinal” *>* “indiscernible” and “spousal” *>* “affinal” *>* “indiscernible”. The exclusion comes from the assumption (not necessarily always true), that spouses are typically affinal relations (blood marriages are highly discouraged and illegal if people are too close genealogically). In addition, we do not treat a spousal relationship as affinal because the behaviors among spouses are different than with other affine relations (e.g. spouses typically live together).

We note that indirect overlaps can emerge for a number of reasons. They may reflect kinship distance or estrangement. However, they may also occur because of our rules for constructing the ordered set **o** for each obituary. As explained in Supplementary Methods S1.2.3, because the presence of names such as “John Doe” and “John Doe Jr.” in the same obituary may lead to subsequent misidentification of people, we require that one of these names and associated relationship be eliminated from **o**. Such eliminations may induce two obituaries that could overlap directly to, instead, overlap indirectly. To evaluate the possibility of errors, we visually inspect 100 indirect overlaps and find 2 errors in the relationships predicted by this method. This represents a small error rate of 2%, not large enough to affect the results of our analysis of *e*(*t, z*) in a significant way. The errors can emerge from various sources, including the elimination of names just described.

## Data Availability

Data is available at the link below, with the limitation that names have been obfuscated for privacy protection, certain data has been aggregated, and original data is unavailable due to data use agreements.

https://zenodo.org/uploads/14928060

## Supplementary Information

Supplementary Methods document. Supplementary Discussion document.

## Acknowledgements

We thank Robin Dunbar, Gesine Reinert, Sam Roberts, Amira Roess, and Serguei Saavedra for critical comments, and Bryan Adams, Alessio Cotroneo, Unchitta Kan, Valentín Vergara Hidd, and Alan Zhang for logistical support.

## Declarations

### Ethics approval

Not applicable.

### Consent for publication

Not applicable.

### Data availability

The datasets necessary to reproduce the results presented in this paper are available on Zenodo: https://doi.org/10.5281/zenodo.14927847 ([51]).

### Materials availability

Not applicable.

### Code availability

Not applicable.

### Contribution

J.M. and E.L. conceptualized this work. J.M. performed analyses. All authors contributed to the writing and reviewing of the manuscript.

