## Supplementary material for "The effect of local extended families in epidemics: the case of COVID-19 deaths in the US": SI.pdf

S2.3.4 A theoretical discussion of the quantitative effects of $\alpha_z$ . 40
S2.3.5 Robustness checks and low and high correlation scenarios 42

### S1 Supplementary Methods

#### S1.1 Obituary Data

Here we describe in further detail the processing of the OB data set.

##### S1.1.1 Obituary Data Source Description

We construct the obituary data set from the web, with the corresponding details provided in Methods.

Consistent with the main text, in what follows a label $i$ means, interchangeably, decedent $i$, their obituary, and $i$'s obituary text file.

##### S1.1.2 Obituary pre-processing and obituary corpus

Each obituary text file is formatted as ASCII text with HTML encoding. In addition to the text of the obituary itself, the files also contain HTML code to advertisement that is consistent across files. This code is removed, resulting in output that is a plain ASCII text file of the narrative of each obituary (see example below). The results of this are $1,874,766$ ASCII-format obituary documents. In addition, duplicate URLs arise from inconsistencies in the order in which search results are returned over the time in which we sampled the records, and we keep only one copy (which corresponds to our first de-duplication). At this stage, we obtain our *obituary corpus*, which consists of $1,630,258$ obituary texts.

#### S1.2 Obituary Data Engineering and Natural Language Processing

##### S1.2.1 General strategy

The most challenging and work-intensive goal of the analysis of obituaries is the extraction of names (e.g. "John Doe" or "Jane Doe") and relationships (e.g. father or sister). To achieve this goal, each obituary is analyzed for name-entity resolution (NER) and relationship extraction (RE). While such tasks could be performed through different methods, the complexity and variability in the writing of each obituary together with the size of the data suggests that an ideal approach is the use of Large Language Models (LLMs). Below we explain the steps involved in the process, starting with an example obituary that shows some of the complexities faced and the approaches taken to solve them. We also explain the extraction of decedent gender and age.

The LLM application is performed multiple times, as described in the Methods section of the main text in order to avoid hallucinations. In Methods, we indicate that we seek consistency in outputs of multiple applications of the LLM to the same obituary prompt. In other words, we use the notions of an ensemble method. Broadly described, we must obtain at least two consistent results for a relationship to be used and this relationship is checked back against the obituary to ensure it is contained there.

Our main description (next) in these Supplementary Methods focuses on what happens during one single application of the LLM to all obituaries in the obituary corpus (Sec. S1.2.3). Once we conclude this description, we briefly discuss how multiple applications of the LLM take place, predominantly to clarify how the remaining de-duplication steps occur.

##### S1.2.2 Obituary Example

Before we describe our data extraction and cleaning process, we show next an example of a fictional obituary. From this example, we extract the information we would seek when using our choice of LLM to accomplish NER and RE, making the same choices we will later explain, and giving practical reasons for them.

A *fictional example* obituary may state:

*"Jane Deer of New York, 90, passed away on February 29, 2020. She was a long time resident. She is preceded in death by her father, John Doe, mother Mary Doe, and her brother John Doe Jr. She is survived by her husband Jim 'Jimbo' Deer, loved son Robert Deers, daughter Lizzy, sis Mary (Harry) Rabbit, cousins Jack, May, June, Peter, and Oliver, and many nieces and nephews. Memorial services will he held at Living Memory Funeral Home from March 3 to March 6."*

Some of the challenges faced when performing NER and RE are captured in this example. The underlined text shows some common difficulties encountered which include errors, suffixes, conventions in obituary writing, nicknames, family members without a last name, adjectives to a relationship (loved son), and handling informal writing such as using the term *sis* instead of *sister*. Any of these difficulties can lead to unreliable information. In what follows we explain our process in detail on the basis of this example, highlighting our logic.

| | |
|---|---|
| Jane Deer | self |
| John Doe | father |
| Mary Doe | mother |
| Jim Deer | husband |
| Robert Deer | son |
| Mary Rabbit | sister |
| Harry Rabbit | brother-in-law |

Table S1: **Results of fictitious obituary analysis according to our process.** Tabular presentation of the result of annotating the fictitious obituary with $i$ = Jane Deer presented in the Sec. S1.2.2. The table shows only the names that we would retain based on the process outlined in the section, and in the format we keep.

To provide ground truth, we first show the information a human reader for this obituary would identify. The output follows the conventions described in the main text (Methods). Each line in Tab. S1 is called a *record* and the term applies to both annotated data and data generated by the application of the LLM.

Note how the annotation handles some of the challenges:

1. it has corrected the sons last name to "Deer" and has omitted the adjective "loved" to the son relationship,
2. has associated the relationship of sister to the name "Mary Rabbit", implicitly recognizing that sis refers to sister,
3. has not added brother "John Doe Jr." because his name could create ambiguities in overlap matching with father John Doe,
4. has not added daughter "Lizzy" to the annotation because there is no last name (she may or may not be married and therefore unclear what her surname is),
5. has omitted the nickname "Jimbo" for husband "Jim Deer",
6. has introduced the relationship brother-in-law to the name "Harry" that appears in parenthesis around "Mary Rabbit", reflecting a common obituary convention to indicate married relations in this way,
7. has added the last name Rabbit to brother-in-law Harry,
8. has ignored the list of cousins "Jack, May, June, Peter, and Oliver" as there are no last names stated, and
9. has ignored the reference to "many nieces and nephews" which are relationships for which a name is not stated.

10. has added the decedent "Jane Deer" to the names in the obituary with a relationship of "self."

All these choices reflect the goal of standardizing the way in which relationship information is handled and to ensure that only the most conservatively accurate information is kept, discarding any elements that could generate ambiguities or false positive overlaps among obituaries.

In terms of the variables we collect from this example (where the symbols have been defined in the main text Methods) which are then used for obituary analysis, we have

$$
\begin{aligned}
g_{i=\text{Jane Deer}} &= \text{New York} \\
a_{i=\text{Jane Deer}} &= 90 \\
x_{i=\text{Jane Deer}} &= \text{female} \\
f_{i=\text{Jane Deer}} &= 7 \\
\mathbf{o}_{1,1}(\text{Jane Deer}) &= \text{Jane Deer}; & \mathbf{o}_{1,2}(\text{Jane Deer}) &= \text{self} \\
\mathbf{o}_{2,1}(\text{Jane Deer}) &= \text{John Doe}; & \mathbf{o}_{2,2}(\text{Jane Deer}) &= \text{father} \\
\mathbf{o}_{3,1}(\text{Jane Deer}) &= \text{Mary Doe}; & \mathbf{o}_{3,2}(\text{Jane Deer}) &= \text{mother} \\
\mathbf{o}_{4,1}(\text{Jane Deer}) &= \text{Jim Deer}; & \mathbf{o}_{4,2}(\text{Jane Deer}) &= \text{husband} \\
\mathbf{o}_{5,1}(\text{Jane Deer}) &= \text{Robert Deer}; & \mathbf{o}_{5,2}(\text{Jane Deer}) &= \text{son} \\
\mathbf{o}_{6,1}(\text{Jane Deer}) &= \text{Mary Rabbit}; & \mathbf{o}_{6,2}(\text{Jane Deer}) &= \text{sister} \\
\mathbf{o}_{7,1}(\text{Jane Deer}) &= \text{Harry Rabbit}; & \mathbf{o}_{7,2}(\text{Jane Deer}) &= \text{brother-in-law}
\end{aligned}
$$

##### S1.2.3 One step of Large Language Model Processing: Prompt, raw output, cleaned output, age extraction, gender extraction

***Generalities.*** In order to implement the strategy indicated in Sec. S1.2.1 and generate output consistent with what is shown in Sec. S1.2.2, we next describe the steps that take place for a single application of our method to the obituary corpus. Beyond the straightforward de-duplication step indicated above that discards copies of an obituary due to repeated URLs, more sophisticated obituary de-duplication occurs and it is described in Sec. S1.3.

To accomplish the NER and RE required, we performed prompt engineering and experimentation against an annotated dataset containing 386 obituaries, 5,002 family members, and sampled from 169 cities. This annotated corpus allowed us to check the statistical performance of the LLM in the NER and RE tasks, in addition to details about the LLM output.

***Large Language Model Used.*** We use Google's PaLM text-bison@001 model with a temperature of 0 to limit model creativity in generating responses.

***Prompt Engineering.*** To prompt the LLM, we provide the decedent names to anchor the model on the correct individual for the purpose of identifying relationships (see box below). In testing, we find that output formatting is inconsistent. Thus, although we use zero-shot prompting for the NER and RE tasks, we apply a few-shot example of the desired output format to limit the impact of output formatting inconsistencies (see in box the format given to the names and relationships with father and mother, both with colon). The zero-shot and few-shot prompts we generate use the following format, with variable expressions for the decedent names and obituary text shown in bold and curly brackets.

> In the following obituary text for **{decedent_name}**, extract the names of all of their family members, as well as what family relationship the decedent has with the extracted family member name.
>
> Return the results with one family member per line, the name of the family member first, then the relationship, with a colon in between. This format is shown in an example here:
>
> John Doe: father
> Jane Doe: mother
>
> The obituary text from which you should extract family members and relationships is:
>
> **{obituary_text}**

***Raw LLM Output Processing.*** The raw output obtained from the application of the LLM to each obituary has to be processed to obtain standardized information. In addition, this output has numerous limitations and defects that must be managed to avoid spurious results. We now describe how we standardize the output and handle the defects.

1. As described above, we prompt the LLM to produce output with colon separation between names and relationships. For a single name-relationship record, this should take the form

   *name field*: *relationship field*

   The colon symbol separates two fields: on the left we have the *name field* and on the right we have the *relationship field.* Illustrating this with one record from the example obituary in Sec. S1.2.2, we have:

   John Doe: father

2. We standardize all relationship terms collected from the data to a single consistent term for each relationship. For example, an obituary may list the relationship *grandchild* using one of many different variations, such as *grand-kid*, *grand-baby*, or others. To standardize, we perform visual inspection to generate a dictionary of terms that are mutually exchangeable among a group and pick a standard term for each such group. The 26 standard relationship terms we use are found in Sec. S1.2.6. A file containing all variants of each name is provided in [53].

3. We discard records that do not conform to the output format 'name: relationship' when either the text output has some defect or the output is

not what we seek (for example a non-family relation). In practice, these cases occur in several ways. We enumerate each type of defect next, which are all identified through the use of regular expression techniques:

(a) the separator character (:) is missing,
(b) the name and/or relationship field(s) are blank,
(c) the relationship field contains a non-family relation such as a friend, co-worker, pet, care-giver, etc.,
(d) a relationship term appears in the name field, and
(e) a non-alpha character (e.g. a number) appears in the name field.

In all cases, the appropriate regular expression search in each field is able to detect the issue, leading to the purge of the record.

4. We process names to standardize them to a format of "nameF+blank+nameS" where "nameF" corresponds to a first name, "nameS" to a surname, and "blank" is a single space character between nameF and nameS. To do this, when necessary we eliminate words or abbreviations that vary from this target format, leaving only first name and surname. We discard records where the name field has certain defects that prevent us from obtaining the desired format or where the information given may lead to complications in the subsequent search for overlaps. Concretely, this takes place through the following steps:

(a) We process each raw LLM outputted name by leaving only alphabetic characters (i.e. eliminating numerical characters, punctuation, dashes, hyphens, and multiple forms of white spaces) and then splitting the resulting string on whitespace characters to create an array of strings, such as title, first name, middle name, last name, and suffix.
(b) After the obituary corpus is processed once, we compute word frequencies on the arrays of strings created in 4a, sort the words in decreasing order, and visually inspect them. This detects prefixes and suffixes that refer to titles, ranks, degrees, and common generational references (e.g. Dr., doctor, Esq., Sr., Jr., junior, etc.). We discard such terms, leaving behind only words appropriate to names.
(c) We discard records where the name field has a single word (e.g. only "John" with no surname, or "Doe" without a first name).
(d) We discard names where the first name or surname is a single letter.
(e) When the LLM produces one or more middle names, these are discarded and only first name and surname are left.
(f) We discard names when the field includes a relationship term (e.g. "John Cousin").
(g) We discard cases where a real first name contains a space causing it to appear to be a first and last name (e.g. "Mary Sue").

5. We update each obituary by adding a record with the decedent listed as a family member with relationship of "self". This simplifies obituary matching. We standardize decedent names in the same way as other names in order to allow for consistency in obituary matching.

6. The prior steps of this process may generate repeated names (e.g. John Doe and John Doe Jr. may be different people but the removal of Jr. leaves equal names). This can create difficulties with overlap detection. Therefore, we examine the set of records that remain after the prior steps and discard all but one record with repeated content in the name field. The surviving relationship is a consequence of the order in processing and not subject to a systematic rule.

When the process above leaves an obituary without records other than the decedent itself, and this occurs consistently after the application of the ensemble treatment described in Sec. S1.2.4, the obituary only affects death counts but not overlap counts.

***Determination of gender.*** Obituaries are descriptions of a person and, while they may have several pronouns for mentioned individuals, they are primarily in reference to the decedent. Therefore, we extract decedent gender by counting gendered pronouns and discarding records where the gender is ambiguous. The ambiguous cases include the absence of pronouns or an equal number of masculine and feminine pronouns.

***Determination of age.*** Although we attempted to obtaining age through the LLM, this proved less reliable than extraction through other means, namely a combination of regular expression and HTML pattern headers together with logical rules. We now describe the problem and our approach. It is important to understand first that we process each obituary to extract *candidate* values for age and then, depending on the type of information that led to the candidate age(s), we apply additional logical rules to make a final determination of age.

The initial collections of candidate ages come from three sources.

1. *Age strings* (AS): strings with the correct format to possibly refer to ages (e.g. two or three consecutive digits separated by spaces). These strings can correspond to many types of information, including the actual age of the decedent, number of years of marriage, ages of people other than the decedent, numerical references to the number of family members of some type (e.g. "12 grandchildren"), etc.

2. *Date strings* (DS): strings with the correct format to refer to a date (e.g. two digit month followed by some separator to a two digit day and followed by another separator to a year of two or four digits). These dates can include date of birth, date of death, weddings, date of funeral services, etc.

3. *HTML code* (HTML): scraped data apart from the body of the obituary text that indicate the year of birth and year of death.

Any obituary can have all, some, or none of these types of information, and each type may provide more than one candidate age. To determine a candidate age $C(m)$ from each type of information whenever present ($m$ represents the type of information), we do the following:

1. From *age strings* ($m$ = AS), we record all strings of this type and eliminate those over 120, then choose the maximum if it is greater than 31 (to avoid confusion with numbers representing months and days). Otherwise, we take the first occurring numeric age as our candidate, following the typical obituary-writing practice of including the age immediately after the decedent's name.

2. From $m$ =*date strings* ($m$ = DS), we filter irrelevant dates (prior to 1900 or after 2022), then utilize the earliest date as date of birth, and the earliest date within 30 days of the latest date as the date of death. This is because obituaries frequently include funeral dates, but tend to not contain other dates immediately around dates of death and funerals.

3. From $m$ =*HTML headers* ($m$ = HTML), we calculate an age from two supplied years. Because this number is approximate, we allow for differences of up to a year when comparing to other candidate ages. The nature of obituary writing means that the HTML candidate age tends to be the most reliable due to its appearance in a specific field of HTML rather than being systematically extracted, but not the most precise, as it is merely the difference between the year of death and year of birth.

Given our collection of candidate ages, if there are any, we employ the following process to select which age $a_i$ applies to obituary $i$:

1. We first filter out invalid candidate ages such as negative numbers or values $\geq 120$.

2. If only a single candidate type $m$ is detected, we accept it as correct.

3. If we find a pair of candidate types $m$

   (a) if the candidates reconcile (taking into account $\pm 1$ tolerance for $m$ = HTML), we accept the reconciled age. If this includes an HTML age, we utilize the other among the pair.

   (b) if the candidates do not reconcile, we introduce an order of reliability preference given by "HTML is better than DS is better than AS" and take the $C(m)$ corresponding to the highest preference.

4. If we find all three candidate types $m$

   (a) if the candidates reconcile (taking into account $\pm 1$ tolerance for $m$ = HTML), we accept the matched candidate age of either DS or AS.

   (b) if a pair of candidates reconcile (taking into account $\pm 1$ tolerance for $m$ = HTML), we accept the value in the pair, again preferring the non-HTML age if HTML constitutes one of the pair.

(c) if none of the candidates reconcile, we accept the $m =$ HTML.

5. If we do not find any candidate ages the record is removed from our analysis.

###### S1.2.4 Ensemble treatment of obituaries

The family of large-language models to which text-bison@001 belongs is known to occasionally generate false results called hallucinations, even when provided factual information [54]. We address this by using repeated submissions of each prompt, processing the output as explained above for standardization, and comparing the results (along with other steps described below). We refer to the submission of each prompt and the output standardization as an *execution*. Our approach to handle hallucinations is inspired by the fact that these originate from randomization and are therefore inconsistent over multiple executions, while factual data extraction is consistent.

The concrete steps are as follows. Records of the execution outputs are compared. If the outputs of two executions of an obituary are completely identical, and the relationships and names found are confirmed to be present in the original obituary document, the output is accepted. If in two executions, discrepancies are found, a third execution is performed as a tie-breaker. If two of the three execution outputs are consistent and their information is present in the original obituary document, the majority output is accepted. For the remaining obituaries with unmatched records, we perform a fourth execution. Records are accepted by the same majority rule as in the case of three executions, with the additional condition that the information is also verified as present in the analyzed obituary. Obituaries for which a majority rule still does not apply after four submissions are assumed to have limitations or unclear results and we process them as follows: we break up the output of each execution into individual records that form a set for that output, perform an intersection of the four sets of outputs, and check that any records present in the intersection are found in the original obituary document. Any record that is found is accepted as a name and relationship of the decedent. If the intersection is an empty set, no records emerge from the obituary. In other words, this fourth submission may eliminate records altogether in which, for instance, no family members are listed or the names of individuals are ambiguous or inconsistent across all executions.

###### S1.2.5 Performance evaluation

We carry out two evaluations of performance. First, at the end of processing the obituary corpus, we count the times a family member and relationship are identified for a given obituary. Then we iterate over the first execution and identify true positive, false positive, and false negative records for each family member and relationship and for each obituary. In other words, we are evaluating the model's ability to detect each named entity in an obituary, as well as their relationship to the decedent. We count as true positives records where the family member and relationship are verified by at least one additional execution, false positives those records which are not verified in subsequent executions, and false negatives as those records identified and verified using only subsequent executions. We limit our examination of model validation across executions to a single execution to maintain its integrity, noting that we are examining the stability of model results rather than validating to ground-truth data using this method. The results are 0.9696 precision, 0.9952 recall, and an F1-score of 0.9822.

Second, we hand-annotate a total of 386 obituaries containing 5,002 identified named entities and relationships fitting our criteria. We subsequently apply the same data cleaning process deployed on the obituary corpus to this annotated data set. This allows us to assess the performance of our method. Comparing the result of the process against the annotated sample, we calculate precision of 0.9963, recall of 0.8065, and an F1-score of 0.8914. Comparing these performance metrics to those obtained between multiple executions, the reduction in recall indicates that when the model fails to identify a record, it generally does not improve on subsequent attempts. This comparison also highlights the efficacy of employing multiple executions to reduce hallucinations, as precision between repeated executions and between our final results and ground-truth data are similar.

###### S1.2.6 Relationship name standardization

The LLM output produces slightly over 900 distinct relationship terms. These terms appear because the LLM typically copies the term from the obituaries submitted. We standardize terms for labeled relationships by visual inspection, reviewing all relationship labels provided by the LLM, grouping together those with the same meaning such as grandchildren and grand-kids, and assigning one such meaning a standard designation for the group (list below). The term affinal below indicates the relationship originates through a marriage. For example, in some records from the LLM, we find relationships such as "brother in law", "brother by marriage", "sister's husband", "wife's brother", and others, all of which qualify as affinal (some relationships such as nieces and nephews may be genetic or affinal, which cannot usually be discerned from obituary writing practices). We further contract gendered terms for nieces and nephews to the term nibling, and introduce a corresponding term for aunts and uncles, pibling.

In summary, we identify 26 relationships:

| | | |
|---|---|---|
| child | child (affinal) | cousin |
| cousin (affinal) | grandchild | grandparent (affinal) |
| grandparent | grandparent (affinal) | great grandchild |
| great grandchild (affinal) | great grandparent | great nibling |
| great nibling (affinal) | great pibling | great pibling (affinal) |
| nibling | nibling (affinal) | parent |
| parent (affinal) | pibling | pibling (affinal) |
| self | sibling | sibling (affinal) |
| spouse | spouse (affinal) | |

The complete list of relationship terms determined by the LLM can be found in [53].

#### S1.3 De-duplicating near-duplicate obituaries

Duplicate obituaries can emerge for a number of reasons. Some arise purely due to data processing and are easily identified in our case because of duplicate URL addresses (see Sec. S1.1.2 and Methods). Other duplicates originate from real-world processes such as an obituary submitted to multiple newspapers or submitted to multiple online resources that lead to slightly different posted documents. These real-world *near-duplicate* obituaries are not always found easily and require a robust search method. Near-duplicate obituaries lead to several problems such as wrong counts of deaths by demographic categories and the detection of *fictitious* overlaps based on our method, where the same person with two very similar obituaries may appear to be two different decedents.

To address this problem, we build a method for detection of near-duplicate obituaries. The method is applied after carrying out the steps outlined in Secs. S1.2.3, S1.2.4, and S1.2.6 for, respectively, cleaning and mining information from each of the obituaries, performing the ensemble approach, and standardizing relationship names. In addition, the method relies on a first detection of overlaps over the obituaries that have already been de-duplicated for exact duplication (as explained in Sec. S1.1.2). Overlap identificaiton is necessary because near-duplicates would not be generally found by more direct methods such as regular expression matching. This overlap detection, performed under the rules explain in Methods, is carried out city by city and with the most permissive parameters for overlap detection, namely $\Delta\theta = 5$ years and $m_o = 1$ as this will find the most possible near-duplicates relevant to our analysis. In particular, our approach here safeguards for the errors near-duplicates can cause to the calculations of $w(g, \mathbf{c}, t)$, $s(g, t)$, $r_\mu(g, t)$, and quantities derived from these.

The method requires three steps, guided by the intuition that any two near-duplicate obituaries $i$ and $j$ create overlaps that are very good, not only with a large value for $m_{i,j}$, but also with strong similarity or even equality in several other features: the decedent names $i$ and $j$, the records for names *and relationships* captured in the sets $\mathbf{o}(i)$ and $\mathbf{o}(j)$, gender, age, and/or date of death. Furthermore, the method uses a machine learning classifier, XGBoost [55], to facilitate the visual inspection for final classification of pairs of obituaries as either near-duplicates or not.

In order to implement XGBoost, we use a set of variables for both training and, subsequently, classifying pairs of obituaries as near-duplicates. Let us label two potential near-duplicates as $i$ and $j$. The combination of variables we use include quantities already defined: the number of people mentioned in each of the obituaries ($f_i$ and $f_j$), the size of the overlap $m_{i,j}$ between $i$ and $j$, calendar months of deaths $t_i$ and $t_j$, ages $a_i$ and $a_j$, and genders $x_i$ and $x_j$. Then, we introduce several additional variables that quantify the intuition that near-duplicates create very good overlaps. This practice is generally referred to as *feature engineering* in the field of machine learning.

***Feature engineering for XGBoost.*** We introduce the following new variables characterizing an obituary overlap $i$ and $j$: (i) the ratios of overlap size to names in the obituaries, $m_{i,j}/f_i$ and $m_{i,j}/f_j$, (ii) normalized string similarities $d_{\mathrm{L}}(i,j)$ and $d_{\mathrm{JW}}(i,j)$ between decedents' names $i$ and $j$, respectively based on the Levenshtein and Jaro-Winkler string distance definitions, (iii) the harmonic means $H^{(1)}_{i,j}$ and $H^{(2)}_{i,j}$, the first for the similarities $d_{\mathrm{L}}(i,j)$ and $d_{\mathrm{JW}}(i,j)$, and the second for the fractions $m_{i,j}/f_i$ and $m_{i,j}/f_j$, (iv) the contra-harmonic mean $C_{i,j} = \left[\left(H^{(1)}_{i,j}\right)^2 + \left(H^{(2)}_{i,j}\right)^2\right]/[H^{(1)}_{i,j} + H^{(2)}_{i,j}]$ of the two harmonic means, (v) a set of indicator variables for exact matches on calendar month, age, and gender of decedents $i$ and $j$, and (vi) a dummy (also known as one-hot) encoding for the relationship category $z_{i,j}$, constituted by a group of indicator variables as described below.

Some of the new variables require additional clarification. Regarding $d_{\mathrm{L}}(i,j)$ based on Levenshtein distance, the quantity is calculated by first using the traditional definition for which each string edit contributes one unit of distance, then this quantity is normalized by the string length of the longest of the two names $i$ or $j$, and the result of this is subtracted from 1 to give $d_{\mathrm{L}}(i,j)$ which is bound between 0 and 1. In this way, identical names have a score of 1 and names that require an edit on every letter in the longest name have a score of 0. The score $d_{\mathrm{JW}}(i,j)$ is straightforwardly the Jaro-Winkler similarity, which also ranges from 0, for completely distinct strings, to 1, for identical strings.

The indicator variables for possible matches between age, calendar month of death, and gender, are straightforwardly defined by using Kronecker deltas. Thus, among our new variables, we include $\delta_{a_i,a_j}$, $\delta_{t_i,t_j}$, and $\delta_{x_i,x_j}$ where the result of any of them is 1 if the two arguments are equal and 0 otherwise.

To understand the dummy encoding for relationship type, we first explain briefly where it comes from. When overlaps are found (for this de-duplication and in general), each is given a relationship type $z$. For the pair $i$ and $j$, $z_{i,j}$ is determined directly from information contained in the sets $\mathbf{o}(i)$ and/or $\mathbf{o}(j)$ for direct overlaps, or from the approach described in Methods for indirect overlaps. This allows us to define a dummy encoding for relationship type that is actually a group of three indicator variables, $\delta_{z_{i,j},\mathrm{BL}}$, $\delta_{z_{i,j},\mathrm{AF}}$, and $\delta_{z_{i,j},\mathrm{SP}}$, where only one of them can be 1 for the overlap $i$ and $j$, or they may all be 0 in cases where the relationship type is indeterminate. Although intuitively one expects that near-duplicates may have a tendency to appear as genetic relations (BL), ultimately XGBoost assesses the relevance of these three indicators.

The method steps involved are described next.

***Sample creation to train XGBoost.*** Duplicates are a rare class, which means that a uniformly chosen sample of overlapping pairs of obituaries will contain many more non-duplicate than near-duplicate pairs. To reduce the imbalance in types of pairs (near-duplicate or non-duplicate) that a random sample would create, we generate our sample by favoring the inclusion of near-duplicate obituary pairs. Thus, we start from the entire sample of overlapping pairs of obituaries, calculate the variables above, and sort the pairs on the basis of the value of the contra-harmonic mean. We select the first $50,000$ pairs with the largest values of contra-harmonic mean, which are consistent with a larger likelihood of being near-duplicate pairs. Then, we perform a repetitive process of annotating sets of obituary pairs as near-duplicates or not on the basis of one variable at a time. We use four variables for this task, $d_{\mathrm{L}}(i,j)$, $d_{\mathrm{JW}}(i,j)$, $m_{i,j}/f_i$, and $m_{i,j}/f_j$. To illustrate, we first pick a string similarity on names, $d_{\mathrm{L}}(i,j)$. From the set of $50,000$ largest-contra-harmonic mean pairs, we sort again by $d_{\mathrm{L}}(i,j)$ from largest to smallest and, in that order, annotate obituary pairs from visual inspection as near-duplicates or not. The first pairs have a large likelihood of being near-duplicates. However, as we progress through human annotation, the likelihood of near-duplication drops until most pairs are not duplicates. All the pairs annotated are then stored and excluded from subsequent annotation. We then move to another variable, $d_{\mathrm{JW}}(i,j)$, sort the remaining un-annotated pairs from largest to smallest, and continue annotating until finding near-duplicates becomes rare. The same is done with the other two calculated variables, $m_{i,j}/f_i$ and $m_{i,j}/f_j$. This annotation leads to 953 annotated obituary pairs, 531 of which are duplicates, or 55.7% of the sample. This reduces dramatically the imbalance problem of the sample, allowing for a better training of the XGBoost model.

***XGBoost model training and execution.*** To train the model, we use the 953 annotated obituary pairs along with the parameters shown in Tab. S2. Each pair $i$ and $j$ is assigned the variables $f_i$, $f_j$, $m_{ij}$, $d_{\mathrm{L}}(i,j)$, $d_{\mathrm{JW}}(i,j)$, $m_{i,j}/f_i$, $m_{i,j}/f_j$, $H^{(1)}_{i,j}$, $H^{(2)}_{i,j}$, $C_{i,j}$, $\delta_{a_i,a_j}$, $\delta_{t_i,t_j}$, $\delta_{x_i,x_j}$, $\delta_{z_{i,j},\mathrm{BL}}$, $\delta_{z_{i,j},\mathrm{AF}}$, $\delta_{z_{i,j},\mathrm{SP}}$, and the annotation value that indicates if the pair is a near-duplicate or not. The result of the training is a set of XGBoost parameters (a collection of decision trees) that, when queried about an unknown obituary pair $i$ and $j$, produces a probability that the pair is a near-duplicate or not. Once the model has been trained, we use 50-fold cross validation and observe both accuracy and F1-scores of approximately 0.92. The model is then applied with the resulting parameters on the entire set of obituary pairs. By construction, any pair that XGBoost calculates to have a probability $\geq 0.5$ to be a near-duplicate is labeled as such, and the remaining pairs are predicted not to be near-duplicates. The result of this is $11,020$ obituary pairs predicted to be near-duplicates.

***Determination of near-duplicates.*** The $11,020$ obituary pairs predicted to be duplicates are then sorted by value of probability from largest to smallest. We then apply three checks. First, we check whether each pair fully matches in decedent name, age, gender, date of death, and complete family members and respective relationships. Any pairs that satisfy this match are recorded as near-duplicates. Second, we check if all the names of relatives are the same and, if that is the case, we also accept the pair as a near-duplicate. Third, we visually inspect the remaining pairs to determine if they are near-duplicates. This requires judging whether or not the information provided in the obituary suggests that the two people are the same. We err on the side of caution and label as near-duplicates even when only partial indications of duplication may be present (e.g. $i$ and $j$ may have similar family but the obituaries have

| Parameter | Value |
|---|---|
| Objective function | Binary logistic |
| Evaluation metric | Log-loss |
| Validation folds ($k$) | 50 |
| Number of estimators | 100 |
| Maximum depth | 3 |
| Learning rate ($\eta$) | 0.1 |
| Row subsample | 0.8 |
| Column subsample | 0.9 |

Table S2: **Parameters used in the XGBoost model.** The model is applied as described in Sec. S1.3. We use a binary classification objective function and log-loss error. The $k$-folded validation parameter was chosen to balance results stability and execution times, with the remaining parameters selected from a grid-search.

| Data set | $\langle a \rangle_F$ (sample size) | $\langle a \rangle_M$ (sample size) |
|---|---|---|
| OB | 80.45 (402,002) | 74.89 (422,574) |
| MCOD | 76.21 (7,573,515) | 69.87 (8,286,280) |

Table S3: **Average ages of decedents by gender in different databases.** Average ages $\langle a \rangle_F$ and $\langle a \rangle_M$ at the time of death of males and females, respectively, in the obituary (OB) and multiple cause of deaths (MCOD) data sets. The sample size is indicated in parenthesis in each of the cells.

decedent names that are similar, like "Christopher" and "Chris"). Finally, from the resulting labeled duplicates, we delete the duplicate which lists fewer family members or, if they have the same number of family members, we pick one at random. Since for each near-duplicate there is, by construction of this deduplication method, another obituary with which it overlaps, by eliminating one of the near-duplicates, we also eliminate a fictitious overlap. In total, we eliminate 5,393 near-duplicates and thus the same number of fictitious overlaps. As an additional check, we also visually inspect obituary pairs that fall into the category of non-duplicates according to XGBoost and do not find any near-duplicates.

#### S1.4 Data Statistics

Obituary publication does not occur uniformly across the population. Here, we provide more context about the population tendencies that apply to obituaries.

##### S1.4.1 Basic population statistics of OB and MCOD data sets

We begin by presenting summary statistics of the information captured in the OB data. To have a baseline for comparison, we also present the same quantities

| c \ Dataset | | OB ('18-'22) | %OB ('18-'22) | MCOD: All ('18-'22) | %MCOD: All ('18-'22) |
|---|---|---|---|---|---|
| Female | $[0, 70)$ | 72,369 | 14.95% | 1,663,036 | 10.49% |
| | $[70, 80)$ | 72,586 | 11.67% | 1,230,212 | 7.76% |
| | $[80, \infty)$ | 257,047 | 24.62% | 4,680,267 | 29.51% |
| Male | $[0, 70)$ | 123,308 | 8.78% | 2,808,364 | 17.71% |
| | $[70, 80)$ | 96,222 | 8.80% | 1,668,959 | 10.52% |
| | $[80, \infty)$ | 203,044 | 31.17% | 3,808,957 | 24.02% |

Table S4: **Sample sizes for the OB and MCOD databases.** This information is presented for the demographic strata $\mathbf{c}$ used in our analysis. Alongside, we show percentages of deaths associated with each $\mathbf{c}$ for both the OB and MCOD datasets. The corresponding years for which the numbers and percentages apply are from the beginning of 2018 to the end 2022.

from the MCOD data. This information can be found in Tab. S3, where we show average ages by gender for the two data sets. The OB sample is taken after the application of the filters $D = 0$ and $\hat{d}^* = 5$, consistent with the results shown in the main text. The table clearly illustrates the bias of our data favoring older age samples.

Another analysis we perform is the determination of the samples sizes used to select the demographic strata that we analyze in the main text. In Tab. S4 we display the numbers of obituaries captured in each $\mathbf{c}$ or, symbolically, $\hat{d}(\mathbf{c}) = \sum_g \hat{d}(g, \mathbf{c})$. For this table, we also work with the sample resulting from the application of the parameters $D = 0$ and $\hat{d}^* = 5$ and the set of $\mathbf{c}$ used in the main text.

###### S1.4.2 Relative rate of sampling of deaths by demographic categories. Age biasing

To understand in more detail the way in which obituaries capture deaths across the population, we introduce the obituary relative rate of sampling of deaths by demographic category, $\rho(\mathbf{c})$, calculated as

$$\rho(\mathbf{c}) = \frac{\hat{d}(\mathbf{c}) / \sum_{\mathbf{c}} \hat{d}(\mathbf{c})}{d(\mathbf{c}) / \sum_{\mathbf{c}} d(\mathbf{c})}. \tag{S1}$$

The numerator captures what proportion of deaths in the obituaries fall into the demographic category $\mathbf{c}$; the corresponding proportion is calculated by the denominator in terms of actual deaths. If the obituary sample captured actual deaths uniformly at random, this would make $\rho(\mathbf{c}) \approx 1$ for all $\mathbf{c}$ reflecting that obituaries capture actual deaths at their corresponding proportions. In general, $\rho(\mathbf{c})$ measures how skewed the proportions of deaths for category $\mathbf{c}$ found in obituaries are with respect to their true values.

To determine the behavior of $\rho(\mathbf{c})$ and hence how skewed the obituary sample is, we present $\rho(\mathbf{c})$ in Fig. S1, where we make a finer binning of ages for $\mathbf{c}$ than

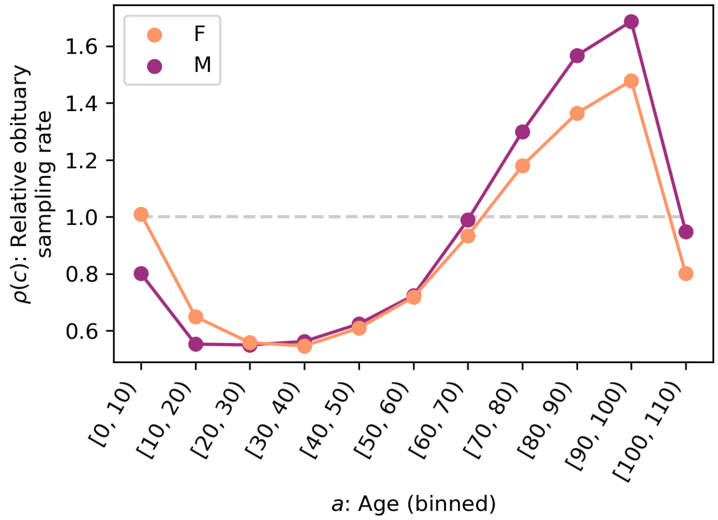

Figure S1: Relative rate of decedent sampling from obituaries, $\rho(\mathbf{c})$ (Eq. S1), split by age and gender. In this plot, age is binned in intervals of 10 year. The rates vary considerably across ages, with gender having a much lesser effect, although males tend to be slightly less represented at young age and more at older age in comparison to females. Around age 70, there is a change from relative undersampling to relative oversampling.

used elsewhere in this research, separating them in intervals of 10 years. We also separate males and females. The plot shows that for ages below 70, obituaries undersample deaths in relative terms. Conversely, once age 70 is reached, obituaries oversample deaths in relative terms. The effect can be quite dramatic, where the ratio between the most pronounced oversampling in contrast to undersampling is near a factor of 3 for males and 2.7 for females.

##### S1.4.3 Ages of local kin-connected deaths

Section S1.4.1 reports summary statistics on age of death information both about the OB and MCOD data sets. However, it is also critical to develop a detailed picture of age effects directly tied to LKDs. To address this, in the current section we present a more focused analysis that compares the age of deaths profiles that focus in more closely at possible COVID-19 effects.

In Fig. S2, we display the probability distributions for the ages of death of several populations for which we have appropriate data from March 2020 to the end of December 2022. Concretely, we show the probabilities of death at age $a$ for (i) MCOD, (ii) deaths encoded as being from COVID-19 in MCOD (labeled MCOD-COVID), (iii) the OB data base with weights taken into account, (iv) the OB data base without using weights, and (v) the combined ages of indexes

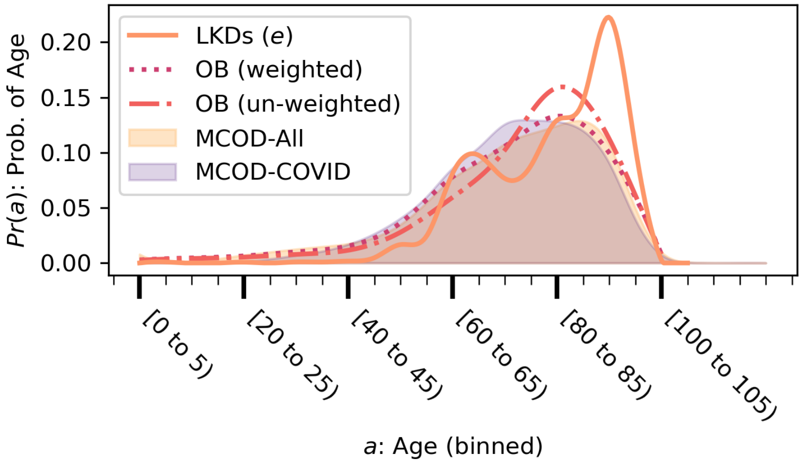

Figure S2: Age probability distributions for several samples. Light purple corresponds to deaths coded as due to COVID-19 in the MCOD data set, light yellow all deaths in MCOD, red curves correspond to OB decedents before (dotted and dashed) and after (dotted) weight corrections, and orange to the indexes and secondaries in overlaps after weighing. All distributions are constructed by binning ages in intervals of size 5 and the smoothing is performed via cubic splines (and it was verified that the soothing does not cause overshooting of the peaks and troughs).

and secondaries in LKDs. The set of COVID-19 deaths in MCOD are those that appear with the International Classification of Diseases version 10 code U07.1. The probability of age $a$ in distribution (v) is constructed by counting excess weighted overlaps where either the index or secondary deaths have age $a$ and normalizing by such counts for all ages. It is evident from the figure that the age profile of LKDs is concentrated to the right, indicating the large representation of the elderly.

The overabundance of detected LKDs among the elderly is likely to be due to a combination of factors. First, as has been clearly determined in prior research, the risk of death from COVID-19 infection increases rapidly with age [56]. Second, considering this against the fact that LKDs are joint events, pairs of elderly people have the largest probability to jointly die from COVID-19 infection among all possible pairs of people within families. Third, the basic bias of obituary sampling, more likely to represent the elderly (see Fig. S1), translates into greater sensitivity in the data to deaths among the elderly. Supporting this last point, it is noteworthy that the age profile of obituaries is indeed more skewed to larger age values in its unweighted than its weighted version (compare dotted and dashed-dotted lines in Fig. S2). Thus, our interpretation of this result is that it likely expresses a combination of a greater tendency of infections within

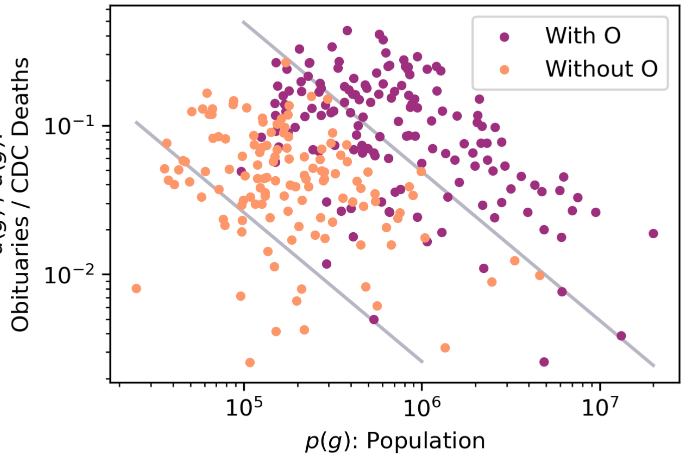

Figure S3: Scatter plot of obituary sampling rates $\hat{d}(g)/d(g)$ as a function of city population $p(g)$. Cities where overlaps are observed ($\sum_t s(g,t) > 0$) are shown as purple dots; cities without observed overlaps are shown in orange. The two curves shown in gray satisfy the functional form from Eq. S2, the upper one with parameter $A$ given by $4.9 \times 10^4$ and the lower one with parameter $A$ given by $2.6 \times 10^3$. These curves bound three regions of the plot, an upper region where all cities show overlaps, a lower region where no cities show overlaps, and a middle region with a mix of cities with and without observed overlaps.

an extended family to lead to deaths among their elders as well as a greater sensitivity of obituary data to detect these deaths. However, the proportional importance of each of these effects is unclear and can only be determined with further research.

One other possible explanation of the age pattern above could be that LKDs are a manifestation of deaths among nursing home residents. However, in main text Sec. 2.4 we consider this possibility and conclude it is unlikely: the temporal pattern of LKDs, $e(t)$, differs from nursing homes deaths. In contrast, nursing home deaths virtually parallel COVID-19 excess deaths $e_o(t)$. Thus, we believe nursing home deaths are a co-residence effect that differs from LKDs. To further support this point, the only known pattern of kin co-residence inside nursing homes that has been reported corresponds to spouses [57], which is in line with our findings in Supplementary Discussion S2.1.

##### S1.4.4 City population obituary sampling rates

As would be expected, obituary sampling rates also vary by city. This variation may play a role in whether the OB data set is able to detect overlaps in any given city (mathematically, whether or not $s(g) := \sum_t s(g,t)$ is either equal or greater than 0 given $g$). To assess sampling rates and their possible role in finding overlaps, in Fig. S3 we present the scatter plot of $\hat{d}(g)/d(g)$ as a function of city population $p(g)$ for the cities that satisfy the filters stated in the main text Methods ($D = 0$ and $\hat{d}^* = 5$), as well as using the overlap parameters $m_o = 4$ and $\Delta\theta = 30$ days. In this analysis, $\hat{d}(g)$ and $d(g)$ are measured from March 2020 to the end of 2022. Population data is obtained from the American Community Survey (ACS) for each CBSA for the year 2020 (5-year sample) [58]. The quotient in the vertical axis is the sampling rate for $g$, i.e. all deaths counted among obituaries in $g$ divided by the deaths in $g$ reported by the CDC. The range of sampling rates is large, going from values just above $2 \times 10^{-3}$ to near $5 \times 10^{-1}$. In addition, the sampling rates show patterns: as population increases from the smallest $p$, there is a general increase in sampling rates until about $p \approx 5 \times 10^5$ where sampling overall begins to decay.

To assess the possible contribution of sampling rates in finding overlaps, Fig. S3 shows cities with $s(g) > 0$ in purple and those with $s(g) = 0$ in orange. We note that checking for observed overlaps ($s(g)$) is an informative approach to learn about the relation between LKDs and city sampling because it is impossible for $e(g)$ to be greater than 0 unless $s(g)$ is as well. In other words, this analysis checks for a necessary condition for $e(g) > 0$. As expected, larger population and sampling rate both make it possible for overlaps to be observed, as evidenced by the purple points generally sitting both to the right and above orange points in Fig. S3. We can also identify a function that relates sampling rate and population, which takes the form

$$\frac{\hat{d}}{d} = Ap^{-\kappa} \quad [\kappa > 0] \tag{S2}$$

above which no city is without overlaps. Intuitively, since the number of deaths in a location is generally proportional to the location's population, we postulate that $\kappa = 1$ and find the value of $A$ that guarantees that any city $g$ such that $\hat{d}(g)/d(g) > A/p(g)$ has observed overlaps. By inspection, the parameter value that satisfies this is $A \approx 4.9 \times 10^4$ (the curve is shown in the figure).

A similar analysis can be conducted to find the corresponding curve below which overlaps are not found. The corresponding parameter for that curve, of the same functional form as Eq. S2, is $\approx 2.6 \times 10^3$ (curve shown on Fig. S3). The region between the two curves is occupied by cities that may or may not display overlaps.

A straightforward algebraic manipulation shows that Eq. S2 can be rewritten as $\hat{d} = Ad/p$, where the right-hand side contains the all-causes mortality rate over the population. Then, cities where $\hat{d}(g) > 4.9 \times 10^4 d(g)/p(g)$ always show $s(g) > 0$. Conversely, cities where $\hat{d}(g) < 2.6 \times 10^3 d(g)/p(g)$ never show $s(g) > 0$. This last case indicates that overlaps require a minimum number of obituary samples to be captured before an overlap can be detected and that this is a function of $d(g)/p(g)$. The clearest conclusion one can draw from this result is that the number of obituaries needed for overlap detection is a linear function of the mortality rate in a city (this is because the $\kappa = 1$ exponent produces curves in Fig. S3 that accurately separate cities). Furthermore, since the

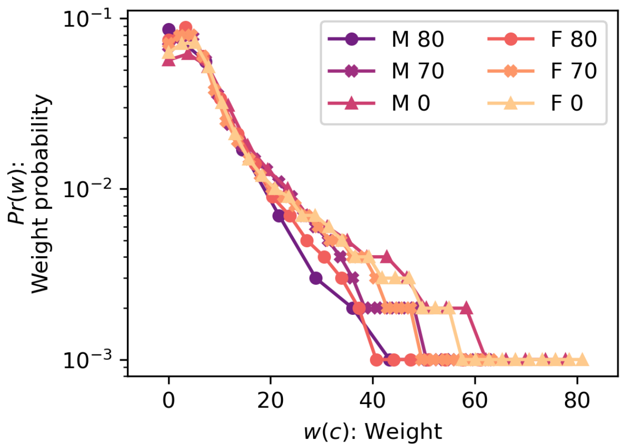

Figure S4: Distribution $\Pr(w)$ of weights $w(g, \mathbf{c}, t)$ grouped by $\mathbf{c}$. The tail of the distributions shows that larger values of $w$ for older age groups are less frequent, an effect consistent with increasing sampling rates with increasing age.

contribution of $d(g)/p(g)$ that causes such LKDs makes it possible for $s(g) > 0$, in principle, this result helps in looking for the frequency of LKDs. However, there are very severe limitations in finding this frequency in this way, such as the fact that $d(g)$ is a time aggregate which includes deaths taking place at times when LKDs may be very low, and that the two numerical coefficients above derive from the nature of the obituary data, which has the numerous shortcomings indicated throughout this work (including age biases, lack or reporting of all family members, tendencies to represent the wealthier population, etc.). Therefore, the utility of this result is quite limited with current data.

To conclude the analysis of this section we point out that, as originally mentioned, there is considerable variability in obituary sampling as well as the ability for those samples to generate a positive signal for LKDs. But given the social nature of the processes that can cause LKDs, this analysis says much more about the sampling limitations of obituaries than about the fact that some cites may experience LKDs while others may not.

##### S1.4.5 Obituary weights statistics

To obtain a more granular view of the sampling of deaths from obituaries, we present in Fig. S4 the distribution of values of weights $w(g, \mathbf{c}, t)$ across the data for the cities that satisfy our filtering criteria for $D = 0$ and $\hat{d}^* = 5$. As explained in the main text Methods, ages are binned into the intervals $[0, 70)$, $[70, 80)$, and $[80, \infty)$ of years of life. The distributions of $w$ are all quite similar to each other up to weights below 20. However, around this value they begin to separate from each other with the likelihood to encounter larger values of $w$ increasing inversely with age (the older the age group, the smaller the weights).

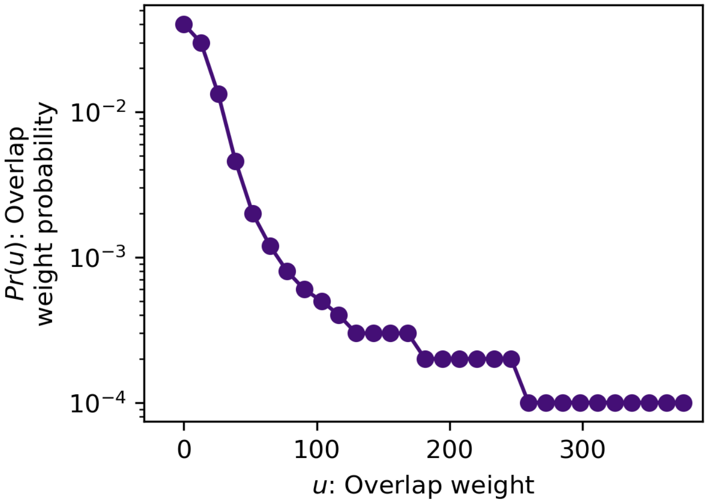

Figure S5: Joint distribution $\Pr(u)$ of overlap weights for all $g$, $t$ and demographic strata $\mathbf{c}$ of index and secondary deaths.

This systematic tendency shows that obituaries sampling improves with age.

###### S1.4.6 Overlap weight statistics

We find that the distribution of overlap weights does not vary significantly among different demographic strata $\mathbf{c}$ for either the index or secondary obituaries and therefore, it is sufficiently informative to show a single joint distribution $\Pr(u)$ to develop an intuition for these weights (see Fig. S5). The weights are calculated applying the family correlations of values $\alpha_{\text{spouse}} = 0.75$ and $\alpha_{\text{non-spouse}} = 0.1$.

##### S1.5 Robustness checks

We now present robustness checks for our results that simultaneously allow us to select the best parameters to use in the main text. We adjust four different parameters: (i) the number of shared names among obituaries ($m_o$), (ii) the date difference between obituaries in an overlap $\Delta\theta$, (iii) the minimum number of obituaries $\hat{d}^*$ we need in order to accept any particular combination of $g$, $\mathbf{c}$, and $t$, and (iv) a tolerance parameter $D$ for possible signs of geographic mismatch between OB and MCOD data. In the checks that follow, the obituary correlation parameters are also set to be equal to the low correlation scenario example used in the main text ($\alpha_{\text{spouse}} = 0.75$ and $\alpha_{\text{non-spouse}} = 0.1$); a thorough discussion of the correlation parameters can be found in Sec. S2.3.

In doing our analysis of overlaps for our main text results, we apply the parameters in a certain order. First, we apply $D$, which eliminates entire cities

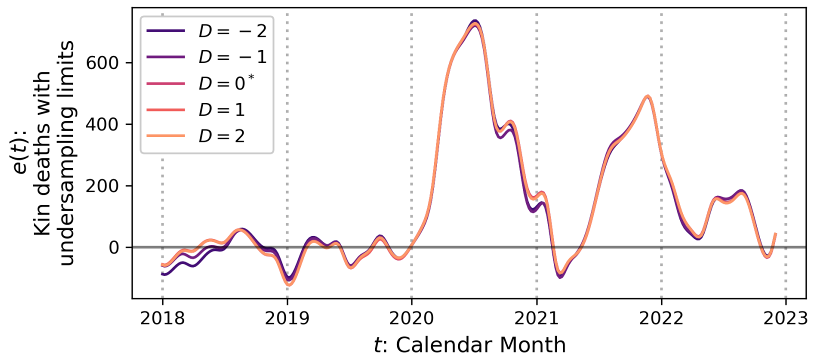

Figure S6: Robustness check of $e(t)$ as a function of $D$. Changes in $D$ produce a minor effect in $e(t)$. For conceptual reasons, we choose $D = 0$ (highlighted in the legend by an asterisk) which is equivalent to eliminating any city in which any month of data shows more obituaries than actual deaths in any demographic category $\mathbf{c}$ that we use.

based on whether *any* combination $\mathbf{c}$ and $t$ in a given $g$ is such that $\hat{d}(g, \mathbf{c}, t) > d(g, \mathbf{c}, t) + D$. The set of cities that survive this filter are then filtered further by eliminating from analysis any $g$, $\mathbf{c}$, and $t$ combination for which $\hat{d}(g, \mathbf{c}, t) < \hat{d}^*$. These filters have the effect of limiting our data and analyses to cities where there is reliable sample. Having applied these filters, we are then left with combinations of $g$, $\mathbf{c}$, and $t$ where we can search for overlaps based on $m_o$ and $\Delta\theta$.

Each check below varies one parameter at a time while maintaining all other parameters fixed to the values used in the main text which correspond to $m_o = 4$, $\Delta = 30$ days, $D = 0$ and $\hat{d}^* = 5$. We show first the checks on $D$ and $\hat{d}^*$ as these limit the data, and then present $m_o$ and $\Delta\theta$.

##### S1.5.1 Effect of threshold $D$

As explained in Methods, since geographic labels of obituaries are affected by the locations of the specific newspapers or funeral homes that generate them, the mismatch of geography can lead to defective sampling of locations where one may find more obituaries than deaths. This effect typically affects small locations more than large ones. For a given location, allowing $\hat{d}(g, \mathbf{c}, t)$ to be greater than $d(g, \mathbf{c}, t)$ attenuates signal rather than enhance it (i.e. makes $e(t)$ smaller because it reduces the values of overlap weights $u$ through a reduction of the values of obituary weights $w$); on the other hand, locations where this defective geographic match occurs could have spurious signal.

We present Fig. S6 where several values of $D$ are exhibited. In the plot, we show how our estimate of aggregate LKDs, $e(t)$, changes based on the value of $D$. Negative values of $D$ are more relaxed, allowing cities with more defective

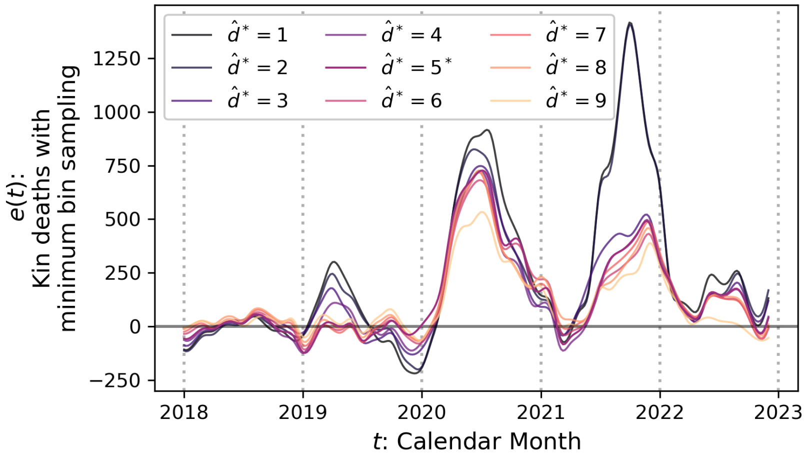

Figure S7: Robustness check of $e(t)$ as a function of $\hat{d}^*$. As $\hat{d}^*$ increases, the magnitudes of the peaks in $e(t)$ gradually diminish. At $\hat{d}^* = 5$ (highlighted in the legend by an asterisk), $e(t)$ becomes stable with respect to additional increases in the parameter, and therefore we select this value for the main text analysis.

sampling to be part of the data analyzed. However, the variation in $e(t)$ is minor with $D$ and it is predominantly in the direction mentioned above (more defective sampling generally decreases $e(t)$). Note that $D = 0$, which we adopt in the main text, enforces the condition that no combination $g$, $\mathbf{c}$, and $t$ has more obituaries than deaths.

##### S1.5.2 Effect of threshold $\hat{d}^*$

Any combination of $g$, $\mathbf{c}$, and $t$ with very poor sampling can lead to unreliable values of $w(g, \mathbf{c}, t)$. Thus, for those cities that survive the application of the filter based on $D$, the condition on $\hat{d}^*$ provides more reliability for the obituary weights we calculate at the expense of reducing the possible obituaries we use.

In Fig. S7, we show how the increase of $\hat{d}^*$ leads to the elimination of more pronounced peaks in the estimation of LKDs, most notably in 2021. At $\hat{d}^* = 5$, the estimates are stable and thus we choose this number as an appropriate value.

After imposing sampling conditions on $D = 0$ and $\hat{d}^* = 5$, 276 cities survive with non-discarded bins. For clarity, we provide the concrete list of cities that remain in [53].

##### S1.5.3 Effect of minimum number of common names $m_o$

Since names are not unique identifiers, one expects that increasing $m_o$ would reduce random overlap detection. On the other hand, one has to be careful not

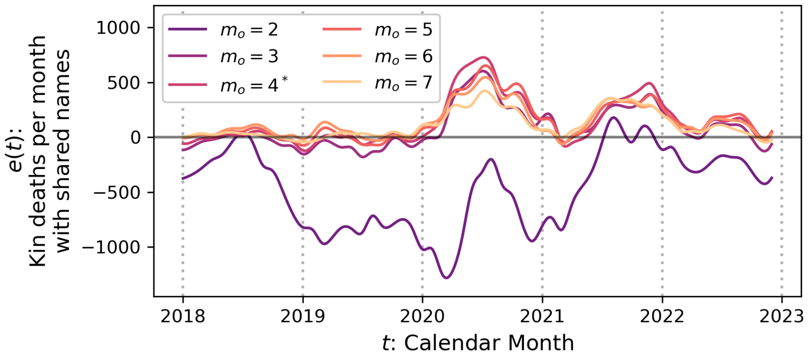

Figure S8: Robustness check of $e(t)$ as a function of the overlap size $m_o$. Values of $m_o \geq 3$ become stable, with $m_o \geq 4$ also leading to a more reliable baseline in $e(t)$ for the years 2018 and 2019. Thus, we choose $m_o = 4$ (highlighted in the legend by an asterisk) for the main text analysis.

to increase this demand too drastically since requiring many shared names can simply discard obituaries that do not mention many names to begin with. We also note that any pair of decedents $i$ and $j$ who share kin are also, individually, related to others in such a way that some of $i$'s kin does not qualify as $j$'s kin and vice versa and, therefore, the use of a moderate value for $m_o$ is justified. We also note that one does not expect this parameter to behave too dramatically given that $r(g, t)$ is also affected by it and therefore an increase in acceptance of overlaps in $s(g, t)$ due to a relatively permissive value of $m_o$ is likely to be compensated by a similar increase in $r(g, t)$.

In Fig. S8, we show the calculation of $e(t)$ constructed with different values of $m_o$. We begin with $m_o = 2$ which shows that two names in common among obituaries are not a reliable condition. The majority of $e(t)$ becomes negative because $r(g, t)$ typically exceeds $s(g, t)$ city by city, signaling that these are mostly random overlaps. However, $m_o \geq 3$ shows stability as the qualitative nature of our results becomes robust to changes in $m_o$. In particular, $m_o = 4$ shows a stabilization of $e(t)$ also during 2018 and 2019, making it our parameter of choice. To provide further context, note that the distribution of numbers of people named in an obituary (excluding the decedent) is relatively narrow. In Sec. S2.1, we discuss this distribution (where spouses are excluded for reasons appropriate to that discussion–see Fig. S14) and find it to be narrow and with an average of 11.95 non-spouse kin mentioned, which means that $m_o = 4$ represents a minimum acceptance of overlaps that covers a significant fraction of names in typical obituaries.

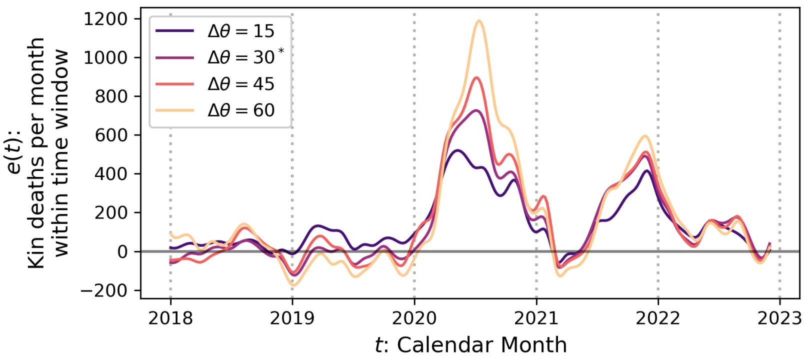

Figure S9: Robustness check of $e(t)$ as a function of $\Delta\theta$. The increase of $\Delta\theta$ consistently shows an increase in the magnitude of peaks in $e(t)$. Our choice of $\Delta\theta = 30$ days in the main text (highlighted in the legend by an asterisk) is based on several reasons, including the observation that this value leaves $e(t)$ close to 0 for 2018 and 2019, and that it is an interval used in determining fatality rates for COVID-19 [61].

##### S1.5.4 Effect of time window between deaths $\Delta\theta$

Kin-connected deaths can occur with a variety to time differences $\Delta\theta$. However, the longer the time, the less likely it is that overlapping deaths may be the consequence of disease propagation. Given that the process of infection and recovery from the initial infection or death from the acute COVID-19 phase takes place within the span of a few weeks [59, 60], we explore how $e(t)$ may be affected by the choice of $\Delta\theta$ taking values of 15, 30, 45, or 60 days. Among these values, one expects that the number of measured LKDs would increase with $\Delta\theta$ since a longer time would be available for overlaps to occur. Note that at the very long limit where $\Delta\theta$ includes all the years of data, our method generates a flat (i.e. no) signal because in that limit both observed and random overlaps become the same (this has been checked and confirmed).

In Fig. S9 we present our exploration of $\Delta\theta$, which shows that the choice of value does not change the qualitative nature of our results in terms of the dates when we observe increased numbers of LKDs. However, as expected, as $\Delta\theta$ increases the peaks of estimated LKDs become more pronounced.

To arrive at the choice of $\Delta\theta = 30$ days, we make several considerations. First, obituary overlaps do not clarify a precise chain of infections from host to host. This means that several possible scenarios may have taken place in terms of infections, such as the index decedent directly infecting the secondary decedent, the index decedent infecting an unobserved intermediary that then infected the secondary decedent, both index and secondary decedents being infected by the same unobserved infective at different times, etc. The quantity $e(t)$ reliably shows that there are associated fatalities that would be hard to explain if it were not for a transmissible cause of death, but not how those

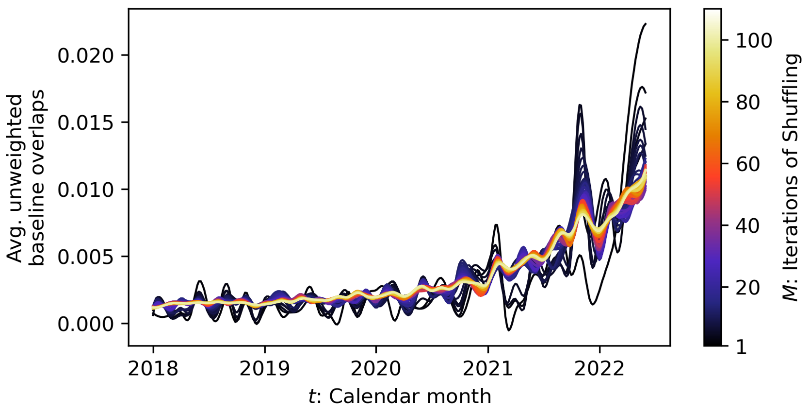

Figure S10: Convergence as a function of $M$ (number of random realizations) of the average count of overlaps for the calculation of the baseline time series $r(t)$. As the plot shows, as $M$ increases the count approaches a stable shape.

transmissions occurred in each case. From the standpoint of the choice of $\Delta\theta$, since we have to allow for the possibility of infections taking various routes between index and secondary decedents, it would be prudent to choose $\Delta\theta$ so that chains of infections of different (although short) lengths are allowed. A second source of uncertainty about how to choose $\Delta\theta$ comes from interactions with the healthcare system. For example, between the first and second waves in New York City (defined for this discussion, respectively, as between March to June of 2020 and November 2020 to March 2021), the time from hospital admission to hospice or death increased from 10.8 to 18.5 days on average, while for the elderly population at least 75 years of age the change observed was from 7.0 to 14.3 days, indicating both the well-reported age cohort vulnerability and differences in medical interventions over time [61]. In addition, while many people make use of the healthcare system as they become acutely ill, some may remain at home as was observed in parts of the US during the COVID-19 pandemic [62]. Those that stay at home typically have a shorter time to fatality than those that go to a hospital, which means that the interval time between two connected deaths may vary by unseen factors such as different choices of treatment routes. It is also instructive that note that a 30 day window is used in empirical studies to determine infection fatality rates for COVID-19 [61]. One final consideration in choosing $\Delta\theta = 30$ days is in Fig. S9 itself, where one can see that for the years 2018 and 2019, this choice is the one that leads to a largely flat (near 0) value for $e(t)$.

##### S1.5.5 Number of realizations to estimate $r(g, t)$

The number of random realizations, $M$, used to calculate the average $r(g, t)$ (main text Eq. 8) is chosen after an analysis of the stability of results as this

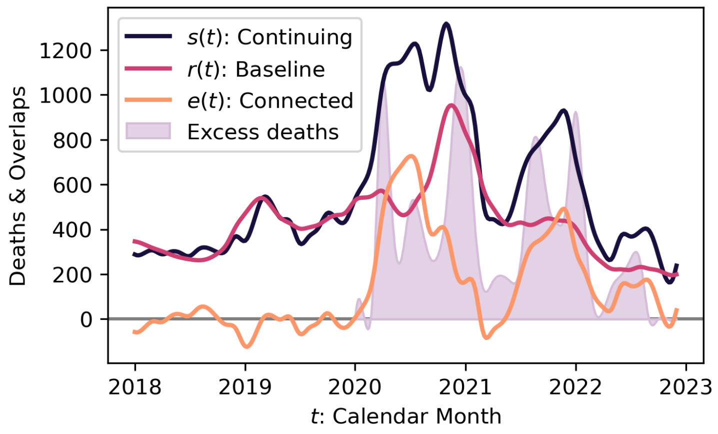

Figure S11: Time series for observed ($s(t)$), baseline ($r(t)$), and aggregate local kin-connected overlaps ($e(t)$) using $\Delta\theta = 30$ days, $m_o = 4$, $D = 0$, and $\hat{d}^* = 5$. For reference, we also present excess death $e_o(t)$ as defined in the main text. The series show how increases in $e(t)$ are predominantly due to increases in $s(t)$ without a matching increase in $r(t)$.

number is increased. In Fig. S10, we present a plot of unweighted $r(g, t)$ (where each overlap has a weight $= 1$) as $M$ increases. This is equivalent to the average time series of count of overlaps, for increasing $M$. The weights are not necessary because they are unaffected by the randomization. From this plot, it is clear that the fluctuations stabilize and that by $M = 100$, the time series has converged.

#### S1.6 Observed, baseline, and estimated LKDs

In order to distinguish the effects of the observed and continuing (baseline) overlaps time series separately and how they combine into the estimated aggregate LKDs, we plot each individually in Fig. S11, along with $e_o(t)$ (excess COVID-19 deaths, defined in detail in the main text). These series are constructed with the main text parameters applied. The separate time series show how the periods of large $e(t)$ typically occur when observed overlaps captured in $s(t)$ become large without a corresponding increase in the number of baseline overlaps $r(t)$, with the exception of the latter part of 2020 when both baseline and observed overlaps increased but the baseline did not fully compensate for the increase.

We also make two observations. First, from the very beginning of 2020 there was an increase in $s(t)$ that was not matched by $r(t)$, happening before the declaration of a pandemic (which took place later, in March of 2020), indicating that obituary overlaps are sensitive enough to detect that COVID-19 was circulating among extended family members and, by extension, the broader population. A second, more technical observation is the fact that between the start of 2018 and the end of 2019, $s(t)$ and $r(t)$ are very similar, supporting the

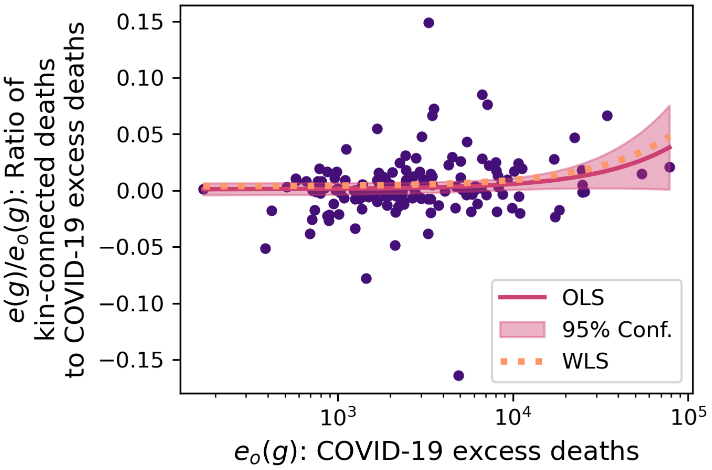

Figure S12: Proportion of local LKDs, $e(g)/e_o(g)$, from March 2020 to August 2022 as a function of local COVID-19 excess deaths $e_o(g)$. Each city $g$ is represented by a dot in the plot. The plot verifies that $e(g)/e_o(g)$ does not vary significantly as $e_o(g)$ increases, supporting the robustness of our results. The absence of a significant trend is captured by the slopes of ordinary (OLS) and weighted (WLS) least-squares regressions, both with slopes $< 10^{-5}$. The weighted least-squares are performed by weighing each $g$ proportionally to the number of obituaries.

logic of our methodology.

#### S1.7 Absence of bias in estimation of LKDs

Since the OB data set is not the product of a controlled sample, it is prudent to check for any possible biases that could lead to systematic errors. In this sense, it is particularly important to determine if the overall number of COVID-19 deaths could have an effect on our estimates of LKDs. We assess this by measuring the ratio $e(g)/e_o(g)$ as a function of $e_o(g)$, where $e(g) = \sum_t e(g,t)$ for $t$ from March 2020 to August 2022 and similarly for $e_o(g)$. The results are shown in Fig. S12. The analysis verifies that $e(g)$ has no significant trend with respect to local COVID-19 fatalities ($e_o(g)$) that could overestimate the number of kin-connected deaths. Specifically, we perform both ordinary and weighted least-squares regressions between $e(g)/e_o(g)$ and $e_o(g)$ and find that the slopes are $< 10^{-5}$, supporting our previous statement (the weighted least-squares are performed by weighing cities by the number of obituaries).

##### S1.8 Plot Smoothing

We perform two types of smoothing, one to attenuate fluctuations in our data from sampling processes, and one to aid in visualization.

For the first, we perform a one-dimensional convolution of an array of data with a kernel [63]. We utilize [1,1,2,3,2,1,1] for our kernel, normalized, as well as mirroring to reflect the edges of the data array. Then, when plotting our data, we interpolate between measurements using a B-spline of degree 3, also known as cubic interpolation. The effect of the convolution is attenuation of our signal, while the interpolation provides continuous lines for plotting purposes. We apply both methods to Figs. 1, 3, 4, and 5 in the main text, and Figures S6, S7, S8, S9, S10, S11, S15, and S16 here.

#### S2 Supplementary Discussion

##### S2.1 Coresidence and non-coresidence

In this section, we discuss the statistics of coresidence and non-coresidence of kin in the US with particular results for the age range that best corresponds to our sample (see discussion on ages among obituaries and overlaps in Secs. S1.4.1 and S1.4.3. Because of the way the US Census and, by extension, the ACS codifies age groups, in this section we emphasize the 65 and older age group which is sufficiently close to the best represented ages in our study. We also discuss the population at large to provide context. The discussion justifies our conclusion that most LKDs measured here are among kin that do not live together.

The key quantity that we need to determine is this: when we find two connected deaths with a relationship of type $z$ between them, what is the probability $\phi_z$ that they live together or apart? The answer to this question depends on $z$ and, as we show below, is very different if $z =$ spouse or non-spouse.

To estimate $\phi_{z=\text{spouse}}$, it is enough to rely on the 2022 ACS 5-year estimate. The information pertinent to this estimate can be found directly from the US Census in the precalculated table [64]. This table provides the number of people age 65 or above who are married and either live with their spouse or are separated (numbers are reproduced here in Tab. S5 for convenience). Because estimates for the probability can be made from information for males or females, in Tab. S5 we present those estimates, calculated from computing the number of males(females) living with their spouse divided by the number of males(females) married (which includes living and separated spouses). At the bottom of the table, we write another estimate of the percentage of people who live with their spouse, calculated by adding the number of males and females married and living with their spouses and dividing the result by the number of married males and females. This leads to

$$\phi_{z=\text{spouse}} \approx \frac{15,933,009 + 12,938,666}{16,256,643 + 13,294,352} = 0.9770. \tag{S3}$$

| | | |
|---|---|---|
| Male, with spouse | 65 to 74 | 10,183,759 |
| Male, with spouse | 75 to 84 | 4,597,049 |
| Male, with spouse | 85 and above | 1,152,201 |
| Total male, with spouse | 65 and over | 15,933,009 |
| Male, separated | 65 to 74 | 226,373 |
| Male, separated | 75 to 84 | 78,929 |
| Male, separated | 85 or above | 18,332 |
| Total male | 65 and over | 16,256,643 |
| % Living together with spouse | 65 and over | 98.01% |
| Female, with spouse | 65 to 74 | 8,953,235 |
| Female, with spouse | 75 to 84 | 3,410,985 |
| Female, with spouse | 85 and above | 574,446 |
| Total female, with spouse | 65 and over | 12,938,666 |
| Female, separated | 5 to 74 | 260,373 |
| Female, separated | 75 to 84 | 77,023 |
| Female, separated | 85 or above | 18,290 |
| Total female | 65 and over | 13,294,352 |
| % Living together with spouse | 65 and over | 97.32% |
| Total | 65 and over | 29,550,995 |
| Average % with spouse | 65 and over | 97.79% |

Table S5: **Coresidence of 65 and older married US males and females.** These numbers show how many people in the age and gender brackets presented by this US Census report live with or are separated from their spouse. The information is taken from the US Census American Community Survey, 5-year estimate 2022. The overwhelming majority of spouses in these age groups live together.

All the values calculated are very similar, near 98%. We note that these are approximations for various reasons, including the fact that the ACS uses sample weights to estimate population values and that our estimates are made from gender groups that are not entirely married to each other. This is because some people can be married to age groups outside the ones we are considering here, reflected in Tab. S5. On the other hand, this is not a far off approximation as the average age gap between spouses that would be 65 or above in 2022 is approximately 3 years [65], which means that indeed the majority are married within the totals shown in the table. The conclusion of this analysis is that when a pair of spouses is found, they have a very high probability to be living together.

The estimation of $\phi_{z=\text{non-spouse}}$ across those aged 65 and older is more complicated. For this purpose, we present two types of estimates given the limitations of the ACS data. The first estimate is based on analyzing the household size distributions. The second estimate takes advantage of the fact that one can calculate the maximum number of possible kin links in a household of size $\sigma$. While the first approach is amenable to focusing on the 65 and older age group, the second is less so. To tackle both approaches, we first discuss household size information in the ACS.

Data from the ACS provides estimates for the number of people $\eta_\sigma$ in the US that live in a household of size $\sigma$. Further, one can divide those people into spouse, family, and non-family to the householder and to specify if the householder is married or not. While in a given household of $\sigma$ people, there may be a mixture of family and non-family occupants, it is an effective approximation for our purposes to assume that out of all the $\eta_\sigma/\sigma$ households of size $\sigma$, all family members are as concentrated as possible. This means that if we were to assign $\eta_\sigma^{(k)}$ (occupants who are kin of the householder) and $\eta_\sigma^{(a)}$ (occupants who are not kin of the householder) onto households of size $\sigma$, we can do so by simply assuming there are $\eta_\sigma^{(k)}/\sigma$ households of size $\sigma$ and in all of those, all occupants are family members. The remaining $\eta_\sigma^{(a)}/\sigma$ households contain no family relations. To maintain consistency, $\eta_\sigma = \eta_\sigma^{(k)} + \eta_\sigma^{(a)}$. The effect of this arrangement is to concentrate family links into coresidence.

Let us label the number of households of size $\sigma$ where all occupants are kin as $\nu_\sigma$. Since the data allows us to identify householders that are married or not, we introduce $\nu^{(h_o)}$ to symbolize the number of such households where the householder is not married and $\nu^{(h_1)}$ to symbolize the case when he/she is; $\nu_\sigma = \nu^{(h_o)} + \nu^{(h_1)}$. The values of $\nu_\sigma^{(h_o)}$ and $\nu_\sigma^{(h_1)}$ are shown in Fig. S13, which decay exponentially with $\sigma$. Then, the average number of people that live in each of these two household types can be calculated via $\sum_\sigma \sigma\nu_\sigma^{(h_o)}/\sum_\sigma \nu_\sigma^{(h_o)}$ and $\sum_\sigma \sigma\nu_\sigma^{(h_1)}/\sum_\sigma \nu_\sigma^{(h_o)}$; the same equation applied to $\nu_\sigma$ gives the average household size. When applied to householders aged 65 and over, the average size of households using $\nu_\sigma^{(h_1)}$ is 2.1677 whereas using $\nu_\sigma$ the value is 1.6884. This clearly shows that the typical household for a householder 65 and over is usually either a single occupant household or one with one other family member. Among

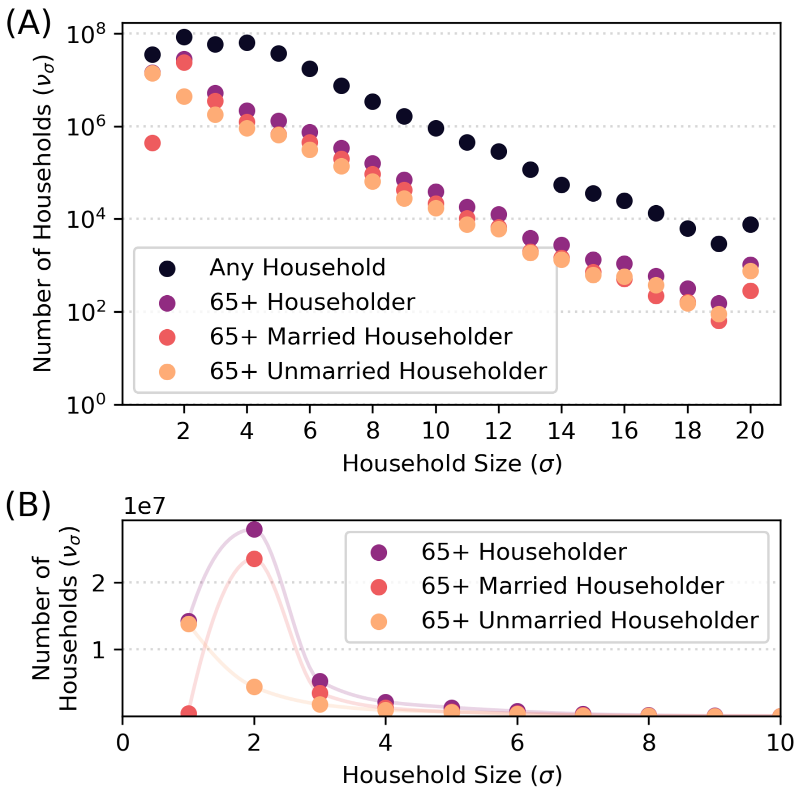

Figure S13: Numbers of households $\nu_\sigma$ by size $\sigma$ with kin-only dwellers. The black curve corresponds to all the US population, the purple curve to households where the householder is aged 65 or older, the light purple curve to households where the householder is aged 65 or older and married, and the orange curve to households where the householder is aged 65 or older and unmarried. Householders 65 and older are predominantly divided by marriage in that those that are married are much more likely to live with their spouse and those that are unmarried are much more likely to live alone. Panel A shows the entire range of $\nu_\sigma$ as we are able to extract from the ACS data, and the vertical axis is in logarithmic scale in order to properly show the detail related to large households. Panel B shows a linear version, limited to $\sigma \leq 10$ which aids in seeing the details of $\nu_\sigma$ for smaller households. The light curves added to the plot are included for visual effect, to draw attention to the changes occurring over different types of households, but are not meant as interpolations, as the data is fundamentally for integers.

those that are married, the average household size is just over two people and thus it means that typically there is only coresidence with the spouse. Visually examining Fig. S13, one clearly sees that $\sigma = 3$ is less probable by an order of magnitude than $\sigma = 2$ in the case of married householders, whereas for unmarried householders $\sigma = 2$ is about one order of magnitude less probable than $\sigma = 1$. In other words, the typical situation is that 65 and older people who are unmarried live alone, and those that are married live with their spouse.

The analysis above notwithstanding, it would be very useful to have an estimate of $\phi_{\text{non-spouse}}$ as the complement to $\phi_{\text{spouse}}$ calculated above. This analysis would provide an assessment of the amount of family coresidence taking place in the tails of the curves in Fig. S13. One difficulty with making this estimation is that while we are able to identify householders by their age, it is not possible to determine the ages of the additional $\sigma - 1$ other people living in the household. Because of this limitation, in what follows we make an estimate of $\phi_{\text{non-spouse}}$ for the entire population from the 2022 5-year ACS estimate. The calculation requires two separate types of information. These are the numbers of households for married and unmarried householders, $\nu^{(h_o)}$ and $\nu^{(h_1)}$, and the sizes of family (i.e. number of kin) for each person, which we estimate here from our OB dataset. For this, we take the obituaries after the application of the $D = 0$ and $\hat{d}^* = 5$ filters.

Let us first focus on the case when there is no spouse in the household, i.e., a household constituted entirely of non-spousal kin. We now introduce a reasonable but overestimating assumption that any two family members in such a household are also family members with one another, leading to a total of $\binom{\sigma}{2} = \sigma(\sigma - 1)/2$ family relations. Therefore, over the US, we estimate that such households contain

$$\ell^{(h_o)} = \sum_{\sigma=2} \binom{\sigma}{2} \frac{n_\sigma^{(h_o)}}{\sigma} = \sum_{\sigma=2} \frac{\sigma - 1}{2} n_\sigma^{(h_o)} \tag{S4}$$

non-spousal kin household links. The fraction $n^{(h_o)}/\sigma$ corresponds to the number of households in the US with $\sigma$ kin members.

Households where there is a spouse follow basically the same pattern but one kin connection needs to be excluded (the spousal connection). Therefore, the number of non-spousal kin ties is given by

$$\ell^{(h_1)} = \sum_{\sigma=2} \left[ \binom{\sigma}{2} - 1 \right] \frac{n_\sigma^{(h_1)}}{\sigma}. \tag{S5}$$

To determine $\phi_{z=\text{non-spouse}}$, we also need an estimate of the number of non-spousal kin connections $\ell^{(k)}$ people have across the US regardless of whether or not they live in the same household. For this purpose, we combine data from the ACS and our OB data set to generate an estimate of non-spousal kin links. We do not use information about family size from the ACS because this is only household information. The 2022 ACS 5-year estimate calculates a total of $n^{(k)} = 331,097,594$. For each individual, the number of non-spousal kin

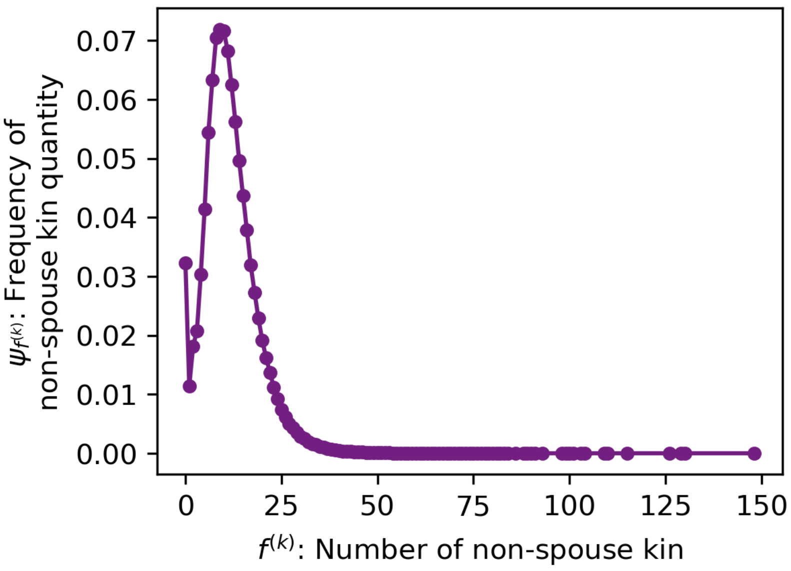

Figure S14: Distribution $\psi_{f^{(k)}}$ of the number of non-spousal relatives $f^{(k)}$ reported in the obituaries obtained after filtering for $D = 0$ and $\hat{d}^* = 5$. The distribution is fairly narrow with a peak at $f^{(k)} = 10$ and an average of $\langle f^{(k)} \rangle = 11.95$. We note that for the fully de-duplicated obituary corpus but without the filters just mentioned, the average is almost identical, with value$\langle f^{(k)} \rangle = 11.63$.

members is a random variable $f^{(k)}$. We now make the simplifying assumption that the number of non-spousal family members is distributed as in the OB data set. With this assumption in place, we can use network concepts to make the estimates. First, we write $\psi_{f^{(k)}}$ for the probability that any individual has $f^{(k)}$ non-spousal links. This means, in turn, that there are $n^{(k)}_{f^{(k)}} = n^{(k)}\psi_{f^{(k)}}$ people with with $f^{(k)}$ non-spousal links. By the well-known handshaking lemma of graph theory for an undirected unweighted network [66], this also means that

$$2\ell^{(k)} = \sum_{f^{(k)}} f^{(k)} n^{(k)}_{f^{(k)}} = n^{(k)} \sum_{f^{(k)}} f^{(k)} \psi_{f^{(k)}} = n^{(k)} \langle f^{(k)} \rangle, \tag{S6}$$

where $\langle f^{(k)} \rangle$ is the average number of non-spousal kin members mentioned in obituaries.

With these result, we can now explicitly produce an estimate of $\phi_{z=\text{non-spouse}}$ which, to be more precise, it is basically an upper bound for this number for the US population, given the way we assign kin to households of given size. Concretely,

$$\phi_{z=\text{non-spouse}} < \frac{\ell^{(h_o)} + \ell^{(h_1)}}{\ell^{(k)}} = \frac{2 \times \left(\ell^{(h_o)} + \ell^{(h_1)}\right)}{n^{(k)} \langle f^{(k)} \rangle}. \tag{S7}$$

The data from ACS needed to apply Eqs. S4 and S5 can be found in Tab. S6, which lead to $\ell^{(h_o)} = 224,580,455$ and $\ell^{(h_1)} = 96,143,143$ to the nearest integers (given that the Census does not separate $\sigma > 20$ from $\sigma = 20$, we approximate the calculations by assuming these household populations are of size $\sigma = 20$, which is a very small correction based on the numbers–see Tab. S6). To determine $\ell^{(k)}$ from Eq. S6, we use the probability distribution displayed in Fig. S14 for the number of non-spousal kin $f^{(k)}$ mentioned in our filtered sample of obituaries, which yields $\langle f^{(k)} \rangle \approx 11.95$. Introducing all relevant numbers into Eq. S7, we obtain

$$\phi_{z=\text{non-spouse}} < \frac{2 \times (224,580,455 + 96,143,143)}{331,097,594 \times 11.95} \approx 0.162. \tag{S8}$$

It should be noted that both Eqs. S4 and S5 do not take into account the possibility that some of the relatives living inside a household are married to each other. However, the only effect this would have is to further reduce the size of the numerator in Eq. S7 further supporting our point. Furthermore, ongoing demographic research suggests that currently in the US the number of living kin members to a person may be considerably larger than what we calculated for $\langle f^{(k)} \rangle$ (perhaps by more than double), indicating that $\phi_{z=\text{non-spouse}}$ may be also smaller than our estimate [67]. Thus, this is yet another effect that reduces the value of the estimate we are making.

The conclusion of this analysis is that, while the likelihood of a married couple to be coresident is quite close to 1, the equivalent chances of coresidence for non-spousal kin is at most around 0.162 based on a population average. Finally, based on inspection of Fig. S13 and the fact that $\nu_{\sigma=1}$ and $\nu_{\sigma=2}$ for people 65 and older have a relatively larger contribution to the entire curve $\nu_\sigma$ than the equivalent curve for the entire population, it is plausible that the coresidence network is even more dilute than what our full population estimate above shows.

#### S2.2 Extended discussion of possible factors affecting periods of low or high LKD activity

In the main text, we provide a compact discussion about the possible mechanisms acting to produce the large surges in LKDs we find in this work. Next, we provide additional arguments and details in support of our interpretation.

While at this stage one can only speculate about the dynamics underlying periods $P_1$ through $P_5$, some clues can be obtained from the dates when each of the periods occurred. Period $P_1$ takes place during the initial months of the pandemic, when the most stringent NPIs where put in place and some people even elected to self-isolate [68, 69]. Also, the provision of child care through schools [70] or pre-schools [71] drastically diminished, affecting the ability for parents to work. It is plausible that these demanding circumstances may have led people to turn to family for companionship in a way consistent with the social theory on extended family [72, 73, 74]. In addition, several studies have shown direct empirical evidence that people sought in-person interaction with family during this period [75, 76], including for childcare support (the largest non-household support by far came from non-coresident family) [77]. The beginning of the subsequent period, $P_2$, in which aggregate LKDs decayed starting in September 2020, when some school districts began to reopen, further supports this hypothesis that the search for support from family at least partially fueled $P_1$. It should be noted that the low levels of aggregate LKDs in $P_2$ are not at odds with the general increase of COVID-19 cases for the 2020 holidays which may have been driven more by intercity travel (which partially rebounded from its minimum early in the pandemic [78]) as opposed to continual interaction with local family.

The increase of aggregate LKDs in 2021 ($P_3$) may be the result of a combination of factors, including again possible school year effects for children, a desire to reconnect face-to-face with family, fatigue to the pandemic along with diminishing adherence to physical distancing measures [79], and perhaps even waning of immunity gained from the first versions of COVID-19 vaccines. Note, in addition, the preponderance of female-to-female genetic overlaps in $P_3$ (as seen from Figs. 3 and 4 of the main text), an effect that suggests the activation of gender roles among family. It is known that women are more active "kin keepers" than men [72], which may have led to more frequent encounters among females after the introduction of vaccines and the easing of some NPIs and, consequently, a significant increase in female-to-female overlaps. These observations are further supported by main text Fig. 5, which shows similarities between LKDs and deaths from COVID-19 among elderly groups.

Periods $P_4$ and $P_5$ are considerably less intense in comparison to 2020 and 2021 but show some qualitative similarities: a depressed aggregate LKDs-pattern at the start of the year followed by an increase, roughly matching excess COVID-19 deaths, during the later part of the spring (similar again to the school year approaching an end). On the other hand, by 2022 social patterns began to be normalized considerably and this may be the reason why aggregate LKDs and COVID-19 excess deaths (as well as aggregate LKDs among genetic and affinal relations–see Fig. 3 of the main text) appear to be more similar.

#### S2.3 Effect of correlated obituary writing behavior on LKDs

##### S2.3.1 Background

As explained in the main text, we introduce $\alpha_z$ to account for two effects, (i) the possibility that the publication of an obituary for one family decedent signals a greater likelihood of publication for another family decedent (including the limit choice of no publication at all), and (ii) that the choice of where obituaries are published makes them detectable in our sample. Both (i) and (ii) express correlations in writing behavior: (i), for example, can be affected by cultural norms and affective motivations; (ii) can express correlations in the availability of venues to publish obituaries and wealth available to pay for them. In addition, (i) and (ii) are mediated by who writes the obituaries. Research about online memorials (similar documents to online obituaries but with opportunities for readers to post comments and perform other activities) has found that the most likely writers of obituaries are the adult children of the decedents [80, 81]. Therefore, to a first approximation, one can assume that the obituary writer(s) of decedent spouses may have belonged to the same nuclear family in the past, would treat obituary writing in the same way (i.e. under cultural norms of that nuclear family), and may even be a single author for both obituaries. The same empirical finding also indicates that correlation would weaken for other types of relationships $z$, even genetic ones, as the children of two non-spouse kin-connected decedents with overlapping obituaries are less likely to have been members of the same nuclear family, distancing the potential equivalences in attitudes towards obituary writing for the two decedents. The existence of choices on where to publish obituaries plays an important role as well, with many people in recent years opting to use a variety of online venues (see [80, 81]) not typically captured in our sample, which is biased towards newspapers and funeral home websites. One useful observation regarding venue choice is the fact that our sample, as large as it is, has a sampling rate near but below $10^{-1}$ of deaths for the period studied, a value we return to in our discussion below.

##### S2.3.2 Correlation Scenarios

Based on the considerations above, the values of $\alpha_z$ we emphasize in the main text and other parts of the Supplementary Methods and Supplementary Discussions assume that spouses are very likely to see correlated obituary writing behavior and that other kin ($z =$ non-spouse), instead, have low obituary writing correlation. While the concrete numerical values used in the main text can be justified in several ways (see end of this section and Sec. S2.3.3), we realize that without a ground truth to guide us, it is necessary to explore the space of values of $\alpha_{\text{spouse}}$ and $\alpha_{\text{non-spouse}}$ to understand the possible impact these numbers could have on our findings. To conduct this exploration, we only consider positive or null correlations and, consequently, $0 \leq \alpha_z \leq 1$ for all $z$.

We first note that in the main text, we quantify the magnitude of the yearly peaks of $e(t)/e_o(t)$ in the space $[0,1] \times [0,1]$ of $(\alpha_{z=\text{spouse}}, \alpha_{z=\text{non-spouse}})$ (shown in Fig. 2). Second, in Sec. S2.3.5 we study the impact of the values of $\alpha_z$ on the entire time series $e(t)$, including seasonality observed in Fig. 1, to determine the robustness of our results. We fix the other parameters involved in $e(t)$ to their main text values, i.e. $m_o = 4$, $\Delta\theta = 30$ days, $D = 0$, and $\hat{d}^* = 5$. In addition, since correlations affect the contributions $\lambda(z)$ and $\lambda(y)$ (see main text Eq. 2), we also present results for these quantities. Third, to aid the discussion in the main text of the key features we find in this Sec. S2.3, in Sec. S2.3.5 we define two scenarios we refer to as low and high correlation. Each scenario is defined by a combination of value ranges for $\alpha_{\text{spouse}}$ and $\alpha_{\text{non-spouse}}$, shown in Tab. S7. In both scenarios, $\alpha_{\text{non-spouse}} \leq \alpha_{\text{spouse}}$ since this makes the most intuitive sense. We use these scenarios to illustrate what may occur to LKD estimates under contrasting levels of correlations, distinct from each other by the range of values that $\alpha_{\text{non-spouse}}$ takes.

To conclude this general discussion on correlation values, we note that it is our belief that the low correlation scenario is more plausible than the high correlation one. A strong indication of this comes from the sampling rate that obituaries offer on deaths in the period studied. We expect that the deaths missing from our data are due to three factors, decedents not being given an obituary, obituaries being published in venues that we were not able to sample, and a lower than 1 correlation in obituary writing behavior within families. For our purposes, all these factors contribute to a lowering of $\alpha_z$ as we define it. Therefore, the sampling rate $\leq 10^{-1}$ (see Sec. S1.4.4 and main text Introduction) can be interpreted as an average value for correlation within our data, which we can associate with $\alpha_{\text{non-spouse}}$ because $z =$ non-spouse constitutes the most common relationship among overlaps. Note that the value for the spouse relationship of 0.75 in the main text is simply a reasonable larger correlation that is compatible with some variability that can occur for spouses in obituary writing because their obituaries can still have different authors (such as one of the spouses before their own death, a grandchild, a different child, or children from different prior marriages in the case one of both spouses were previously married to others). In any case, as the results in Sec. S2.3.5 and those in Fig. 2 of the main text show, the effect of $\alpha_{\text{spouse}}$ is minor for LKDs given that the majority of overlaps are not among spouses (see Tab. S8).

###### S2.3.3 Estimating non-spouse obituary writing correlation based on wealth

One possible driver for the low correlation of obituary writing could be wealth, due to the fact that obituaries constitute an expense. In addition, considering that an obituary is only one of the multiple significant expenses that come with funeral arrangements, one if further encouraged to take into account its potential relevance. We acknowledge it is unclear that by itself wealth can fully explain variation in obituary writing behavior, but it may play an important role in the choice of venue where it is published which, in turn, may make it undetectable to our collection method. The analysis we perform next offers a picture of how some relevant family factors could lead to a resulting value of correlation. To develop a principled estimation, we combine empirical results that measure family correlation of wealth as well as notions about family structure.

We employ two sources of information for wealth correlation. First, intergenerational wealth correlation has been measured between the incomes of fathers and sons to be approximately 0.4 from the Panel Study on Income Dynamics [82]. For this discussion, we use $\alpha_1 \approx 0.4$. Second, studies of homogamy have estimated that the correlation between the wealth of the parents of married couples is approximately 0.3 [83]. Thus, we define $\alpha_2 \approx 0.3$.

On the basis of these values, we can crudely estimate orders of magnitude for the wealth correlation between extended family members. Siblings of adult parents are correlated to the order of $\alpha_1^2 \approx 0.16$. Correlations between genetic kin separated by an additional generation have correlations of the order of $\alpha_1^3 \approx 0.064$. The nearest affine relationships have correlations of the order of $\alpha_1\alpha_2 \approx 0.12$, with the second nearest level correlated to the order of $\alpha_1^2\alpha_2 \approx 0.048$. For ease of discussion, correlations involving a product of two $\alpha$s are said to be of order 2; correlations involving a product of three $\alpha$s are said to be of order 3.

The values calculated above provide a rough order of magnitude of wealth correlation depending on a notion of kin *distance* which is equivalent to order as defined in the previous paragraph. It should be noted that greater distances are associated with greater numbers of kin by the combination of the multiplicative process behind reproduction and the connections between such reproduction trees via marriage. By these arguments, averaging the correlation factors above, more of them of order 3 would be used for a typical extended family than those of order 2. It is not necessary to perform any detailed calculations. Instead, it is simple to note that an $\alpha_{\text{non-spouse}} \approx 0.1$ is a conservatively reasonable approximation given the numbers in the previous paragraph.

The similarity in values between this estimate and the sampling rate of our obituary sample is probably not entirely coincidental but the exact relation between them can only be determined through future research.

##### S2.3.4 A theoretical discussion of the quantitative effects of $\alpha_z$

In order to provide an intuitive description of the role played by writing correlations, we discuss the effect of $\alpha_z$ on $e(t)$ in general terms. To this end, we first clarify that one way to understand $\alpha_z$ is as a bias that increases the likelihood that overlaps are recorded in the data. This is because the larger $\alpha_z$ is, the more likely that two kin-related people that die are sampled in our data. Conversely, the lower $\alpha_z$ is, the less one can guarantee that both people are sampled in our data and thus an overlap becomes closer to a random sampling process. Therefore, when a small value of $\alpha_z$ is assumed, one is expressing the belief that many overlaps are not sampled and that each observed overlap should be responsible for representing many unobserved overlaps. Conversely, if one assumes a large correlation, this indicates a belief that the overlaps have a preferential tendency to be written and thus need not be representative of many unobserved overlaps. As we have said above, for us to estimate the number of overlaps in the population as a whole, we must assume the magnitudes of such writing correlations as they have not been reported.

To describe the quantitative nature of the relation between the magnitude of $e(t)$ and writing correlations, we recall the main text Eq. 5 which captures the effect of $\alpha_z$ on overlap weights. Evaluating the equation with $\alpha_z = 1$, it is straightforward to see that the weight $u_{i,j}$ of an overlap between $i$ and $j$ becomes equal to the inverse of the probability to sample the index $i$; that inverse is itself the sample weight of $i$ (main text Eq. 6). Therefore, in this limit, the weight of the overlap is given only by the weight of the index obituary. This has a direct interpretation: if we assume that the secondary obituary is *guaranteed* to be sampled given that the index has been sampled (i.e. $\alpha_z = 1$) then the overlap's weight is merely a consequence of the sampling of the index. Stated in terms of representativeness, because once the index is sampled the overlap is sure to be sampled, the only sampling that determines if an overlap is observed is that of its index, making the overlap weight equal to the index weight.

The opposite limit is that of $\alpha_z = 0$ which, from main text Eq. 5, shows that the weight $u_{i,j}$ of the overlap between $i$ and $j$ is the product of the weights for $i$ and $j$ (where both weights use main text Eq. 6). Conceptually, this is the case when the sampling of both the index and secondary obituaries of overlaps are independently and identically distributed events, with a joint sampling probability that is equal to the product of their independent sampling probabilities. We can interpret this case by comparison to the case of total correlation. When $\alpha_z = 1$, once we sample the index of an overlap the secondary is guaranteed. In contrast, with no correlation, the secondary is only observed with rate equal to its sampling rate, which means overlaps would be observed in the data less frequently than with any positive correlation between index and secondary. Therefore, the weights predicted by the uncorrelated case are the largest possible since they assume the most unobserved secondary overlaps, and lead to the largest estimates of the magnitude of $e(t)$. (We note that here we exclude the possibility of anticorrelation between obituaries as we have not identified a possible social process where that would be expected).

Mathematically, it is easy to see from main text Eq. 5 that $1/u_{i,j}$, the inverse of the overlap weight, monotonically increases with $\alpha_z$ as exhibited by

$$\frac{d}{d\alpha_z}\left(\frac{1}{u_{i,j}}\right) = q_i \times \left(1 - \frac{1}{w_j}\right) \tag{S9}$$

which is $> 0$ given that $w_j$ is greater than 1. Hence, increases of $\alpha_z$ lead to a monotonic decrease of $u_{i,j}$, consistent with our intuitive discussion above. The result is a statement that, as the likelihood of sampling secondaries decreases with decreasing correlation, each overlap has to account for more unobserved overlaps in the population.

##### S2.3.5 Robustness checks and low and high correlation scenarios

***Strategy for unrestricted exploration of*** $\alpha_{\text{spouse}}$ ***and*** $\alpha_{\text{non-spouse}}$***.*** To concretely measure correlation effects on our analysis, we first explore $\alpha_{\text{spouse}}$ and $\alpha_{\text{non-spouse}}$ broadly (not constrained to the scenarios defined above). Our approach is to explore one parameter at a time, allowing the other to vary between 0 and 1. As is made clear by our results, this approach is sufficient to elucidate the effects of correlation writing on $e(t)$.

Two practical points arise in interpreting the results presented next. First, in an unrestricted exploration of writing correlations, some results concern $\alpha_{\text{spouse}} < \alpha_{\text{non-spouse}}$ which we have indicated above is likely not well justified qualitatively. However, we feel it is useful to show these as a matter of transparency. To interpret these results, one must remember that $e(t, z = \text{spouse})$ and $e(t, z = \text{non-spouse})$ do not have to have the same temporal patterns because, as discussed in Sec. S2.1, in our sample spouses typically live together and non-spouses typically do not. This means that for sufficiently larger correlations of $\alpha_{\text{non-spouse}}$ over $\alpha_{\text{spouse}}$, the temporal pattern of $e(t)$ would be dominated by that of $e(t, z = \text{spouse})$, different than what we observe in the results through most of our analysis (see e.g. main text Figs. 1, 3, 4, and 5), which are dominated by $e(t, \text{non-spouse})$. This occurs because 71.5% of all detected (unweighted) overlaps from 2020 to the end of 2022 are among non-spousal kin (and, correspondingly, 28.5% of overlaps are among spouses). We make this point merely for clarity, but we are deeply skeptical that the regime of $\alpha_{\text{spouse}} < \alpha_{\text{non-spouse}}$ is relevant in reality. The second practical point we highlight is that, because the magnitude of $e(t)$ grows significantly when $\alpha_z = 0$, some of the results below separate that case. For this case, we also caution that we are not convinced that no-correlation cases are realistic but do believe that, as an approximation, the case with $\alpha_{\text{non-spouse}} \to 0$ is much more plausible than with $\alpha_{\text{spouse}} \to 0$.

***Summary of results of unrestricted correlation exploration on*** $e(t)$***.*** As Figs. S15 and S16 highlight, all the periods identified in the main text ($\text{P}_0$ to $\text{P}_5$) are robust. This means that regardless of the values of $\alpha_{\text{non-spouse}}$ and $\alpha_{\text{spouse}}$, these periods exist. However, for some of the cases we consider unrealistic $\alpha_{\text{spouse}} < \alpha_{\text{non-spouse}}$, the specific times that characterize $\text{P}_1$ and $\text{P}_2$ are affected. Concretely, the elevated $e(t)$ period $\text{P}_1$ lasts a longer part of 2020 and the subsequent depressed $e(t)$ period $\text{P}_2$ is shorter. However, the time frames for the remaining periods are all robust to correlation changes.

Another important shared feature we find is that, as anticipated by our results in Sec. S2.3.4, decreasing $\alpha_z$ for any $z$ leads to the increase of $e(t)$.

***Particular results for fixed*** $\alpha_{\text{non-spouse}}$***, changing*** $\alpha_{\text{spouse}}$***.*** Beyond the common features described above, Fig. S15 helps us highlight the effect of varying $\alpha_{\text{spouse}}$ from a greater to a lesser value than $\alpha_{\text{non-spouse}}$. From Fig. S15(top), which excludes $\alpha_{\text{spouse}} = 0$, we observe the change in temporal pattern of in 2020

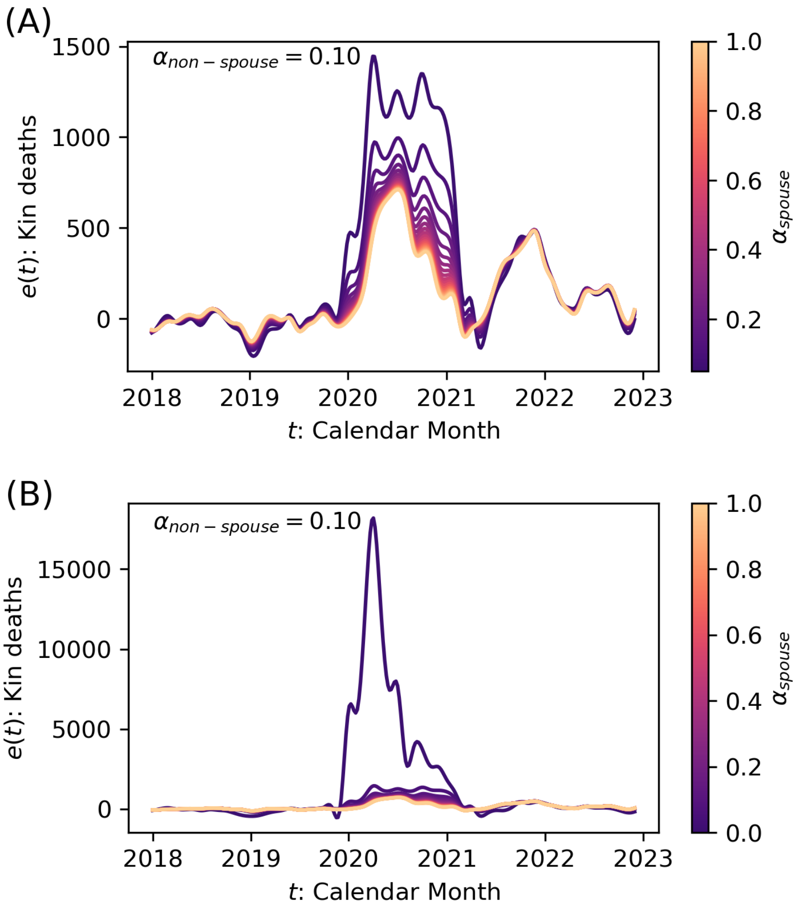

Figure S15: Exploration of the impact of $\alpha_{\text{spouse}}$ on LKDs, with $\alpha_{\text{non-spouse}} = 0.1$. We separate the exploration into two panels given the disproportionate impact of the case $\alpha_{\text{spouse}} = 0$, which obscures the behavior of $e(t)$ for other values of $\alpha_{\text{spouse}}$. In panel A ($\alpha_{\text{spouse}} > 0$), one can observe that in qualitative terms, $e(t)$ maintain the periods identified in Fig. 1 of the main text, although $P_1$ covers a greater proportion of 2020 as $\alpha_{\text{spouse}} \to 0$. The magnitude of $e(t)$ increases as $\alpha_{\text{spouse}}$ decreases. Furthermore, the pattern of peaks of $e(t)$ changes as $\alpha_{\text{spouse}}$ becomes sufficiently smaller than $\alpha_{\text{non-spouse}}$, dominating the overall signal. As discussed, however, this reversal of correlation values is unlikely. Panel B expands the results in panel A to include the case of $\alpha_{\text{spouse}} = 0$, which overwhelms the signal for $e(t)$. This is shown mainly for completeness, but the scenario is not plausible.

(periods $P_1$ and $P_2$) which occupy more of the year and show different peaks as $\alpha_{\text{spouse}} \to 0$. As explained above, in this limit, spouse overlaps dominate $e(t)$. The case when $\alpha_{\text{spouse}} = 0$, shown in Fig. S15(bottom), exhibits a prominent spike in 2020 that is not generally representative of other analyses performed in this study (it is due to some rare events with large obituary weights), supporting our claim that eliminating correlations altogether is unlikely to be reliable. The years 2021 and 2022 show little changes under varying $\alpha_{\text{spouse}}$, which is a reflection of the very limited contribution that spouse overlaps make to $e(t)$ in those years.

***Particular results for fixed*** $\alpha_{\text{spouse}}$***, changing*** $\alpha_{\text{non-spouse}}$***.*** Again, consistent with our description above, in Fig. S16 we observe the effect of varying $\alpha_{\text{spouse}}$ from a greater to a lesser value than $\alpha_{\text{non-spouse}}$. From Fig. S16(top), which excludes $\alpha_{\text{non-spouse}} = 0$, we observe that the increase of $\alpha_{\text{non-spouse}}$ eventually leads to a small signal of $e(t)$ which has a temporal pattern dominated by $e(t, z = \text{spouse})$ (with its corresponding changes for the time frames $P_1$ and $P_2$). The case of $\alpha_{\text{non-spouse}} = 0$ is presented in Fig. S16(bottom), and shows a more dramatic increase of $e(t)$ but no qualitative changes to the temporal pattern (the locations of yearly peaks and subpeaks remain the same, as do the time frames of all periods). The relative increase of the 2021 peak is slightly greater than the 2020 peak, driven by non-representative rare events.

***Unrestricted exploration of relationship contributions*** $\lambda(z)$***.*** The relative contributions of each relationship type $z$ to the total number of estimated LKDs (as defined in Eq. 2 of the main text) are also affected by writing correlations. As stated above, since a considerable majority of overlaps occur among non-spouses, the estimates of proportional contributions of spouses on overall to LKDs should, in reality, be a minority. Thus, estimates that show a large spousal contribution to overall LKDs are unlikely to be realistic and are merely presented for completeness of analysis. The time frame for our estimation of the relationship contributions corresponds to the period starting in March 2020 and ending at the end of 2022.

Focusing on the quantitative details, in Fig. S17, we show a matrix of plots of $\lambda(z)$ for $z = \text{spouse (SP), genetic (GN), and affinal (AF)}$ relationships, where the values of $\alpha_{\text{spouse}}$ and $\alpha_{\text{non-spouse}}$ are both combined in pairs, each taking values from $0.05, 0.25, 0.50, 0.75$, and 1. The diagonal (from top left to bottom right) contains the cases with $\alpha_{\text{spouse}} = \alpha_{\text{non-spouse}}$. Based on our arguments about the relation between spousal and non-spousal writing correlation, we lend more credence to the upper right corner of the matrix of plots where, in all cases, spousal LKDs are the minority contribution, an effect that becomes more pronounced as $\alpha_{\text{spouse}}$ becomes larger.

***Unrestricted exploration of gender pairs*** $\lambda(y)$***.*** The relative contributions of each gender pairing $y$ to the total number of LKDs are also affected by writing correlations. The relevant results are presented in Fig. S18. The matrix of plots is arranged in the same way as for Fig. S17, with the values of $\alpha_{\text{spouse}}$ and $\alpha_{\text{non-spouse}}$ combined in pairs from the values $0.05, 0.25, 0.50, 0.75$, and 1. Drawing a comparison with the case for $z$, the contributions for each gender pairing are more evenly spread, with the only exception across the matrix of plots being the male to female pairing which shows a slightly elevated proportional contribution. No other major patterns arise from this analysis, particularly on the upper right corner of the matrix of plots that we find more plausible from the standpoint of writing correlation.

***High and low correlation scenarios.*** The relative contributions of LKDs ($\lambda(z)$ and $\lambda(y)$) under these more restricted ranges of writing correlations can be extracted from the results for the unrestricted analysis above. The scenarios are defined by the intervals of $\alpha_{\text{spouse}}$ and $\alpha_{\text{non-spouse}}$ indicated in Tab. S7.

In order to assess the ranges of values that $\lambda(z)$ reach in the low or high correlation scenarios, we present Fig. S19. Panel A shows the low correlation case, where the contribution of genetic kin is by far the largest, with values ranging from just over 0.6 to close to 0.7 (detailed ranges described in main text Sec. 2.3). Affinal kin has the second largest contribution and spouses the lowest. In the high correlation scenario, genetic kin continues to have the largest contribution, although its dominant value is reduced somewhat in comparison to the low correlation case. The roles of affinal and spousal kin are reversed in comparison to the low correlation scenario. The reason why these changes occur is because, as $\alpha_{\text{non-spouse}}$ increases, it makes estimates of LKDs under the assumption that the non-spouse overlaps we detect are a greater fraction of all overlaps actually occurring.

Similarly, we present results for the relative contributions $\lambda(y)$ of gender pairs $y$ under the low and high correlation scenarios in Fig. S20. The relative contributions of different pairs are more comparable with one another across scenarios. Some notable features are that $M \to M$ pairs are consistently the lowest contribution, $M \to F$ is larger in contribution than $F \to M$, and that $F \to F$ is dominant in the low correlation scenario but becomes the second lowest contribution in the high correlation case.

[66] Béla Bollobás. *Modern graph theory*, volume 184. Springer Science & Business Media, 2013.

| $\sigma$ | $n_\sigma^{(h_o)}$ | $n_\sigma^{(h_1)}$ |
|---|---:|---:|
| 2 | 30,037,978 | 54,630,760 |
| 3 | 33,595,405 | 25,024,451 |
| 4 | 38,677,402 | 25,093,926 |
| 5 | 25,073,378 | 12,516,415 |
| 6 | 12,209,849 | 5,354,579 |
| 7 | 5,404,744 | 2,125,519 |
| 8 | 2,526,963 | 921,420 |
| 9 | 1,215,090 | 411,810 |
| 10 | 684,350 | 225,880 |
| 11 | 341,332 | 109,627 |
| 12 | 222,874 | 64,844 |
| 13 | 89,904 | 26,829 |
| 14 | 41,532 | 13,677 |
| 15 | 27,541 | 8,519 |
| 16 | 19,207 | 5,424 |
| 17 | 10,811 | 2,392 |
| 18 | 4,892 | 1,316 |
| 19 | 2,432 | 514 |
| $\geq 20$ | 5,410 | 2,200 |

Table S6: **Numbers of people in US households of size $\sigma$.** The middle column, $n_\sigma^{(h_o)}$, represents numbers for people where the householder is not married, and the right column, $n_\sigma^{(h_1)}$, corresponds to the case when the householder is married. The numbers are derived from the Public Use Microdata Sample from the American Community Survey, 5-year estimates of 2022, using the US Census data tool interface. The tool generates the numbers using household and person weights and, therefore, they are not exact counts but estimates. In the analysis through the US Census tool, we define population strata from ACS variables in the following way: AGEP variable separates householders into ages $< 65$,
$\geq 65$, or the entire population, NP variable into household sizes from 2 to 20, MAR variable separates householders into married and not currently married, and RELSHIPP variable separates house dwellers into spouse, non-spouse relatives including self, and non-relative. Although unmarried householders should not appear as married, the Census data tool produces small non-zero numbers of spouses, likely due to a combination of issues such as weight estimates or sampling limitation. Thus, both $n_\sigma^{(h_o)}$ and $n_\sigma^{(h_1)}$ are constructed by adding the appropriate counts of spouses and non-spouse relatives including self that the Census interface generates. The reported data can also be found in [53].

| Correlation scenario | $\alpha_{z=\text{spouse}}$ | $\alpha_{z=\text{non-spouse}}$ |
|---|---|---|
| High | [0.75,0.95] | [0.50,0.75] |
| Low | [0.75,0.95] | [0.05,0.25] |

Table S7: Definition of the low and high correlation scenarios for writing correlation in obituaries. These scenarios are meant to be representative of different closed ranges of the correlation space $(\alpha_{\text{spouse}}, \alpha_{\text{non-spouse}})$ that are distinct from each other in a qualitative way. While we always assume that $\alpha_{\text{spouse}}$ is relatively large, we explore distinct ranges of values for $\alpha_{\text{non-spouse}}$ that could produce different qualitative results for LKDs. These ranges reflect what is known about who typical writers of obituaries are [80, 81], i.e. most probably the children of decedents. Note that, consistent with these ideas, we always work with $\alpha_{\text{spouse}} > \alpha_{\text{non-spouse}}$.

| Calculation method | spouses | non-spouses |
|---|---|---|
| Observed overlap count | 26.34% | 73.66% |
| $\sum_t \sum_g \Theta[s(g,t,z)]$ with $\alpha_z = 1$ | 29.35% | 70.65% |

Table S8: Percentages of spouse and non-spouse overlaps based on different forms of overlap counting. These percentages are obtained under the application of $\Delta\theta = 30$ days, $m_o = 4$, $D = 0$, and $\hat{d}^* = 5$. The top row is based on the direct count of observed overlaps given $z$, with no weighing scheme applied and no subtraction of a statistical baseline. The second row shows percentages of observed overlaps above 0 under the weighing scheme with a correlation $\alpha_z = 1$ for both spouses and non-spouses. The correlation values in this case are not particularly realistic because they assign equality to spouses and non-spouses when this is unlikely to be a valid assumption. However, in both cases, it is clear that non-spouse overlaps are much more common than spouse overlaps. Excess overlap contributions can be seen in Fig. S17.

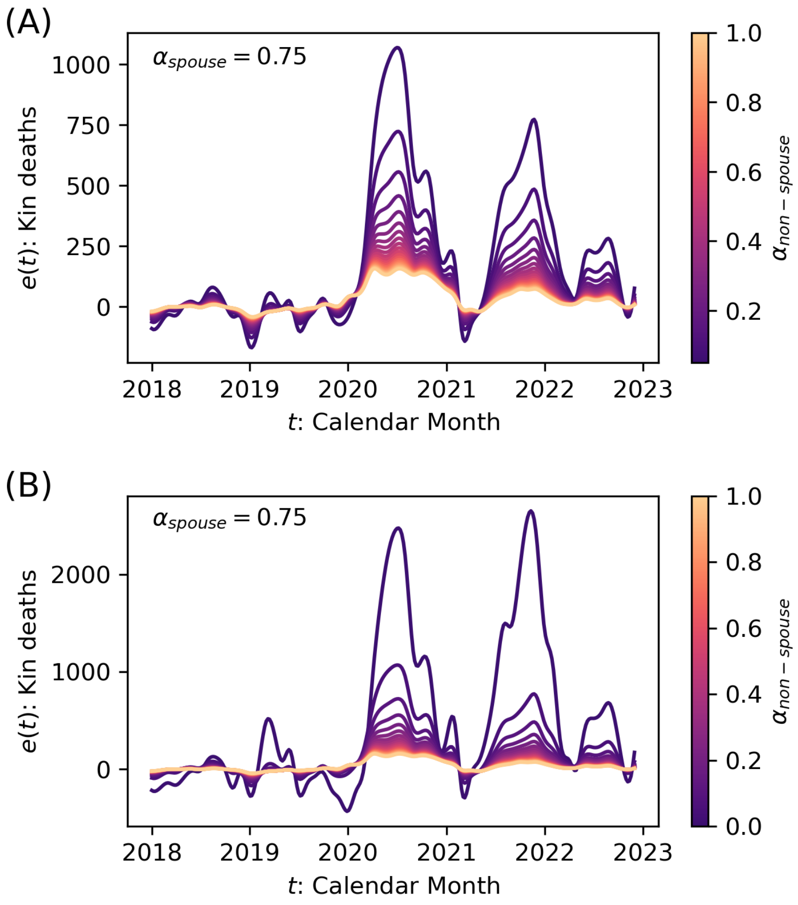

Figure S16: Exploration of the impact of $\alpha_{\text{non-spouse}}$ on LKDs, with $\alpha_{\text{spouse}} = 0.75$. We separate the exploration into two panels given the large impact of the case $\alpha_{\text{non-spouse}} = 0$. In panel A ($\alpha_{\text{non-spouse}} > 0$), we find that the periods identified in Fig. 1 of the main text are preserved, and the decrease of $\alpha_{\text{non-spouse}}$ leads to an increase in the magnitude $e(t)$. At the opposite limit, as $\alpha_{\text{non-spouse}} \to 1$, the magnitude of $e(t)$ decreases but does not vanish and, as $\alpha_{\text{non-spouse}}$ becomes sufficiently larger than $\alpha_{\text{spouse}}$, the pattern of peaks changes to reflect the dominant peaks due to spouse overlaps. However, this limit is unlikely to be realistic. Panel B expands the results in panel A to include the case of $\alpha_{\text{non-spouse}} = 0$. Although here too the magnitude of $e(t)$ becomes large, unlike the case of $\alpha_{\text{spouse}} = 0$, it is not implausible. However, given the arguments we lay out with regards to the relation between sampling rates and $\alpha_{\text{non-spouse}}$, we believe that $\alpha_{\text{non-spouse}} > 0$.

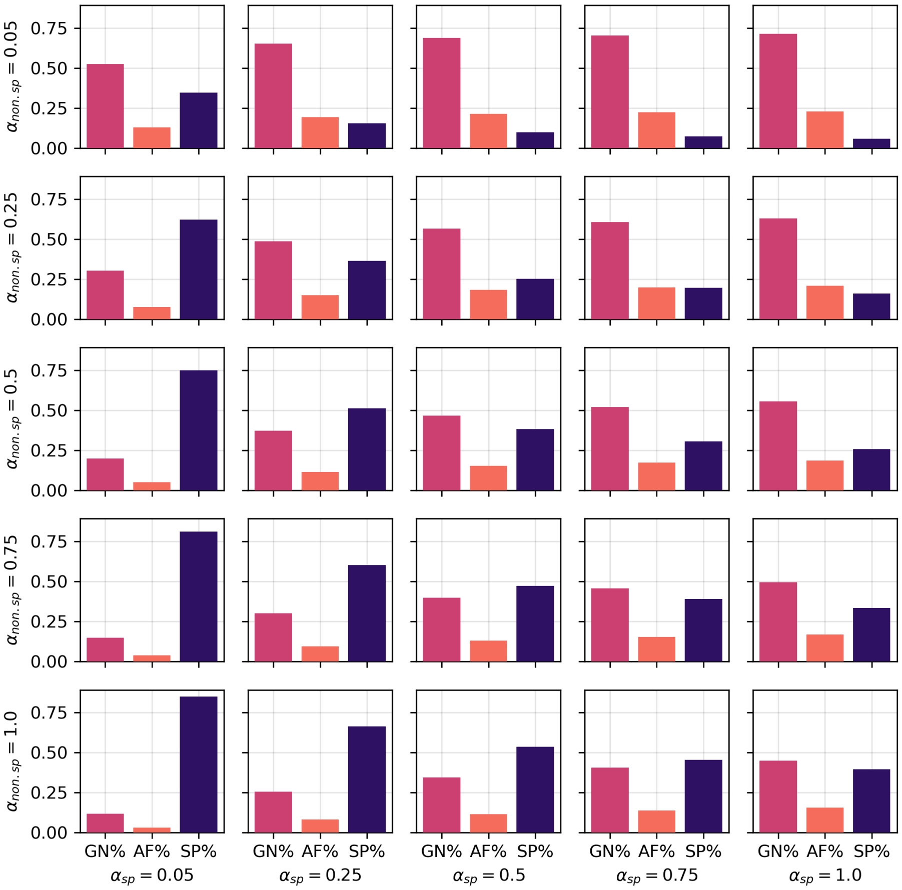

Figure S17: Matrix of plots for the relative cumulative LKD contributions $\lambda(z)$ for relationship type $z$ and specific writing correlations. These contributions are calculated through Eq. 2 of the main text. Across a large selection of correlation values, $z =$ GN kin is the largest contributor to LKDs. In contrast, $z =$ AF and $z =$ SP are generally smaller contributions, although which of them is the largest contribution varies depending on the specific values of writing correlations. In the scenarios we consider most plausible, $\lambda(\text{AF}) > \lambda(\text{SP})$. Given what is known about obituary writers [80, 81], we believe that $\alpha_{\text{spouse}} >$ $\alpha$non-spouse is reasonable (upper right triangle of the matrix of plots) and, in contrast, $\alpha_{\text{spouse}} <$ $\alpha$non-spouse is not. In the unlikely case of $\alpha_{\text{spouse}} <$ $\alpha$non-spouse, $\lambda(\text{SP})$ can emerge as the largest contributor.

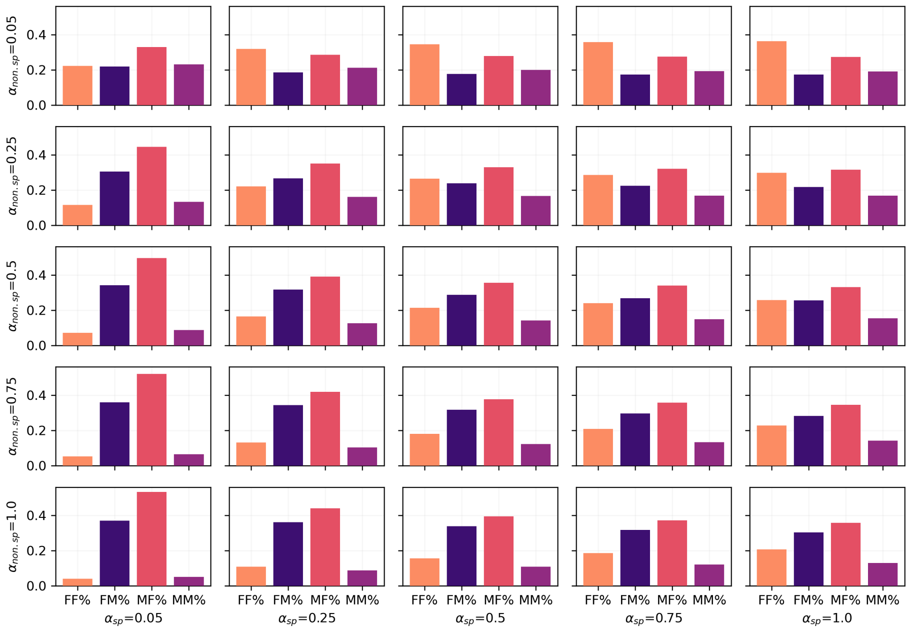

Figure S18: Matrix of plots for the relative cumulative LKD contributions $\lambda(y)$ for gender pair $y$ and specific writing correlations. These contributions are calculated as explained in main text, using Eq. 2 specialized to $y$. In general, there are no overtly dominant gender pairs, particularly in the region of correlations $\alpha_{\text{spouse}} > \alpha_{\text{non-spouse}}$, which is the most reasonable (upper right triangle of the matrix of plots). For small correlations $\alpha_{\text{non-spouse}}$ and large correlations $\alpha_{\text{spouse}}$, we find that female secondaries seem to be more common. Only in the unrealistic range of correlations where $\alpha_{\text{spouse}} < \alpha_{\text{non-spouse}}$, we find particularly dominant contributions to LKDs from the gender pairings $F \to M$ and $F \to M$.

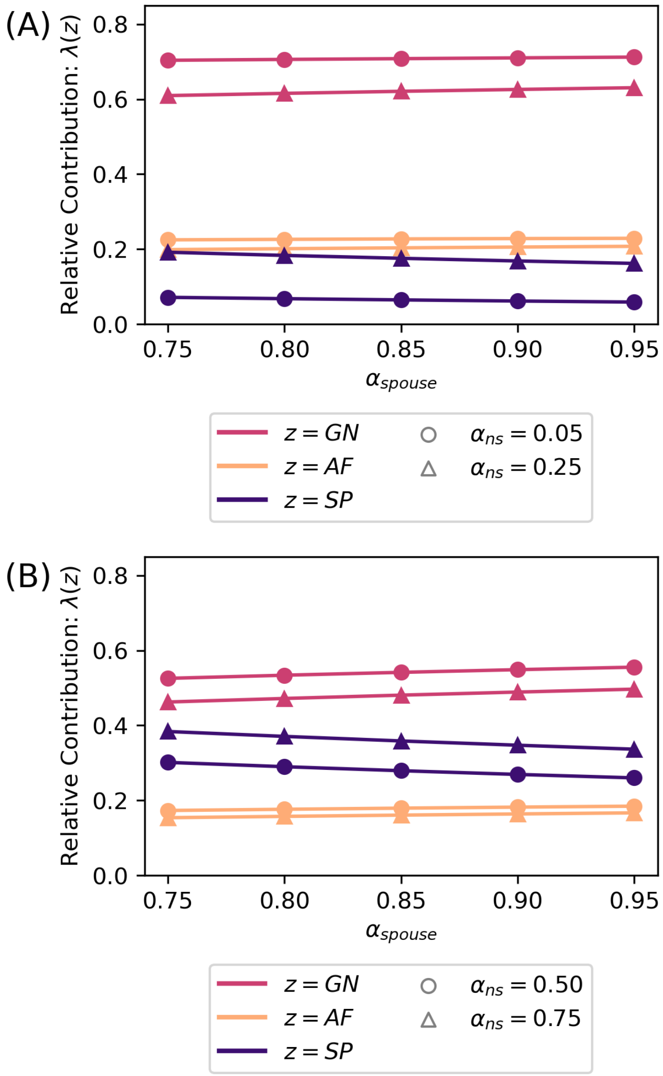

Figure S19: Correlation scenario results for $\lambda(z)$ for $z = \text{GN}$, AF, and SP. Panel A contains low correlation scenario values and Panel B high correlation scenario values. In both cases, $z = \text{GN}$ show the largest contribution to LKDs. In the low correlation scenario, $z = \text{AF}$ is generally the second largest contribution whereas $z = \text{SP}$ is the smallest contribution. In contrast, the roles are reversed for these two types of relationships in the high correlation scenario, where $z = \text{SP}$ is the second largest contribution and $z = \text{AF}$ is the smallest contribution. Note that the ranges within which each scenario is defined produce greater changes in contributions for $z = \text{GN}$ and SP, but $\lambda(\text{AF})$ changes very little between the smallest and largest values of $\alpha_{\text{non-spouse}}$ under either scenario.

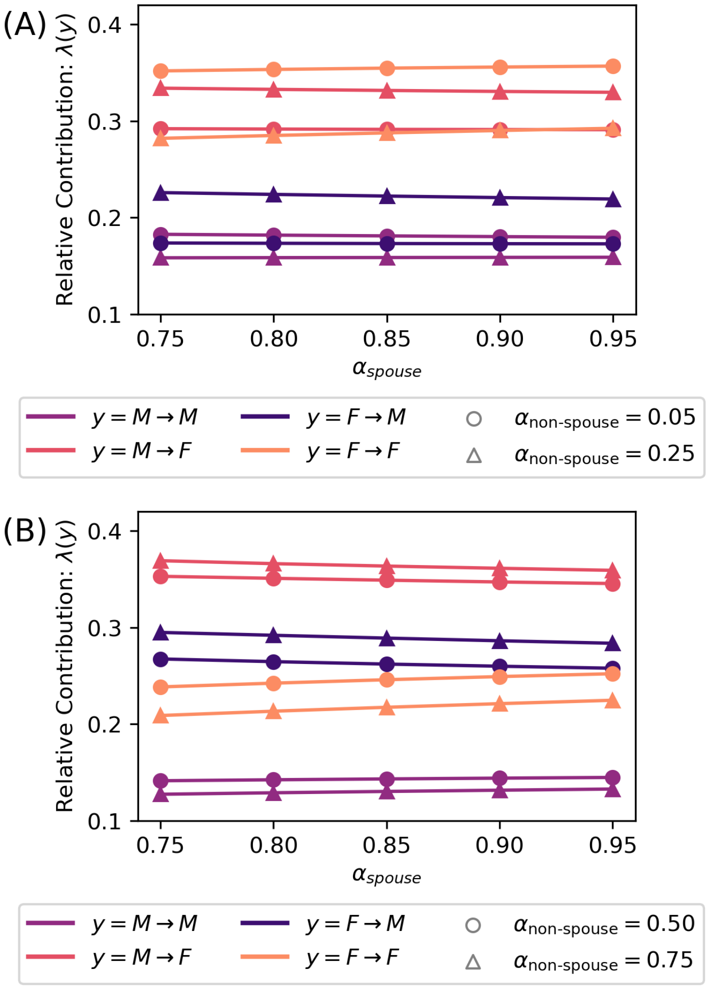

Figure S20: Correlation scenario results for $\lambda(y)$ for $y = M \to M$, $M \to F$, $F \to M$, and $F \to F$. Panel A contains low correlation scenario values and Panel B high correlation scenario values. A consistent feature is that $M \to M$ pairings are generally the lowest contributors to LKDs. In addition, among the $F \to M$ and $M \to F$ pairings, the latter is a greater contributor in both scenarios. The role of $F \to F$ changes the most, with $\lambda(F \to F)$ being the largest contribution under low correlations but the nest to lowest under high correlations.
